# SARS-CoV-2 Transmission in Alberta, British Columbia, and Ontario, Canada, January 1-July 6, 2020

**DOI:** 10.1101/2020.07.18.20156992

**Authors:** Isaac Chun-Hai Fung, Yuen Wai Hung, Sylvia K. Ofori, Kamalich Muniz-Rodriguez, Po-Ying Lai, Gerardo Chowell

**Author notes:** Correspondence should be addressed to Dr. Isaac Chun-Hai Fung, Department of Biostatistics, Epidemiology and Environmental Health Sciences, Jiann-Ping Hsu College of Public Health, P.O. Box 7989, Georgia Southern University, Statesboro, GA 30460-7989, USA. Isaac Chun-Hai Fung: https://orcid.org/0000-0001-5496-2529. Yuen Wai Hung: https://orcid.org/0000-0002-1171-109X. Sylvia Ofori: https://orcid.org/0000-0002-9961-9975. Kamalich Muniz-Rodriguez: https://orcid.org/0000-0002-4786-9438. Po Ying Lai: https://orcid.org/0000-0002-6156-1878. Gerardo Chowell: https://orcid.org/0000-0003-2194-2251.

## Abstract

**Objective:** To investigate COVID-19 epidemiology in Alberta, British Columbia and Ontario, Canada.

**Methods:** We calculated the incidence rate ratio (January 1—July 6, 2020) between the 3 provinces, and estimated time-varying reproduction number, *R_t_*, starting from March 1, using EpiEstim package in R.

**Results:** Using British Columbia as a reference, the incidence rate ratios in Alberta and Ontario are 3.1 and 4.3 among females, and 3.4 and 4.0 among males. In Ontario, *R_t_* fluctuated ~1 in March, reached values >1 in early and mid-April, then dropped <1 in late April and early May. *R_t_* rose to ~1 in mid-May and then remained <1 from late May through early July. In British Columbia, *R_t_* dropped <1 in early April, but it increased towards the end of April. *R_t_* <1 in May while it fluctuated around 1.0 in June and early July. In Alberta, *R_t_* > 1 in March; *R_t_* dropped in early April and rose again in late April. In much of May, *R_t_* <1, but *R_t_* increases in early June and fluctuates ~1 since mid-June.

**Conclusions:** *R_t_* wavering around 1.0 indicated that three provinces of Canada have managed to achieve limited onward transmission of SARS-CoV-2 as of early July 2020.

## Introduction

In 2020, the pandemic of coronavirus disease 2019 (COVID-19), caused by the severe acute respiratory syndrome virus 2 (SARS-CoV-2), spread across Canada. The first imported case was presented in a Toronto hospital on January 23. 2020.^1,2^ On March 13, Quebec was the first province to declare a public health emergency;^3^ four days later, Alberta, British Columbia and Ontario also declared public health emergencies (Table S1).^4^ As of July 10, a cumulative total of 107,126 cases, including 8759 deaths, have been reported in Canada.^5^ As seen in the Canadian government’s outbreak update,^6,7^ the national epidemic trajectory appears to have stabilized in early July, but spatial-temporal pattern displays substantial heterogeneity across provinces.

Epidemiologists have explored various aspects of the pandemic in Canada, including data availability,^8^ syndromic surveillance,^9^ disease burden and mortality,^10^ as well as the epidemiology in specific settings or subpopulations, such as nursing homes,^11^ and intensive care units.^12^ Mathematical modelers have also made projections of epidemic trajectories to assist Ottawa and the provincial governments in their decision-making.^7,13-16^ Against the backdrop of recent literature, our study provides a unique perspective by estimating and interpreting the time-varying reproduction number, *R_t_*, of SARS-CoV-2 at both provincial and subprovincial levels in 3 Canadian provinces.

This study described the COVID-19 pandemic as it unfolded in Alberta, British Columbia, and Ontario. Our objectives are to compare COVID-19 incidence rates from January 1 to July 6, 2020, and to estimate the time-varying reproduction number, *R_t_*, of SARS-CoV-2 since the beginning of March 2020.

## Methods

### Scope and data sources

We investigated the descriptive epidemiology and the *R_t_* of SARS-CoV-2 in Alberta, British Columbia, and Ontario, and their public health subdivisions, using publicly available line lists of COVID-19 case data downloaded from provincial government websites. Alberta, British Columbia, and Ontario were chosen for this analysis given their line list data availability. These datasets contained no personal identifiers to protect patients’ privacy. Population data by age and sex for Alberta, British Columbia and Ontario was downloaded from Alberta government website.^17^ At the time of our study, Quebec has not yet made the line list of cases publicly available and was not included herein. As of early July, the cumulative number of cases in the other provinces were relatively small and therefore those provinces were not included. The Canadian government’s national line list dataset provided the accurate episode time in epidemiologic weeks instead of dates, and thus could not be analyzed using our method here. All the subprovincial incidence curves and *R_t_* trajectories were presented in the Supplementary Materials.

### Timeframe

The timeframe of this study started on January 1, 2020 (the accurate episode date of the earliest confirmed case in Ontario) and ended on July 5 (Alberta and Ontario) or 6 (British Columbia), the last episode date or reported date in each of our datasets. This timeframe is characterized by importation of cases from overseas followed by community transmission while social distancing measures were gradually introduced. March 17 was the day when the public health emergency was declared in Alberta, British Columbia and Ontario.^4^ By early July, some mandatory social distancing measures remain implemented therein.

The timeframe for *R_t_* estimation began with March 1. We did not estimate *R_t_* in January and February because the small number of cases in Ontario and British Columbia during that time would lead to very uncertain *R_t_* estimates. No cases were reported in Alberta before March.

### Ontario dataset

The Ontario dataset included cases until the end of July 5, 2020 and was downloaded on July 6, 2020, at 5.34pm (EDT).^18^ In Ontario, the subprovincial unit is a Public Health Unit (PHU). The Ontario dataset contained the following variables: accurate episode date, case reported date, test reported date, specimen date, age group, gender, case acquisition information, clinical outcome, outbreak-related (yes or no), reporting PHU, and the address, city, postal code, website, latitude and longitude of the reporting PHU.

In contrast to the Alberta and British Columbia datasets, which did not contain information on how the case-patients acquired the virus, the Ontario dataset provided information on how a case-patient acquired the virus (“Case Acquisition Info”): 1761 travel-related cases, 10839 “close contact” cases, 6687 cases with “no epi-link”, 14259 outbreak-related cases, 1120 cases with “no info-missing” and 1280 cases with “no info-unknown”. In our *R_t_* estimation, we categorized all travel-related cases as “imported” cases and all others as “local” as per user instructions for the EpiEstim package. Please note that to run the EpiEstim package, the first case(s) on the first day of a time series must be rendered as “imported” cases. Thus, if the first case(s) of a time series was not travel-related, we manually denoted them as “imported” cases so that *R_t_* can be estimated using the EpiEstim package.

### Alberta dataset

The Alberta dataset included cases until the end of July 5, 2020 and was downloaded on July 7, 2020 at 7.59am (Eastern Daylight Time, EDT).^19^ In Alberta, each subprovincial unit is a Health Services Zone. The Alberta dataset contained the following variables: date of report, health services zone, gender, age group, case status (clinical outcome) and case type (confirmed or probable). The Alberta dataset contained 7678 confirmed cases and 711 probable cases; both types are included in the main analysis. In the sensitivity analysis, only confirmed cases were analyzed.

### British Columbia dataset

The British Columbia dataset included cases until July 6, 2020 and was downloaded on July 7 at 8.01am (EDT).^20^ In British Columbia, each subprovincial unit is a Health Authority. The British Columbia dataset contained the following variables: date of report, health authorities, sex, age group and classification reported (“Epi-linked” versus laboratory-diagnosed). The British Columbia dataset contained 2970 laboratory-diagnosed cases (aka confirmed cases) and 8 “epi-linked” cases (aka probable cases); both are included in the main analysis. In the sensitivity analysis, only laboratory-diagnosed cases were analyzed.

### Incidence rate ratio

The incidence rate was calculated by dividing the cumulative number of cases in a province by its total population in 2019. Given that the timeframe of our datasets is essentially the same (with only a difference in 1 day), we did not use person-time as the denominator. We used British Columbia as the reference, and calculate the incidence rate ratio (IRR) of Alberta and that of Ontario. We did not calculate the 95% confidence interval for the estimates, because the estimates were true population estimates and not sample estimates, as we used data from province-wide surveillance systems that cover everyone in the province.

### Case fatality ratio

We limited our analysis to individuals whose clinical outcomes are known (either deaths or recovered), and used the following simple estimator to obtain an approximation of the case fatality ratio: case fatality ratio = number of deaths / (number of deaths + number of recovered).^21^

### Time-varying reproduction number

In contrast with the basic reproduction number, *R*_0_, which represents the average number of secondary cases generated by an infectious individual in a totally susceptible population in the absence of interventions or behavioral changes, *R_t_* is a time-varying indicator which represents the average number of secondary cases per infectious individual in a population as the epidemic unfolds in the presence of interventions and behavioral changes. *R_t_* >1 indicates sustained transmission and epidemic growth; *R_t_* <1 indicates unsustainable transmission and epidemic decline. As the epidemic runs its course, it is possible to quantify *R_t_* over time after accounting for reductions in susceptibility in the population as more people acquire immunity through natural infection, behavior changes in the population, and interventions that mitigate the transmission rate.^22^

### Instantaneous reproduction number method implemented in R package EpiEstim

There are multiple statistical methods available for *R_t_* estimation,^23,24^ one of which is the instantaneous reproduction number method implemented in the R package EpiEstim.^25,26^ This method is considered to be well-suited to near-real-time estimation of *R_t_*, and is sensitive to signals of changes in transmission potential given recent implementation and cessation of interventions or behavioral changes.^23^ This method has been applied in various studies to estimate the *R_t_* of SARS-CoV-2 in different locations globally.^27-34^ A summary of the instantaneous reproduction number method is provided in the Online Supplementary Materials.

### Software version

Data management and analysis (except for maps) was performed using Microsoft Excel and R version 3.6.2 (R Core Team, Vienna, Austria). Version 2.2-3 of the EpiEstim package was used. Maps were made in R version 3.5.1 (R Core Team, Vienna, Austria).

### Ethics

The Georgia Southern University Institutional Review Board made a non-human subjects determination for this project (H20364), under the G8 exemption category.

## Results

The dates of the first cases and the total number of cases in our datasets by each subprovincial public health unit in Alberta, British Columbia and Ontario are presented in Table S2.

### Incidence rate ratio and case-fatality ratio at the provincial level

Table S3 presents the number of cases per 100,000 population, stratified by sex/gender and age groups (see Tables S4-S6 for details). Interestingly, the incidence rates in Alberta and Ontario were higher than those in British Columbia, which served as the reference group. Among females, the respective IRR in Alberta and Ontario were 3.1 (187.3/59.9) and 4.3 (259.9/59.9). Among males, the respective IRR in Alberta and Ontario were 3.4 (196.2/57.2) and 4.0 (196.2/57.2). This may reflect a more aggressive testing strategy in Alberta and Ontario. As of July 11, 2020, Alberta and Ontario had the highest rate of testing in Canada (102,299 and 110,612 tests per million population, respectively) whereas in British Columbia the rate was only 36,059 tests per million population.^6^ In both provinces, anyone who feels needing a test, including asymptomatic individuals, can get a test without referral. For example, Ontario waived the requirement to get a referral to qualify for testing on May 24, 2020, and increased testing capacity to over 20,000 a day.^35^ Likewise, Alberta started testing asymptomatic cases around mid-May. The alternative hypothesis was that this might reflect a genuine difference across the 3 provinces in their epidemiology of COVID-19. Tables S7 and S8 present the clinical outcomes of cases in Alberta and Ontario. As expected, the elderly has a high case-fatality ratio than other age groups, as described in a recent study by Bignami and Van Assche,^10^ and this is similar to observations in other parts of the world, such as England.^36^ Likewise, as in other countries, there had been outbreaks in long-term care facilities in Canada.^11,37,38^

### Epidemic curve and R_t_

The accurate episode date of the first two cases in Ontario was January 1, 2020, one without epi-link and one being a close contact. The “no epi-link” case was a male in his 80s whose specimen date was April 23 and whose test result was reported on April 24 (case reported on the same date). The “close contact” case was a male in his 50s whose specimen date was May 15 and whose test result was reported on May 17 (case reported on the same date). A third case was a male in his 50s whose accurate episode date was January 10 and was a “close contact” case but whose specimen was taken on June 9 (test reported and case reported on the next day). This cluster of 3 cases had episode dates before the first two reported cases in Ontario (date of report: January 23, 2020) whose accurate episode dates were January 21 and 22 respectively.^1,2^ In Ontario (Figure 1), *R_t_* fluctuated around 1 in March, rose to >1 but <1.25 in early and mid-April, then dropped <1 in late April and early May. *R_t_* rose to ~1 in mid-May and then remained <1 from late May through early July.

**Figure 1.**
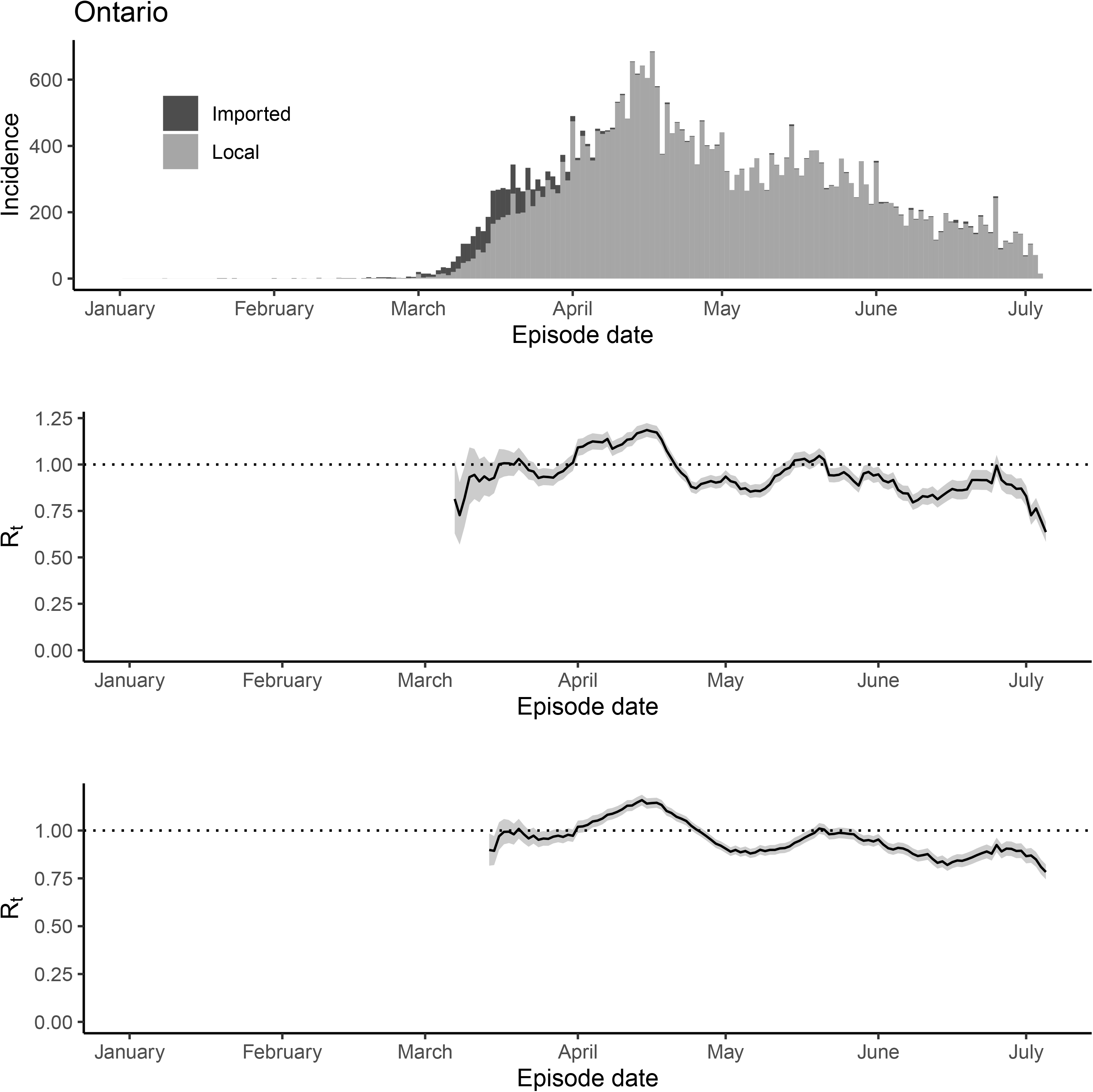
The epidemic trajectory of coronavirus disease 2019 in Ontario, January 1-July 5, 2020: Upper panel: Daily number of new imported and local cases by date of symptom onset (aka accurate episode date) (upper panel); *R_t_* with a 1-week window (middle panel); *R_t_* with a 2-week window (lower panel).

A map showing the cumulative number of cases over time by PHU in Ontario is presented in Figure 2, and a similar map showing the cumulative number of deaths is in Figure S1. Epidemic trajectories in selected PHUs are as follows. In City of Toronto PHU (Figure S2) and its neighbors, namely, Durham Regional PHU (Figure S3), Peel Regional PHU (Figure S4), and York Regional PHU (Figure S5), we observed similar tragectories: *R_t_* ≥ 1 in sustained period of time from late March to mid-April. *R_t_* subsided to <1 in late April and early May but rose to ~1 in mid-May. *R_t_* decreased to <1 consistently in early June but it was on an increasing trend towards early July. Slightly further away from Toronto, in Halton Regional PHU (Figure S6), *R_t_* fluctuates around 1 through the study period. It was <1 in much of June. In City of Hamilton PHU (Figure S7), *R_t_* fluctuates around 1 through the timeframe of this study, except for mid-May when *R_t_* reached a peak of 2 (1-window estimate). In Region of Waterloo PHU (Figure S8), *R_t_* fluctuates around 1 in March but increases steadily to >1 in mid-April, followed by a continuous drop to <1, until late May when *R_t_* rebounded to around 1 in June. In Niagara Region PHU (Figure S9), *R_t_* fluctuates around 1 across the timeframe of this study, except for late May when there was a surge in cases leading to *R_t_* >1 in late May followed by a trough <1 in June. In Ottawa PHU (Figure S10), *R_t_* was <1 in late March but it increased to slightly >1 in April. It maintained <1 in much of May and fluctuates around 1 in June. For the other PHUs from Algoma PHU to Windsor-Essex County Health PHU, see Figures S11 to S35.

**Figure 2.**
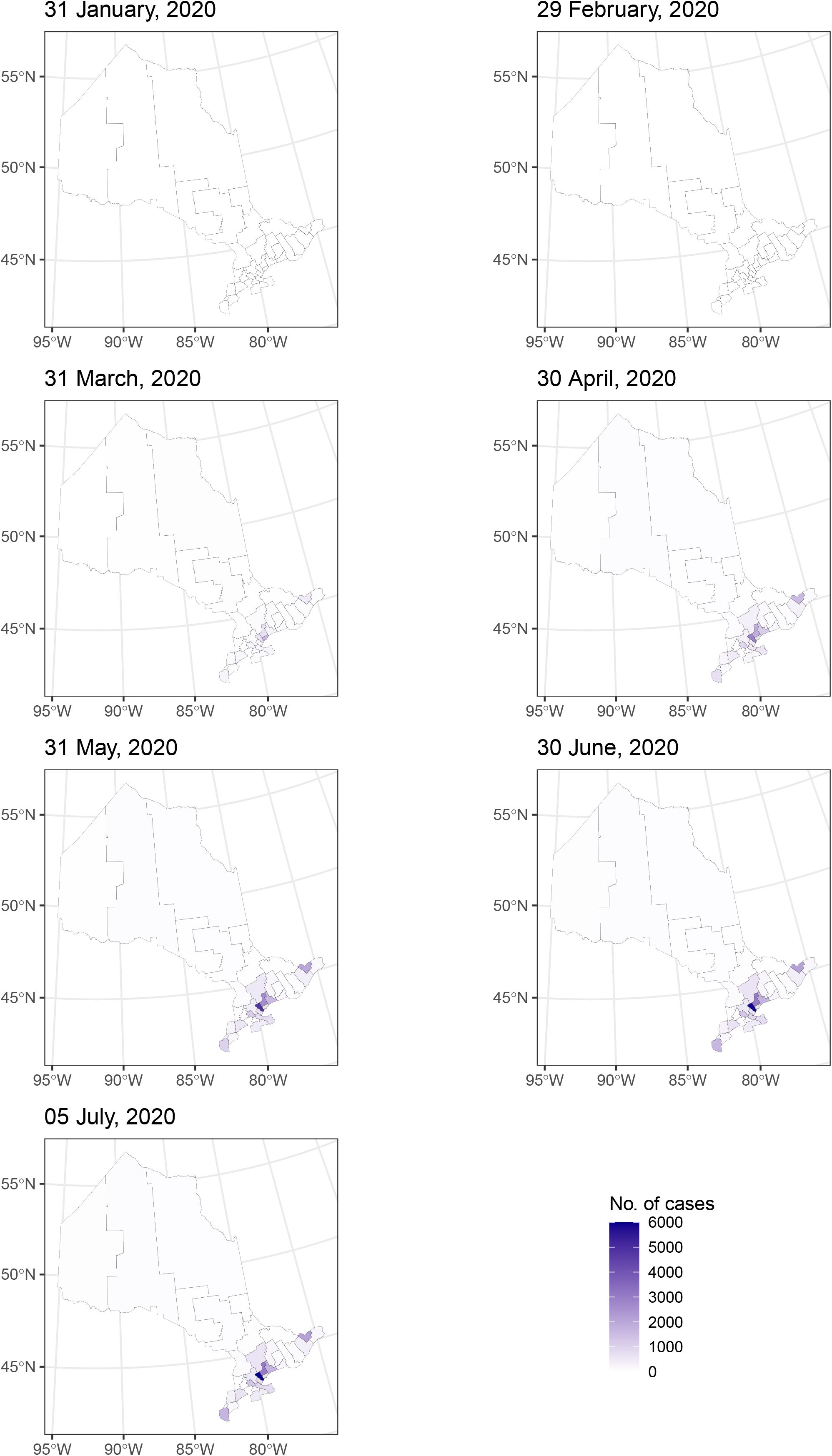
Map of cumulative number of cases in Ontario by public health unit by the end of month and on July 5, 2020.

The first case in British Columbia was reported by Vancouver Coastal Health Authority on January 26, 2020. In British Columbia, *R_t_* dropped below 1 the first time in early April, but it increased towards the end of April. For most of May, *R_t_* <1 while it fluctuated around 1 in June and early July (Figure 3). Figure 4 is a map showing the cumulative number of cases in British Columbia by Health Authority over time. Below are epidemic trajectories in selected Health Authorities. In Vancouver Coastal Health Authority (Figure S36), *R_t_* was ~2 in mid-March and dropped to 1 by April and below <1 in mid-April but increased in late April to ~1. It fluctuates <1 for much for May but rose to ~2 in mid-June, followed by fluctuation around 1 in late June and early July. In Fraser Health Authority (Figure S37), *R_t_* fluctuated between 1.5 and 2 in mid-March before dropping to ~1 through April. In May it fluctuated <1 but increased to 1 and fluctuated around 1 in June and early July. For Interior, Northern and Vancouver Island Health Authorities, see Figure S38 to S40. As a sensitivity analysis we analyzed the data excluding 8 probable cases (“epi-linked cases”) (Table S9); focusing at the 2970 confirmed cases, the results in *R_t_* are similar (Figure S41).

**Figure 3.**
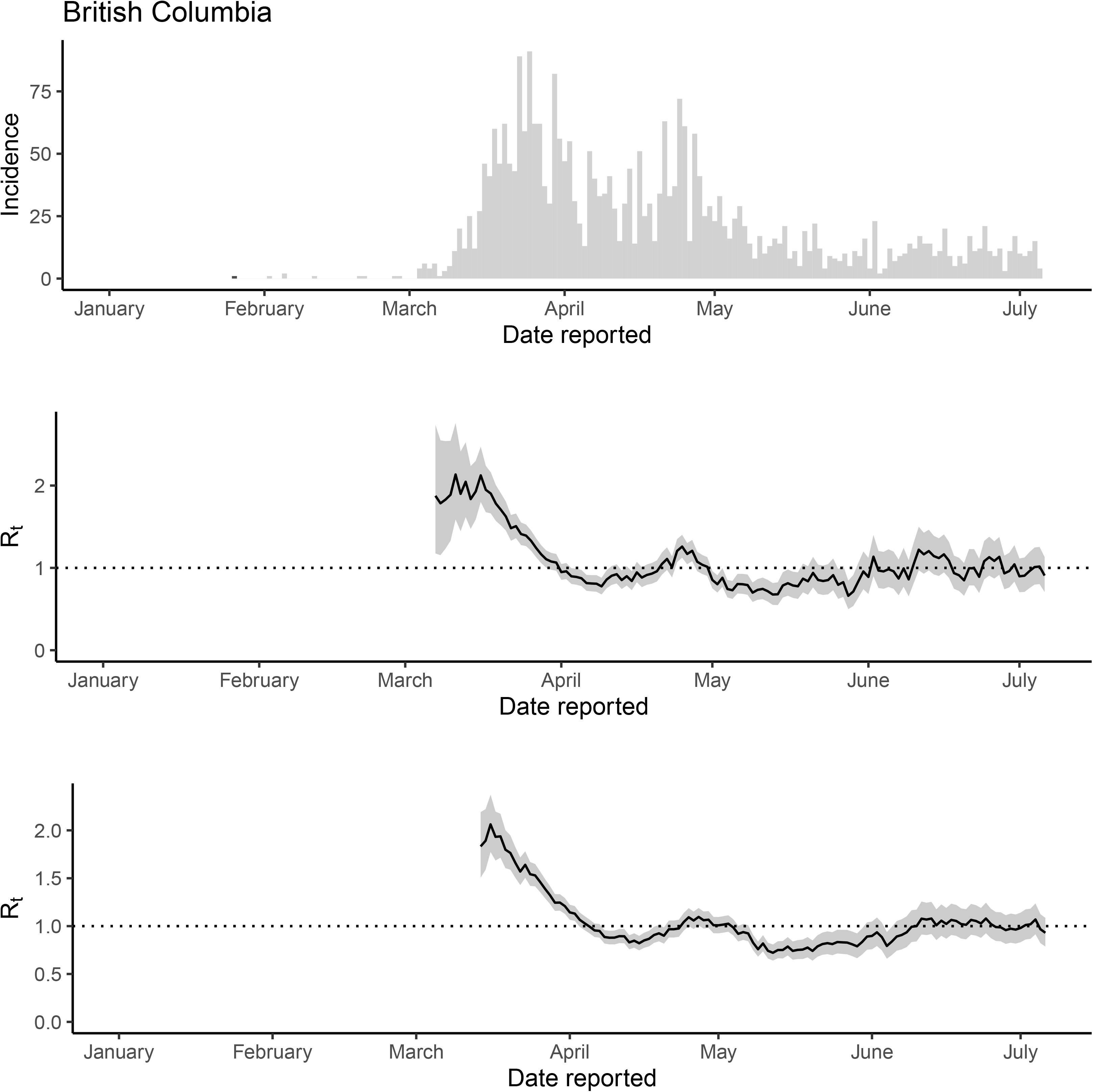
The epidemic trajectory of coronavirus disease 2019 in British Columbia, January 1-July 6, 2020: Daily number of new cases by date of report (upper panel); *R_t_* with a 1-week window (middle panel); *R_t_* with a 2-week window (lower panel).

**Figure 4.**
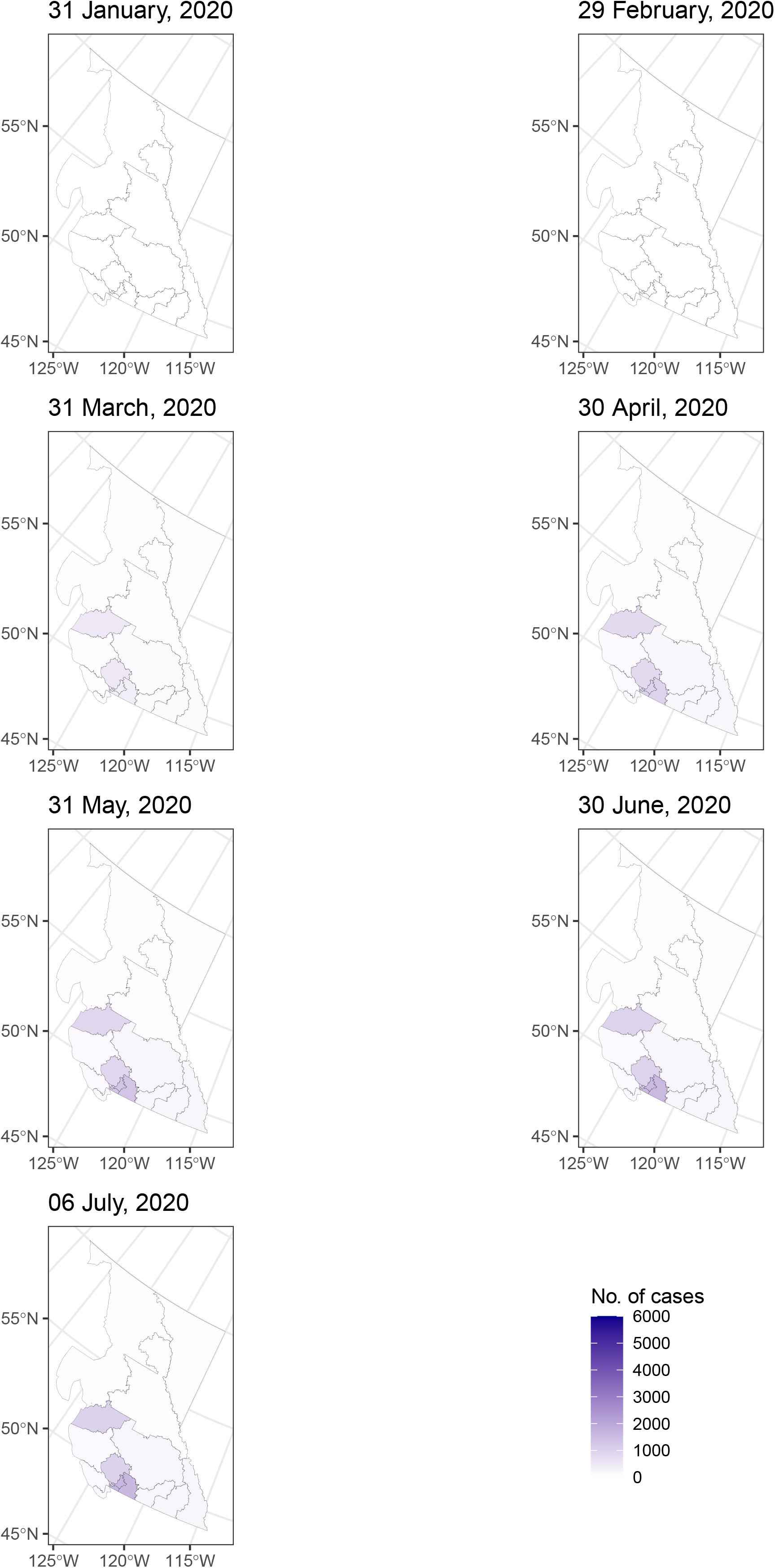
Map of cumulative number of cases in British Columbia by health authority by the end of month and on July 6, 2020.

The first case in Alberta was reported in Calgary Zone on March 6, 2020. In Alberta, *R_t_* > 1 in March, corresponding to reaching a peak in incidence in late March and early April. A small dip in *R_t_* happened in early April and *R_t_* rose again in late April, as the incidence increased and reached a peak in late April.

In much of May, *R_t_* <1, but *R_t_* increases in early June and fluctuates around 1 (slightly above 1) since mid-June (Figure 5). Figure 6 is a map showing the cumulative number of cases in Alberta by Health Services Zone over time, while Figure S42 is a similar map showing the cumulative number of deaths. Below are epidemic trajectories in selected Health Services Zones. In Calgary Zone (Figure S43), *R_t_* began at ~2 in mid-March and dropped to ~1 in early April and rose again to ~1.5 and maintained >1 in the second half of April. In May and early July, *R_t_* was <1, but increased to ~1 from mid-June through early July. In Edmonton Zone (Figure S44), it fluctuates around 1.5 in late March, dropped to 1 in early April and <1 in mid-April. *R_t_* rose to ~1 in late April and then maintained at a <1 level for much of May. *R_t_* increased to ~2 in late May and early June, and then gradually dropped to ~1 in early July. In South Zone (Figure S45), Rt fluctuated around 1 in late March and early April and rose to ~3 in the second half of April as there was an outbreak from mid-April to early May. Rt below <1 in much of May and fluctuated around 1 in much of June and made a sharp increase to ~2 in early July. For Central and North Zones, see Figures S46 and S47. As a sensitivity analysis we analyzed the data excluding 711 probable cases; focusing at 7678 confirmed cases only, the results in *R_t_* are similar (Figure S48).

**Figure 5.**
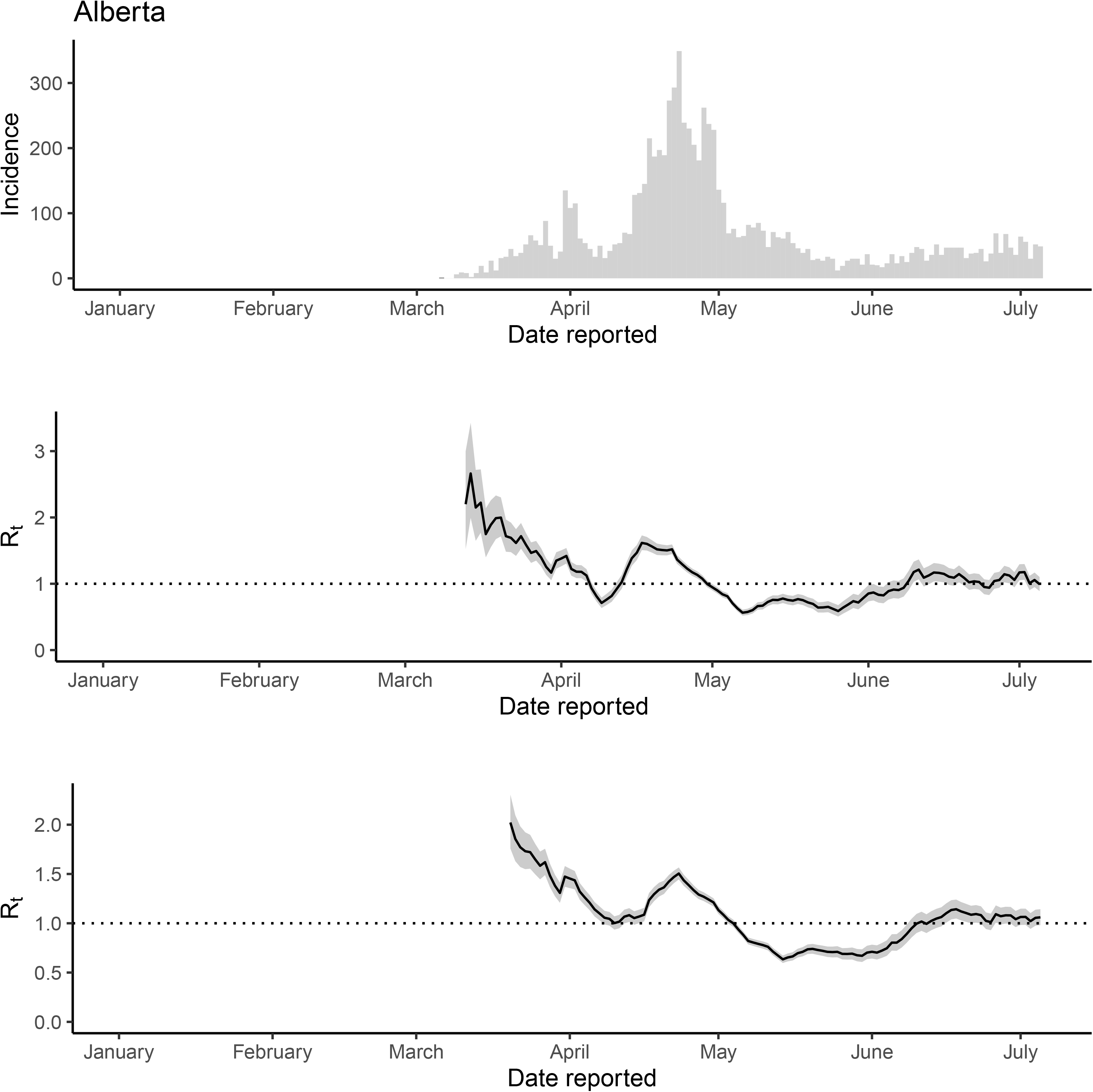
The epidemic trajectory of coronavirus disease 2019 in Alberta, January 1-July 5, 2020: Daily number of new cases by date of report (upper panel), *R_t_* with a 1-week window (middle panel) and *R_t_* with a 2-week window (lower panel).

**Figure 6.**
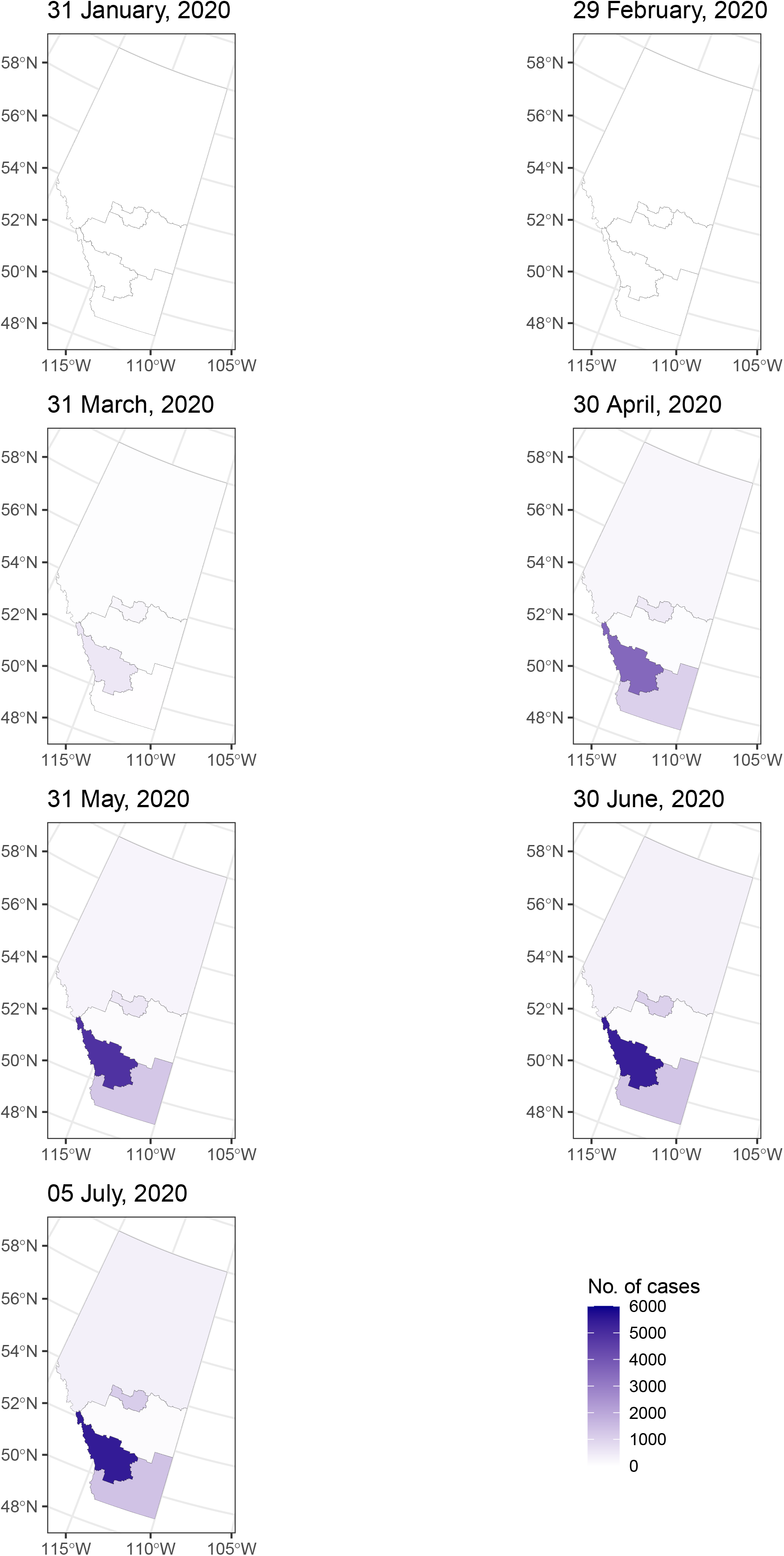
Map of cumulative number of cases in Alberta by health service zone by the end of month and on July 5, 2020.

### Specimen collection date in Ontario dataset

Among the 35948 case-patients in Ontario, the median delay in specimen collection for testing is 1 day after symptom onset (accurate episode date), with an interquartile range from 0 to 5 days. A decrease over time in the average time difference between date of symptom onset and the date of specimen taken, and the variance of that time difference (Figure 7 Panel a) represents the improvement in the testing capacity allowing individuals to get tested promptly after symptom onset. As seen in Figure 4a, the delay on March 1 would be 7.4 days and the delay was close to 0 around June 21. See Appendix for details of the regression model. The longest time difference (an outlier) was a delay of 151 days (that might suggest that these were serological tests and not polymerase chain reaction tests,^39^ but the dataset do not provide further details). A total of 305 (0.8%) individuals do not have specimen dates. There were 2391 (6.7%) whose specimens were collected prior to date of symptom onset, i.e. (accurate episode date - specimen date) < 0. Of these 2391 individuals, 926, 766, 397 and 302 individuals developed symptom 1, 2, 3 or ≥4 days, respectively, *after* having specimen taken for testing (Table S10). Most of these 2391 case-patients were outbreak-related (n=1473) or close contacts of cases (n=619), while 28 were travel-related. However, 211 of them did not have any epi-link, and 24 and 36 were without case acquisition info (“missing” or “unknown” respectively) (Table S11). This is not surprising, since Ontario enhanced testing capacity for testing asymptomatic people at risk of exposure. Of these 2391 individuals whose specimens taken prior to symptom onset, except for 8 who did not have a test report date, 1463 (61%) had their tests reported the same day as their symptom onset; 307 (13%), 1 day after symptom onset, 385 (16%), ≥2 days later, and 228 (9.5%) had their test reported *before* symptom onset (Table S10). Of these 2391 individuals, 1641 (69%) had their case reported the day they developed symptoms, 337 (14%) on the next day of symptom onset, 413 (17%), ≥2 days after symptom onset (Table S10).

**Figure 7.**
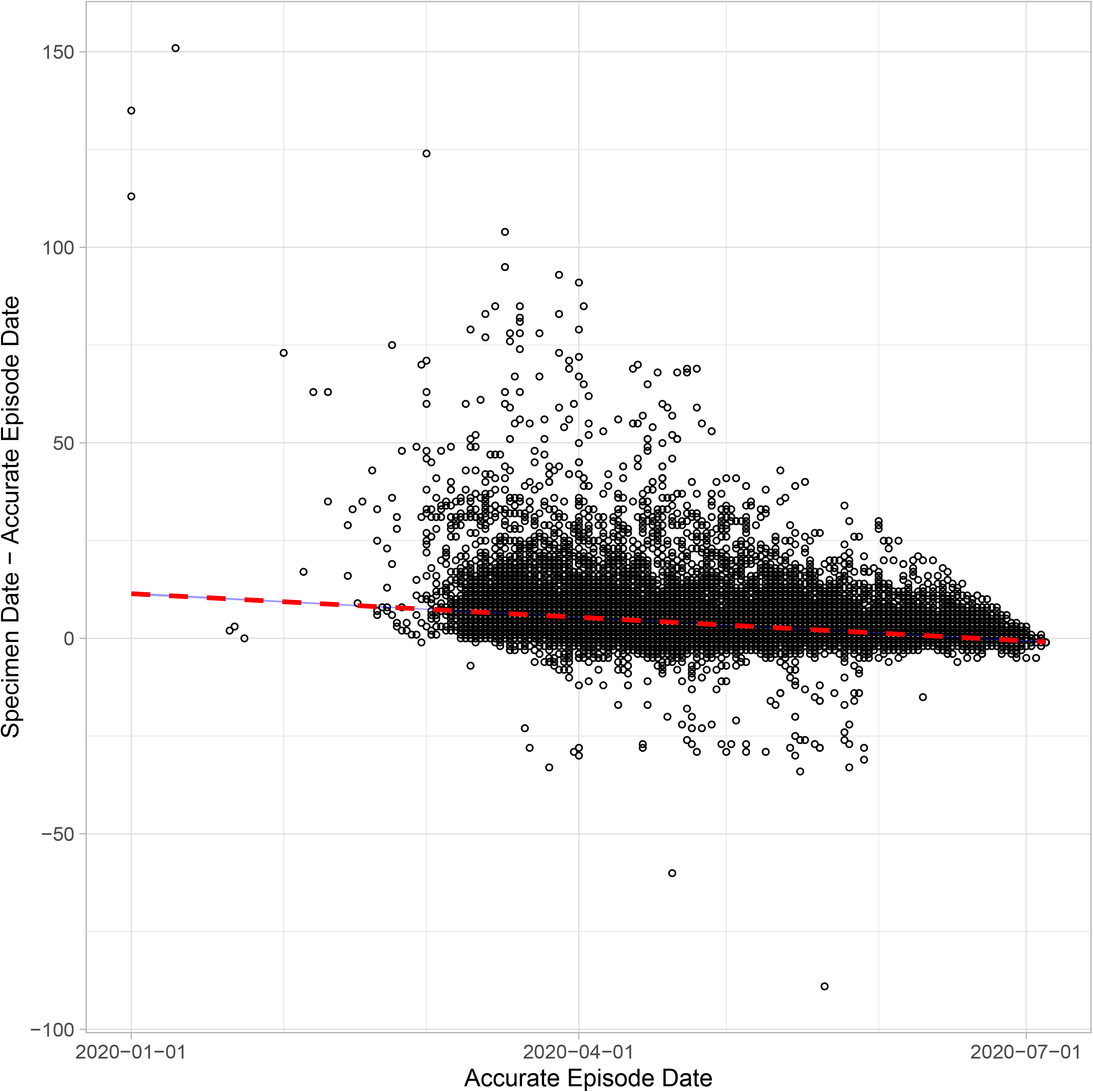
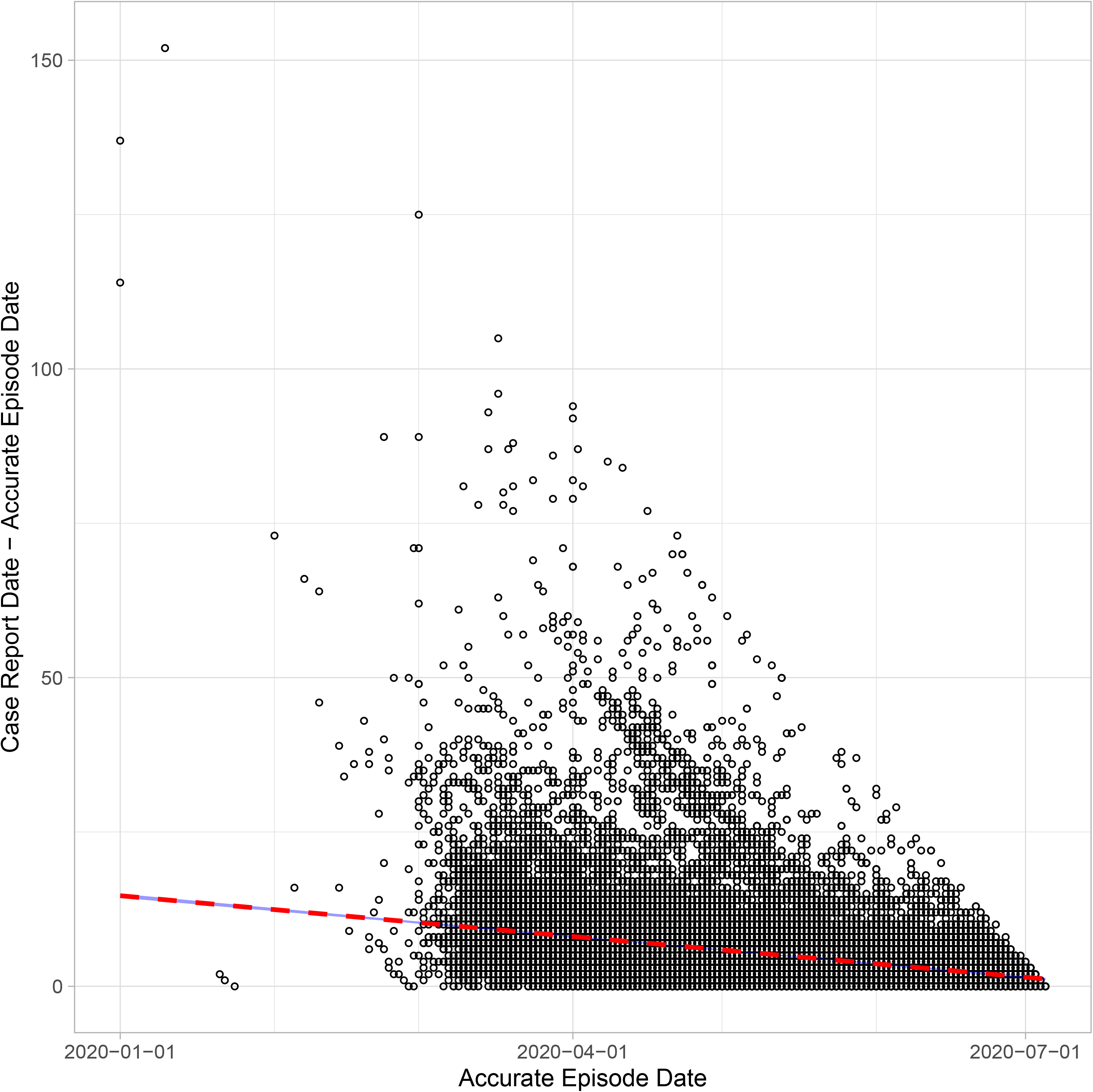
**(a)** Scatter plot of the time difference between symptom onset (accurate episode date) and testing (specimen date) of each case-patient in Ontario over time (accurate episode date), Jan 1-July 6, 2020. **(b)** Scatter plot of the time difference between symptom onset (accurate episode date) and case report date of each case-patient in Ontario over time (accurate episode date), Jan 1-July 6, 2020.

### Delay in test report and case report in Ontario dataset

The delay in test report decreased over time. On March 1, the delay was approximately 11 days, while on July 5, it was about 1 day (Figure S49). The delay in case report has also decreased over time. On March 1, the delay was approximately 10 days, while on July 5, it was about 1 day (Figure 7 panel b). See Appendix for details of the regression models.

## Discussion

Alberta, British Columbia and Ontario, Canada, appeared to have managed to achieve limited transmission of SARS-CoV-2 with R_t_~1 at the time of writing in early July 2020 (Figures 1, 3, 5), through a combination of multiple interventions, including community social distancing, increased testing of individuals at higher risk or suspected of SARS-CoV-2 infection, and isolation of COVID-19 cases. Canada’s experience with SARS epidemic in 2003 may have contributed to the coordinated government’s response in COVID-19. The 2003 SARS epidemic in Canada resulted in a set of recommendations to federal, provincial, and territorial leader, and in particular, led to the creation of the Public Health Agency of Canada which has been leading the response to COVID-19.^40^

Our results show that since mid-March in the 3 Canadian provinces under study, SARS-CoV-2 has been spreading with an *R_t_* <2 that is smaller than the estimated R_0_ of >2^41,42^ For the majority of the time period under study, *R_t_* fluctuated around 1. Our analysis of the COVID-19 epidemic by Canadian subprovincial units present a similar picture as our province-level analysis with few exceptions. Nowhere in Canada experienced the trauma that residents in Wuhan, Milan and New York City had experienced. While Canadian provinces have substantial administrative authority over COVID-19 response, the federal and provincial government have collaborated to produce a coordinated response to the pandemic, as evidenced by the similar COVID-19 response measures in the three provinces, as well as the united politicians’ communications on the nature and severity of the COVID-19 pandemic, and the necessity of social distancing across the different Canadian political parties.^43^ As the three provinces began reopening businesses, services, and public spaces, all three provinces have been gradually relaunching by stages which depended on local infection rate and the capacity of the healthcare system. ^44-46^

Our study has several limitations. First, our *R_t_* estimation for Alberta and British Columbia relied on data by reporting date and not date of onset, as the latter was unavailable to us. Only Ontario provided the accurate episode date. Future research can study the epidemic data by date of onset when it is available to researchers to better estimate *R_t_*. Second, the number of reported cases per day in the Alberta and British Columbia datasets would be influenced by the testing rate, since limited testing capacity could lead to underdiagnosis. We did not attempt to do a nationwide analysis for Canada because the testing criteria has not been consistent across Canadian provinces. Third, our estimates are contingent that the testing ratio does not change significantly over the study period. However, provincial governments might have been ramping out testing capacity during the early epidemic phase in their jurisdiction. Fourth, reporting delays are known to vary over time and could influence the observed trajectory of the epidemics by date of report in Alberta and British Columbia. For Ontario, our results showed that the delay to testing has been reduced over time and Ontario achieved testing on the same or next date since symptom onset in June. It is possible that similar improvement in reduction in testing delay may have happened in Alberta and British Columbia, but we do not have such data. Fifth, improvement in testing over time could potentially reduce generation time over time. As we do not have data on infector-infectee pairs, we cannot estimate if serial interval has changed over time. In our *R_t_* estimation, our serial interval distribution is parametrically determined *a priori* based on the literature.^47^ Sixth, the Ontario dataset is by date of symptom onset and therefore the most recent data points may be lower than what they should have been, because there could be cases who had not yet been reported to the provincial government. Thus, the low *R_t_* estimate for Ontario in the 1^st^ week of July may be an artifact due to right-censoring of the data. Seventh, we obtained an approximation of the case-fatality ratio using a simple estimator by restricting the analysis to cases with known outcomes. There are estimation methods that are more statistically advanced,^21,48,49^ but they are less accessible to practitioners of applied epidemiology and they are beyond the scope of this paper.

## Conclusions

In conclusion, Alberta, British Columbia, and Ontario managed to keep the *R_t_* of SARS-CoV-2 wavering around 1, with our recent estimates indicating sporadic episodes of sustained transmission in Alberta and British Columbia where *R_t_* fluctuates around 1 or slightly >1 in the first week of July. In Ontario, *R_t_* may have been suppressed <1 in early July. Sustained social distancing interventions and further limiting case importations could potentially suppress the epidemic by driving *R_t_* <1 for a sustained period of time.

## Data Availability

All the data used are publicly available data downloaded from websites of the provincial governments of Alberta, British Columbia and Ontario. The URL links to those websites are provided in the reference list of the manuscript.

## Acknowledgements

G.C. received support from NSF grant 1414374 as part of the joint NSF-NIH-USDA Ecology and Evolution of Infectious Diseases program. I.C.-H.F. received salary support from the Centers for Disease Control and Prevention (CDC) (19IPA1908208). This article is not part of I.C.-H.F’s CDC-sponsored projects.

## Disclaimers

The opinions expressed in this article do not represent the official positions of the Centers for Disease Control and Prevention or the United States Government.

## Author Contributions

Dr. Fung is the first and corresponding author. Dr. Hung, Ms. Ofori, Ms. Muniz-Rodriguez and Ms. Lai contributed equally as co-second authors. Dr. Chowell serves as the senior author.

### Instantaneous reproduction number method implemented in R package EpiEstim

We summarized the instantaneous reproduction number method below, of which detailed description has been made available elsewhere.^4,5^ The instantaneous reproduction number is the ratio between the number of incident cases at time t, and the total infectiousness of all the infected individuals at time t: *R_t_* = *I_t_*/*Λ_t_*, where 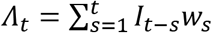. The total infectiousness is the sum of infection incidence up to the previous time point (*t-1*) weighted by an infectivity function *w_s_*. The infectivity function is a probability distribution that describes the average infectiousness profile of an individual after being infected, and it is usually represented by the serial interval distribution as an approximation to the generation time distribution. The number of new cases at time *t* is therefore a Poisson distribution with a mean of 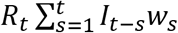. Given the reproduction number *R_t_*, and conditional on the time series of daily new cases, *I_0_, …, I_t-1_*, the likelihood of the incidence *I_t_* is therefore, 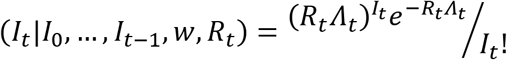 where 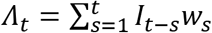. Therefore, 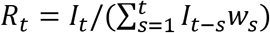. We can interpret the instantaneous reproduction number as the average number of secondary cases that an infectious person will infect at time *t*, if conditions in previous time points remain the same at time *t*. Given this formulation of the instantaneous reproduction number, *R_t_* is highly sensitive to fluctuation in the incidence data. To minimize *R_t_* variation, the EpiEstim package allows user to specify a time window over which the *R_t_* estimate is assumed to be constant. The window is of size *τ* and ends at time t. If transmissibility is assumed constant over time from (*t*-*τ*+1) to *t*, and is denoted by a reproduction number, *R_t_*,*_τ_* the likelihood of the incidence during the time period, from *I*_(_*_t-τ+_*_1)_ to *I_t_*, conditional on incidence prior to the time period, from *I_0_*, to *I_(t-τ)_*, is found to be:^5^

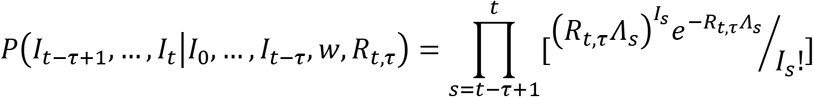

The EpiEstim package utilized a Bayesian framework with a Gamma-distributed prior for *R_t,τ_*, and the analytical expression of the posterior distribution of *R_t_* and estimated its median, the variance and the 95% credible interval (CrI).^5^

Given the small number of cases in January and February and thus wide uncertainty bounds of the *R_t_* estimates, we presented *R_t_* estimates from March to early July. *R_t_* estimates of two respective time windows of 7 days and 14 days, over which the *R_t_* estimates is assumed to be constant, were presented with their 95% CrI, as in Muniz-Rodriguez et al.^14^ As a sensitivity analysis, the 14-day time window provides a more stable estimate as it would not be as responsive to sudden fluctuations in incident case count. We did not use a time window shorter than 7 days because such *R_t_* estimate would be very sensitive to daily incidence fluctuation. Furthermore, for surveillance data by date of report (Alberta and British Columbia), weekend effect is common given the 5-day work week, i.e., daily number of cases reported over Saturday and Sunday would be lower than those from Monday to Friday. Therefore, it is reasonable to use time-windows of 7 or 14 days.

Serial interval is assumed to follow a gamma distribution (mean: 4.60 days; standard deviation: 5.55).^15^

#### Map creation

The health region boundaries shapefile was downloaded at Statistics Canada at https://www150.statcan.gc.ca/n1/pub/82-402-x/2018001/hrbf-flrs-eng.htm. The health regions were based on 2016 Census geographic units. Maps were made in R version 3.5.1 (R Core Team, Vienna, Austria). The boundaries shapefiles are transformed by R packages rnaturalearth (version 0.1.0) and sf (version 0. 9.0), merged with the study data by health regions, and presented by R package ggplot2 (version 3.3.0). The maps show the cumulative number of cases and fatal cases on the last day of each month between January 2020 to June 2020, and the last date of the data (5 or 6 July 2020) with non-missing values.

### Time delay in specimen collection, test report and case report in Ontario, January 1-July 5, 2020

We ran linear regression models to obtain the trend line over time.

a. Time delay in specimen collection With (accurate episode date - specimen date) as the outcome variable (y) and accurate episode date as the predictor variable (x), the linear model is: y = −0.06627x + 1221.67026 Given that in R, date is counted from Jan 1, 1970 (when it is “day 0”), for January 1, 2020, x = 18262, y = 11.41 days; for March 1, 2020, x = 18322, y = 7.43 days; for June 21, 2020, x = 18434, y = 0.0062 day(s).
b. Time delay in test report With (accurate episode date - test report date) as y and accurate episode date as x, the linear model is: y = −0.08383x+1547.21994 For January 1, 2020, x = 18262, y = 16.31 days; for March 1, 2020, x = 18322, y = 11.29 days; for July 5, 2020, x = 18448, y = 0.72 day(s).
c. Time delay in case report With (accurate episode date - case report date) as y and accurate episode date as x, the linear model is: y = −0.07254+1339.42505 For January 1, 2020, x = 18262, y = 14.67 days; for March 1, 2020, x = 18322, y = 10.32 days; for July 5, 2020, x = 18448, y = 1.18 day(s).

## Supplementary Tables

**Table S1.** Timeline of COVID-19-related events in Alberta, British Columbia, and Ontario in 2020.

| Date<br>(yyyy-mm-dd) | Events | Reference |
| --- | --- | --- |
| 2020-01-25 | First case of COVID-19 in Canada was reported in Toronto, Ontario. | 33 |
| 2020-01-28 | First presumptive case of COVID-19 was reported in British Columbia. | 33 |
| 2020-03-05 | First case of community transmission in British Columbia was detected through the annual influenza surveillance program; first presumptive case of travel-related COVID-19 was reported in Alberta. | 33 |
| 2020-03-12 | Ontario ordered public school closure. | 34 |
| 2020-03-15 | First community transmission case was reported in Alberta. | 33 |
| 2020-03-16 | Change in Public Health Ontario Laboratory testing algorithm of COVID-19. | 35 |
| 2020-03-17 | Alberta, British Columbia and Ontario declare states of emergency due to COVID-19, ordered many businesses, including bars, cinemas and private schools, to close; first COVID-19 related death in Ontario | 36 |
| 2020-03-18 | Canada closed border to non-essential travels (including the United States) | 34 |
| 2020-03-19 | Alberta had its first death from COVID-19 | 33 |
| 2020-03-23 | Ontario ordered mandatory closure of non-essential businesses | 34 |
| 2020-03-26 | British Columbia placed restrictions on business operations, adopting approaches similar to Québec. | 33 |
| 2020-03-27 | Alberta ordered closure of all non-essential businesses. | 37 |
| 2020-03-30 | Ontario's Chief Medical Officer of Health strongly recommended all people over 70 and those with compromised immune systems stop going out. Everyone else in the province should stay home as much as possible to limit the spread of COVID-19. | 38 |
| 2020-04-06 | Canada's Chief Public Health Officer advised Canadians to wear "non-medical" masks to reduce the risk that asymptomatic people will spread SARS-CoV-2 | 38 |
| 2020-04-09 | Ontario broadened its criteria for testing people with symptoms of COVID-19 to include all hospital inpatients, long-term care residents, healthcare workers and caregivers, first responders, and residents of remote or Indigenous communities.<br>Ontario's Medical Officer of Health also directed that all staff and essential visitors to long-term care homes must wear personal protective equipment for the entirety of their shift or visit, regardless of whether the home is experiencing an outbreak.<br>All residents of British Columbia returning home from abroad would have to submit detailed quarantine plans or be taken to a federal quarantine site. | 38 |
| 2020-04-15 | In Ontario, all symptomatic staff and residents of long-term care facilities would be screened, and staff are no longer allowed to work in more than one home. | 38 |
| 2020-05-01 | Alberta started slowly reopening provincial parks and golf courses and would resume elective surgeries and other non-urgent medical services next week. | 38 |
| 2020-05-06 | British Columbia began to reopen its economy, easing lockdown restrictions on some summer sports activities, retail businesses and hair salons. | 38 |
| 2020-05-19 | Ontario began to ease pandemic restrictions, gradually resuming scheduled surgeries and reopening some businesses and services, including retail stores with street entrances, construction, library pickups and some outdoor sporting competitions. Workplaces could also gradually reopen, but people should work from home as much as possible. The province also expanded eligibility for testing to include anyone with symptoms of COVID-19.<br>British Columbia began to ease pandemic restrictions, allowing various businesses, restaurants, and childcare to reopen as long as social distancing (2m distance) is maintained. Restaurants could have guests up to 6 people in a group. | 32,39 |
| 2020-05-20 | Ontario recommended that anyone over age two should wear a face covering when riding public transit unless they have trouble breathing or removing a mask. Provincial officials also said that transit agencies should erect plexiglass barriers on vehicles, make hand sanitizer available and clear high- touch areas. | 38 |
| 2020-05-21 | Ontario Premier promised to ramp up testing in the next three to four weeks, but the province was only completing about half its target of 16,000 tests per day. | 38 |
| 2020-05-27 | Ontario extended pandemic restrictions to June 9 that would have expired on May 29. Under the extended orders, playgrounds and pools would remain closed, bars and restaurants may only provide takeout and delivery services, and people may not gather in groups larger than five people. | 38 |
| 2020-05-29 | Ontario started rolling out an expanded testing strategy for COVID-19, which included testing workers in key sectors and communities experiencing outbreaks, as well as asymptomatic individuals. Provincial labs hit testing targets of 16000 tests a day. | 38 |
| 2020-06-01 | The Canada federal government opened 38 national parks and 171 historic sites for hikes and other daytime activities while camping overnight was not yet permitted.<br>Ontario reopened provincial parks, drive-in movie theatres and batting cages.<br>British Columbia reopened schools on a part-time basis. | 38 |
| 2020-06-07 | Alberta began distributing 20 million non-medical masks at almost 600 drive-through locations across the province, including fast food restaurants. While mask use was not mandatory, Alberta's chief medical officer recommended people wear a non-medical mask when physical distancing is not possible. Calgary and Edmonton would distribute an additional 500,000 masks through the transit system. | 38 |
| 2020-06-12 | 24 of Ontario's 34 public health units started second phase of easing pandemic restrictions, and 10 other Greater Toronto and Hamilton Area and near the U.S. border would wait until new daily cases in those areas consistently decrease.<br>The second phase increased the limit on gatherings to 10 people and allowed selected personal care services, outdoor dining, and some outdoor recreational facilities to resume operations. | 40 |
| 2020-06-18 | Ontario began allowing visitors to long-term care and retirement homes that do not have active cases of COVID-19. | 40 |
| 2020-06-19 | Ontario allowed the following public health unit regions to move into stage 2: Durham region health department, Haldimand-Norfolk health unit, Halton region health department, Hamilton public health services, Lambton health unit, Niagara region public health department, and York region public health services | 40 |
| 2020-06-24 | Ontario allowed Peel public health and Toronto public health units to move into stage 2. British Columbia entered into phase 3 of reopening, which allowed overnight camping in parks. | 40,41 |
| 2020-06-25 | Ontario allowed the City of Windsor and all other parts of Essex County to reopen, except the Municipality of Leamington and the Town of Kingsville | 40 |
| 2020-07-01 | British Columbia allowed long-term care facility residents to have visit with one designated family member or friend. | 41 |

**Table S2.** Date of first cases and total number of cases as of July 5, 2020 (Alberta and Ontario) or July 6, 2020 (British Columbia), by provincial and subprovincial jurisdiction.

|  | Figures | Date of first case in<br>2020* | Total number of cases in the<br>datasets ** |
| --- | --- | --- | --- |
| **Alberta** | **5, 6, S42** | **Mar 6** | **8389** |
| Calgary Zone, AB | S43 | Mar 6 | 5440 |
| Central Zone, AB | S46 | Mar 11 | 93 |
| Edmonton Zone, AB | S44 | Mar 9 | 1128 |
| North Zone, AB | S47 | Mar 16 | 338 |
| South Zone, AB | S45 | Mar 16 | 1373 |
| unknown | Not applied | Not applied | 17 |
| **British Columbia** | **3, 4** | **Jan 26** | **2978** |
| Fraser Health Authority, BC | S37 | Feb 20 | 1570 |
| Interior Health Authority, BC | S38 | Feb 11 | 203 |
| Northern Health Authority, BC | S39 | Mar 9 | 65 |
| Vancouver Coastal Health Authority, BC | S36 | Jan 26 | 1008 |
| Vancouver Island Health Authority, BC | S40 | Mar 10 | 132 |
| **Ontario** | **1, 2, S1** | **Jan 1** | **35948** |
| Algoma Public Health Unit, ON | S11 | Mar 8 | 24 |
| Brant County Health Unit, ON | S12 | Mar 6 | 133 |
| Chatham-Kent Health Unit, ON | S13 | Mar 7 | 161 |
| Durham Region Health Department, ON | S3 | Feb 24 | 1715 |
| Eastern Ontario Health Unit, ON | S14 | Feb 25 | 166 |
| Grey Bruce Health Unit, ON | S15 | Mar 6 | 107 |
| Haldimand-Norfolk Health Unit, ON | S16 | Jan 1 | 430 |
| Haliburton, Kawartha, Pine Ridge District Health Unit, ON | S17 | Mar 2 | 200 |
| Halton Region Health Department, ON | S6 | Mar 2 | 771 |
| Hamilton Public Health Services, ON | S7 | Mar 1 | 845 |
| Hastings and Prince Edward Counties Health Unit, ON | S18 | Feb 1 | 44 |
| Huron Perth District Health Unit, ON | S19 | Mar 7 | 60 |
| Kingston, Frontenac and Lennox & Addington Public Health, ON | S20 | Mar 5 | 104 |
| Lambton Public Health, ON | S21 | Mar 6 | 286 |
| Leeds, Grenville and Lanark District Health Unit, ON | S22 | Mar 1 | 354 |
| Middlesex-London Health Unit, ON | S23 | Jan 24 | 630 |
| Niagara Region Public Health Department, ON | S9 | Mar 6 | 758 |
| North Bay Parry Sound District Health Unit, ON | S24 | Mar 9 | 35 |
| Northwestern Health Unit, ON | S25 | Mar 12 | 39 |
| Ottawa Public Health, ON | S10 | Feb 10 | 2117 |
| Peel Public Health, ON | S4 | Feb 24 | 5961 |
| Peterborough Public Health, ON | S26 | Mar 1 | 95 |
| Porcupine Health Unit, ON | S27 | Mar 5 | 67 |
| Renfrew County and District Health Unit, ON | S28 | Mar 10 | 29 |
| Simcoe Muskoka District Health Unit, ON | S29 | Mar 3 | 605 |
| Southwestern Public Health, ON | S30 | Mar 1 | 84 |
| Sudbury & District Health Unit, ON | S31 | Mar 3 | 67 |
| Thunder Bay District Health Unit, ON | S32 | Feb 29 | 93 |
| Timiskaming Health Unit, ON | S33 | Mar 10 | 18 |
| Toronto Public Health, ON | S2 | Jan 21 | 13431 |
| Region of Waterloo, Public Health, ON | S8 | Jan 1 | 1307 |
| Wellington-Dufferin-Guelph Public Health, ON | S34 | Feb 20 | 491 |
| Windsor-Essex County Health Unit, ON | S35 | Feb 23 | 1662 |
| York Region Public Health Services, ON | S5 | Feb 10 | 3059 |

\*The dates for Alberta and British Columbia are dates of report. The dates of Ontario are accurate episode dates. \*\* Cumulative number of cases as of July 5, 2020 (AB, ON) or July 6, 2020 (BC).

**Table S3.** Cases per 100,000 and incidence rate ratio between Alberta, British Columbia and Ontario by sex (or gender) and age group.

| Cases per 100,000 |  |  |  | Incidence Rate Ratio |  |  |
| --- | --- | --- | --- | --- | --- | --- |
| Female | AB | BC | ON | AB | BC | ON |
| <20 | 122.0 | 9.9 | 55.2 | 12.3 | Ref | 5.6 |
| 20-29 | 208.4 | 55.7 | 265.5 | 21.0 | 5.6 | 26.7 |
| 30-39 | 225.6 | 75.4 | 243.7 | 22.7 | 7.6 | 24.5 |
| 40-49 | 271.9 | 68.3 | 286.5 | 27.3 | 6.9 | 28.8 |
| 50-59 | 201.0 | 79.0 | 310.1 | 20.2 | 7.9 | 31.2 |
| 60-69 | 129.1 | 44.6 | 225.4 | 13.0 | 4.5 | 22.7 |
| 70-79 | 119.3 | 57.8 | 227.9 | 12.0 | 5.8 | 22.9 |
| 80+ | 290.4 | 180.7 | 1027.3 | 29.2 | 18.2 | 103.3 |
| All age groups | 187.3 | 59.9 | 259.9 | 3.1 | Ref | 4.3 |
| Male | AB | BC | ON | AB | BC | ON |
| <20 | 110.1 | 15.6 | 55.7 | 7.1 | Ref | 3.6 |
| 20-29 | 211.1 | 47.4 | 261.4 | 13.6 | 3.1 | 16.8 |
| 30-39 | 254.8 | 67.8 | 262.4 | 16.4 | 4.4 | 16.9 |
| 40-49 | 315.9 | 67.8 | 260.7 | 20.3 | 4.4 | 16.8 |
| 50-59 | 216.3 | 71.2 | 251.8 | 13.9 | 4.6 | 16.2 |
| 60-69 | 139.1 | 59.6 | 249.9 | 8.9 | 3.8 | 16.1 |
| 70-79 | 112.9 | 70.4 | 264.3 | 7.3 | 4.5 | 17.0 |
| 80+ | 242.5 | 149.5 | 719.3 | 15.6 | 9.6 | 46.2 |
| All age groups | 196.2 | 57.2 | 229.4 | 3.4 | Ref | 4.0 |

AB: Alberta; BC: British Columbia; ON: Ontario.

**Table S4.** Cumulative number of cases and cumulative incidence rate by age and gender in Alberta (N=8,389), as of July 5, 2020.

| Age group<br>(year) | Cases |  |  |  |  | Population<br>(2019) |  | Cases per<br>100,000 |  |
| --- | --- | --- | --- | --- | --- | --- | --- | --- | --- |
|  | F | M | Unk | % female<br>by age<br>group | Total by<br>age group<br>(%) | F | M | F | M |
| 0-4 | 121 | 123 | 0 | 49.6 | 244 (2.9) | 134487 | 139868 | 90.0 | 87.9 |
| <1 | 28 | 27 | 0 | 50.9 | 55 (0.7) | No data | No data | n/a | n/a |
| 1-4 | 93 | 96 | 0 | 49.2 | 189 (2.3) | No data | No data | n/a | n/a |
| 5-9 | 116 | 120 | 1 | 48.9 | 237 (2.8) | 135170 | 141947 | 85.8 | 84.5 |
| 10-19 | 404 | 362 | 1 | 52.7 | 767 (9.1) | 131345 | 137050 | 307.6 | 264.1 |
| 20-29 | 604 | 655 | 1 | 47.9 | 1260 (15.0) | 289835 | 310327 | 208.4 | 211.1 |
| 30-39 | 789 | 914 | 0 | 46.3 | 1703 (20.3) | 349731 | 358741 | 225.6 | 254.8 |
| 40-49 | 798 | 955 | 0 | 45.5 | 1753 (20.9) | 293475 | 302344 | 271.9 | 315.9 |
| 50-59 | 552 | 604 | 1 | 47.7 | 1157 (13.8) | 274579 | 279245 | 201.0 | 216.3 |
| 60-69 | 296 | 316 | 1 | 48.3 | 613 (7.3) | 229311 | 227188 | 129.1 | 139.1 |
| 70-79 | 154 | 132 | 0 | 53.8 | 286 (3.4) | 129090 | 116890 | 119.3 | 112.9 |
| 80+ | 234 | 134 | 0 | 63.6 | 368 (4.4) | 80569 | 55247 | 290.4 | 242.5 |
| Unk | 0 | 0 | 1 | 0 | 1 (0.01) | n/a | n/a | n/a | n/a |
| All age<br>groups | 4068 | 4315 | 6 | 48.5 | 8389 (100) | 2171882 | 2199434 | 187.3 | 196.2 |

CIR: cumulative incidence rate F: female; M: male; n/a: not applied; Unk: Unknown

**Table S5.**
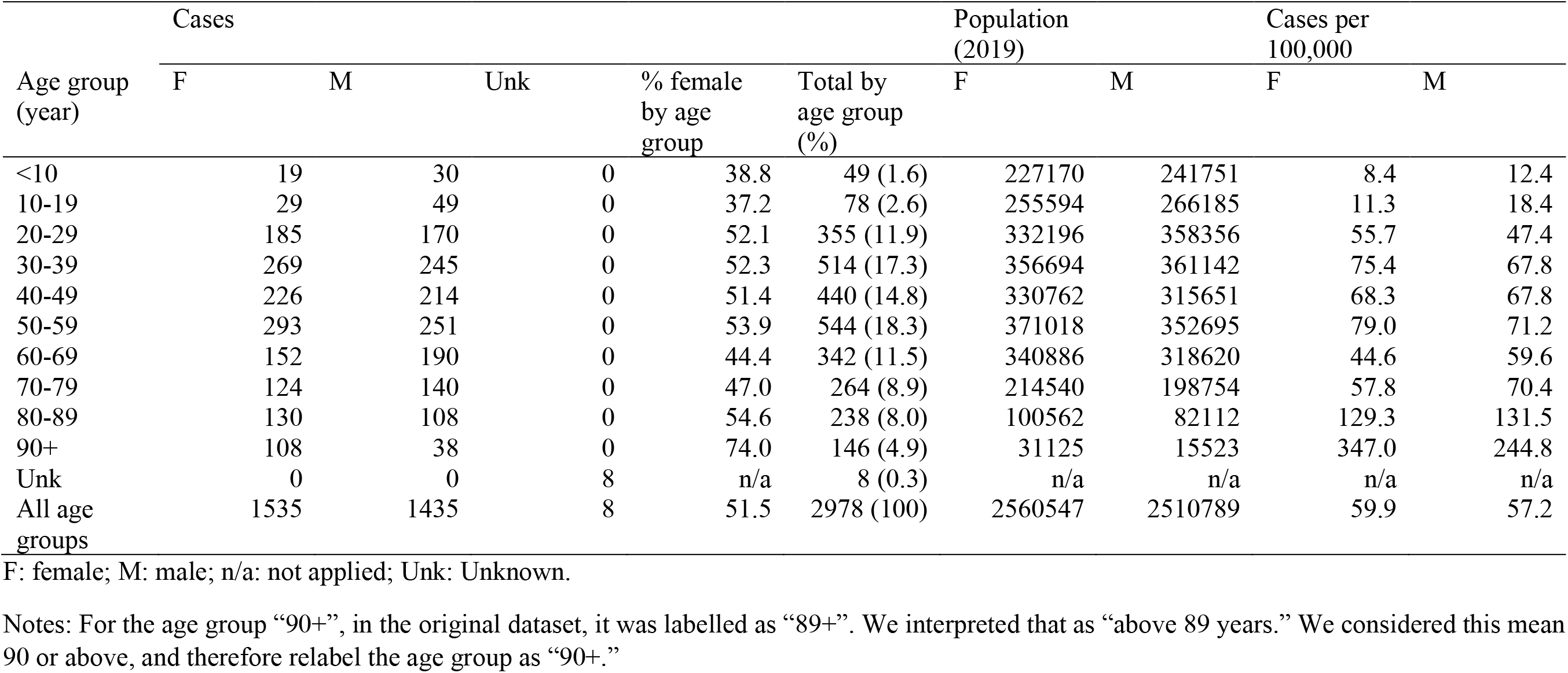
Cumulative number of cases and cumulative incidence rate by age and sex in British Columbia (N=2,978), as of July 6, 2020.

**Table S6.**
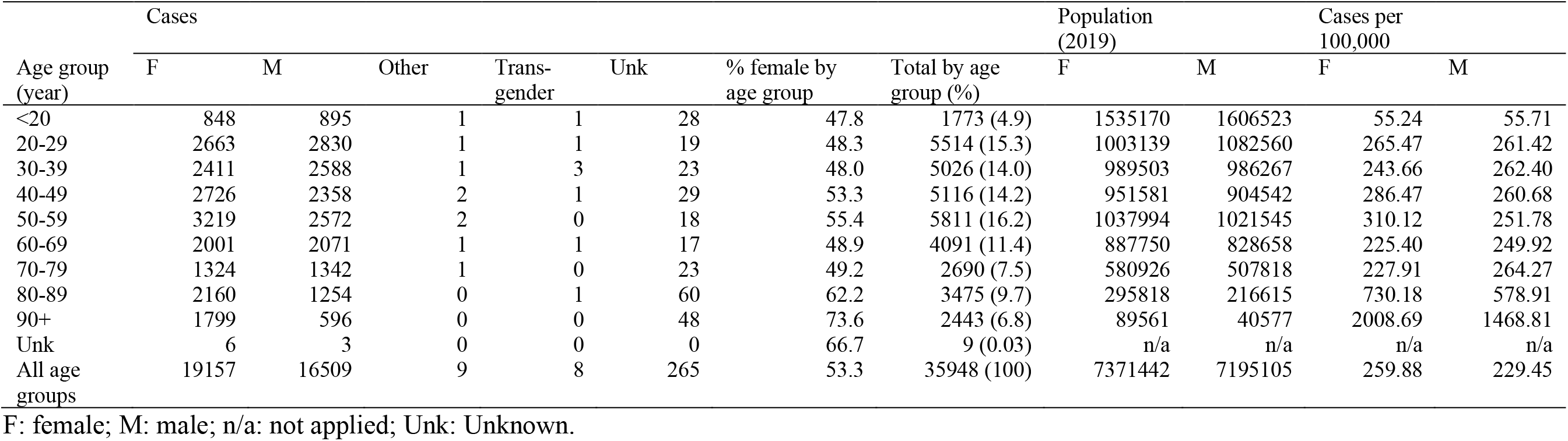
Cumulative number of cases and cumulative incidence rate by age and gender in Ontario, as of July 5, 2020.

**Table S7.** Cases by age, type and clinical outcome in Alberta, as of July 5, 2020.

| Age group (year) | Confirmed |  |  |  |  | Probable |  |  |  |  | All |  |  |  |  |
| --- | --- | --- | --- | --- | --- | --- | --- | --- | --- | --- | --- | --- | --- | --- | --- |
|  | A | D | R | T | D/(D+R) | A | D | R | T | D/(D+R) | A | D | R | T | D/(D+R) |
| <1 | 5 | 0 | 43 | 48 | 0% | 0 | 0 | 7 | 7 | 0% | 5 | 0 | 50 | 55 | 0% |
| 1-4 | 17 | 0 | 125 | 142 | 0% | 1 | 0 | 46 | 47 | 0% | 18 | 0 | 171 | 189 | 0% |
| 5-9 | 24 | 0 | 169 | 193 | 0% | 1 | 0 | 43 | 44 | 0% | 25 | 0 | 212 | 237 | 0% |
| 10-19 | 66 | 0 | 598 | 664 | 0% | 2 | 0 | 101 | 103 | 0% | 68 | 0 | 699 | 767 | 0% |
| 20-29 | 127 | 1 | 1023 | 1151 | 0.1% | 3 | 0 | 106 | 109 | 0% | 130 | 1 | 1129 | 1260 | 0.1% |
| 30-39 | 116 | 1 | 1471 | 1588 | 0.1% | 4 | 0 | 111 | 115 | 0% | 120 | 1 | 1582 | 1703 | 0.1% |
| 40-49 | 76 | 1 | 1559 | 1636 | 0.1% | 3 | 0 | 114 | 117 | 0% | 79 | 1 | 1673 | 1753 | 0.1% |
| 50-59 | 59 | 2 | 1009 | 1070 | 0.2% | 0 | 0 | 87 | 87 | 0% | 59 | 2 | 1096 | 1157 | 0.2% |
| 60-69 | 43 | 10 | 496 | 549 | 2.0% | 2 | 0 | 62 | 64 | 0% | 45 | 10 | 558 | 613 | 1.8% |
| 70-79 | 29 | 31 | 214 | 274 | 12.7% | 1 | 0 | 11 | 12 | 0% | 30 | 31 | 225 | 286 | 12.1% |
| 80+ | 28 | 108 | 226 | 362 | 32.3% | 0 | 1 | 5 | 6 | 16.7% | 28 | 109 | 231 | 368 | 32.1% |
| Unknown | 0 | 0 | 1 | 1 | 0% | 0 | 0 | 0 | 0 | n/a | 0 | 0 | 1 | 1 | 0% |
| All age groups | 590 | 154 | 6934 | 7678 | 2.2% | 17 | 1 | 693 | 711 | 0.1% | 607 | 155 | 7627 | 8389 | 2.0% |

A: Active; R: Recovered; D: Died; T: Total; D/(D+R): Died/(Died+Recovered); n/a: not applied

**Table S8.** Cases by age and clinical outcome in Ontario, as of July 5, 2020.

| Age group (years) | A | D | R | T | D/(D+R) |
| --- | --- | --- | --- | --- | --- |
| <20 | 167 | 1 | 1605 | 1773 | 0.1% |
| 20-29 | 452 | 4 | 5058 | 5514 | 0.1% |
| 30-39 | 321 | 7 | 4698 | 5026 | 0.1% |
| 40-49 | 240 | 21 | 4855 | 5116 | 0.4% |
| 50-59 | 265 | 87 | 5459 | 5811 | 1.6% |
| 60-69 | 177 | 236 | 3678 | 4091 | 6.0% |
| 70-79 | 92 | 481 | 2117 | 2690 | 18.5% |
| 80-89 | 71 | 976 | 2428 | 3475 | 28.7% |
| 90+ | 47 | 876 | 1520 | 2443 | 36.6% |
| Unknown | 1 | 0 | 8 | 9 | 0% |

A: Active (“Not Resolved” in the dataset); D: Died (“Fatal” in the dataset); R: Recovered (“Resolved in the dataset); T: Total; D/(D+R):
Died/(Died+Recovered); n/a: not applied

**Table S9.** Cases by age and classification reported in British Columbia, as of July 6, 2020.

| Age group (year) | Lab-diagnosed (confirmed) (% by age group) | Epi-linked (probable) (% by age group) | Total |
| --- | --- | --- | --- |
| <10 | 48 (98.0) | 1 (2.0) | 49 (100) |
| 10-19 | 78 (100) | 0 (0.0) | 78 (100) |
| 20-29 | 354 (99.7) | 1 (0.3) | 355 (100) |
| 30-39 | 512 (99.6) | 2 (0.4) | 514 (100) |
| 40-49 | 439 (99.8) | 1 (0.2) | 440 (100) |
| 50-59 | 543 (99.8) | 1 (0.2) | 544 (100) |
| 60-69 | 341 (99.7) | 1 (0.2) | 342 (100) |
| 70-79 | 264 (100) | 0 (0.0) | 264 (100) |
| 80-89 | 237 (99.6) | 1 (0.4) | 238 (100) |
| 90+ | 146 (100) | 0 (0.0) | 146 (100) |
| Unknown | 8 (100) | 0 (0.0) | 8 (100) |
| All age groups | 2970 (99.7) | 8 (0.3) | 2978 (100) |

**Table S10.** Among 2391 Ontario cases whose specimen collection (specimen date) happened prior to symptom onset (accurate episode date), as of July 5, 2020.

|  | Time difference between accurate episode date and specimen collection date | Time difference between accurate episode date and test report date | Time difference between accurate episode date and case report date |
| --- | --- | --- | --- |
| Minimum | -89 | -33 | 0 |
| 1<sup>st</sup> quartile | -3 | 0 | 0 |
| Median | -2 | 0 | 0 |
| Mean | -2.716 | 1.028 | 1.127 |
| 3<sup>rd</sup> quartile | -1 | 1 | 1 |
| Maximum | -1 | 55 | 38 |
| NA | n=0 | n=8 * | n=0 |

\*All 8 case-patients whose test reported date is NA, developed symptoms after the date of specimen collection. Perhaps their tests were not positive. But they were reported as cases on or after the date of their symptom onset.

**Table S11.**
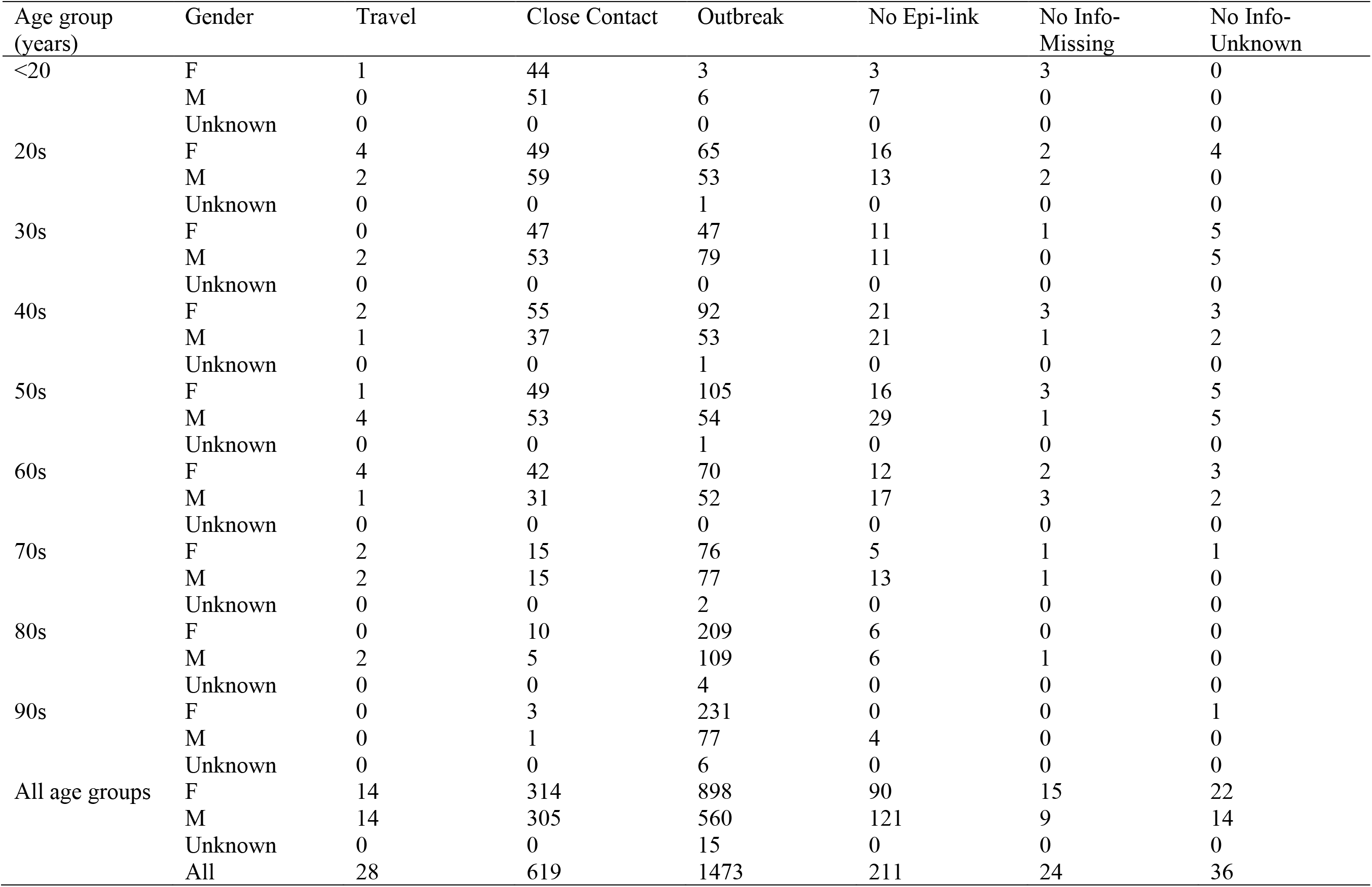
Among 2391 Ontario cases whose specimen collection (specimen date) happened prior to symptom onset (accurate episode date), as of July 5, 2020.

| Age group<br>(years) | Gender | Travel | Close Contact | Outbreak | No Epi-link | No Info-Missing | No Info-Unknown |
| --- | --- | --- | --- | --- | --- | --- | --- |
| <20 | F | 1 | 44 | 3 | 3 | 3 | 0 |
|  | M | 0 | 51 | 6 | 7 | 0 | 0 |
|  | Unknown | 0 | 0 | 0 | 0 | 0 | 0 |
| 20s | F | 4 | 49 | 65 | 16 | 2 | 4 |
|  | M | 2 | 59 | 53 | 13 | 2 | 0 |
|  | Unknown | 0 | 0 | 1 | 0 | 0 | 0 |
| 30s | F | 0 | 47 | 47 | 11 | 1 | 5 |
|  | M | 2 | 53 | 79 | 11 | 0 | 5 |
|  | Unknown | 0 | 0 | 0 | 0 | 0 | 0 |
| 40s | F | 2 | 55 | 92 | 21 | 3 | 3 |
|  | M | 1 | 37 | 53 | 21 | 1 | 2 |
|  | Unknown | 0 | 0 | 1 | 0 | 0 | 0 |
| 50s | F | 1 | 49 | 105 | 16 | 3 | 5 |
|  | M | 4 | 53 | 54 | 29 | 1 | 5 |
|  | Unknown | 0 | 0 | 1 | 0 | 0 | 0 |
| 60s | F | 4 | 42 | 70 | 12 | 2 | 3 |
|  | M | 1 | 31 | 52 | 17 | 3 | 2 |
|  | Unknown | 0 | 0 | 0 | 0 | 0 | 0 |
| 70s | F | 2 | 15 | 76 | 5 | 1 | 1 |
|  | M | 2 | 15 | 77 | 13 | 1 | 0 |
|  | Unknown | 0 | 0 | 2 | 0 | 0 | 0 |
| 80s | F | 0 | 10 | 209 | 6 | 0 | 0 |
|  | M | 2 | 5 | 109 | 6 | 1 | 0 |
|  | Unknown | 0 | 0 | 4 | 0 | 0 | 0 |
| 90s | F | 0 | 3 | 231 | 0 | 0 | 1 |
|  | M | 0 | 1 | 77 | 4 | 0 | 0 |
|  | Unknown | 0 | 0 | 6 | 0 | 0 | 0 |
| All age groups | F | 14 | 314 | 898 | 90 | 15 | 22 |
|  | M | 14 | 305 | 560 | 121 | 9 | 14 |
|  | Unknown | 0 | 0 | 15 | 0 | 0 | 0 |
|  | All | 28 | 619 | 1473 | 211 | 24 | 36 |

## Supplementary Figure legends

**Figure S1.**
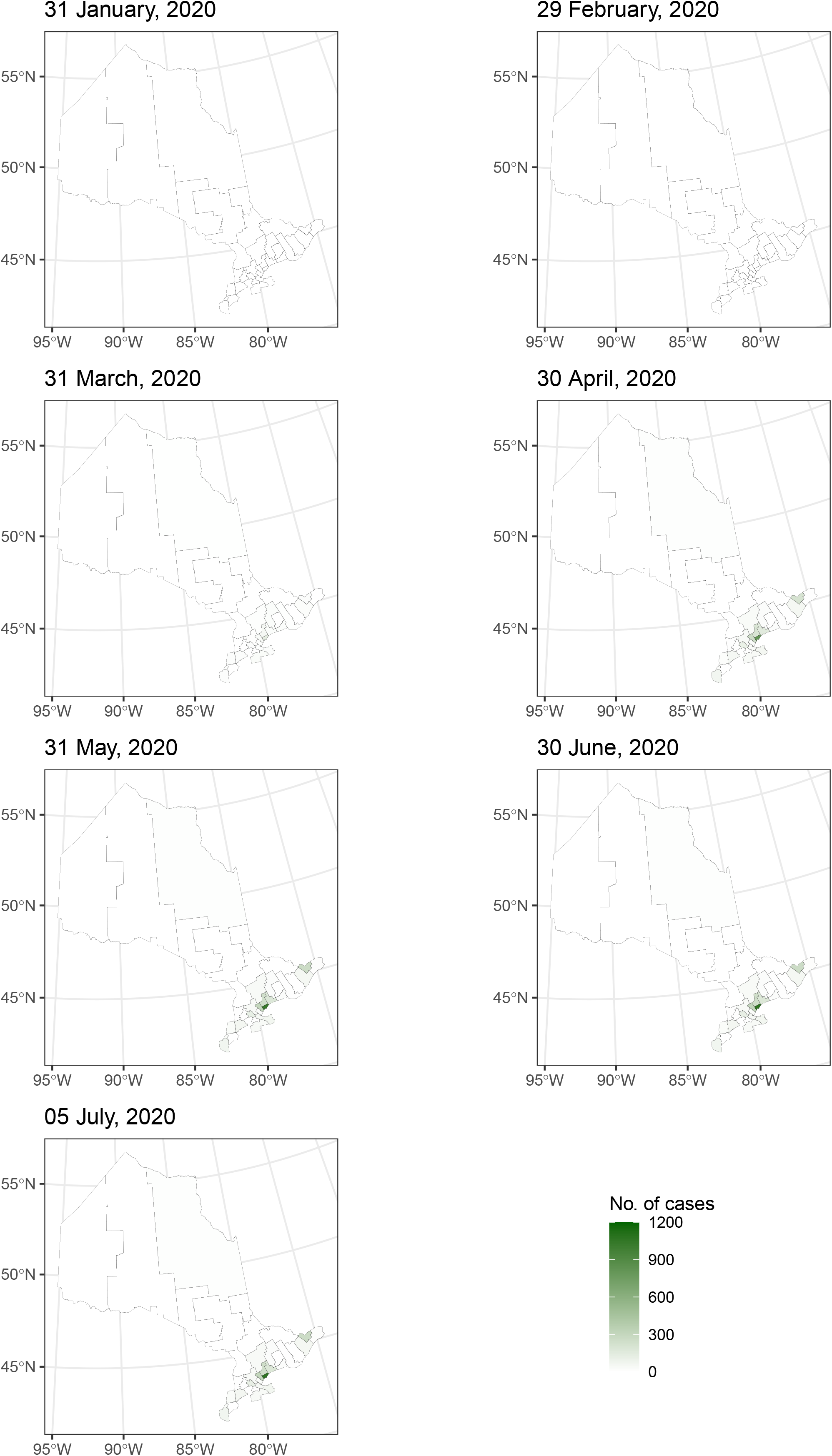
Map of cumulative number of deaths in Ontario by public health unit by the end of month and on July 6, 2020.

**Figure S2.**
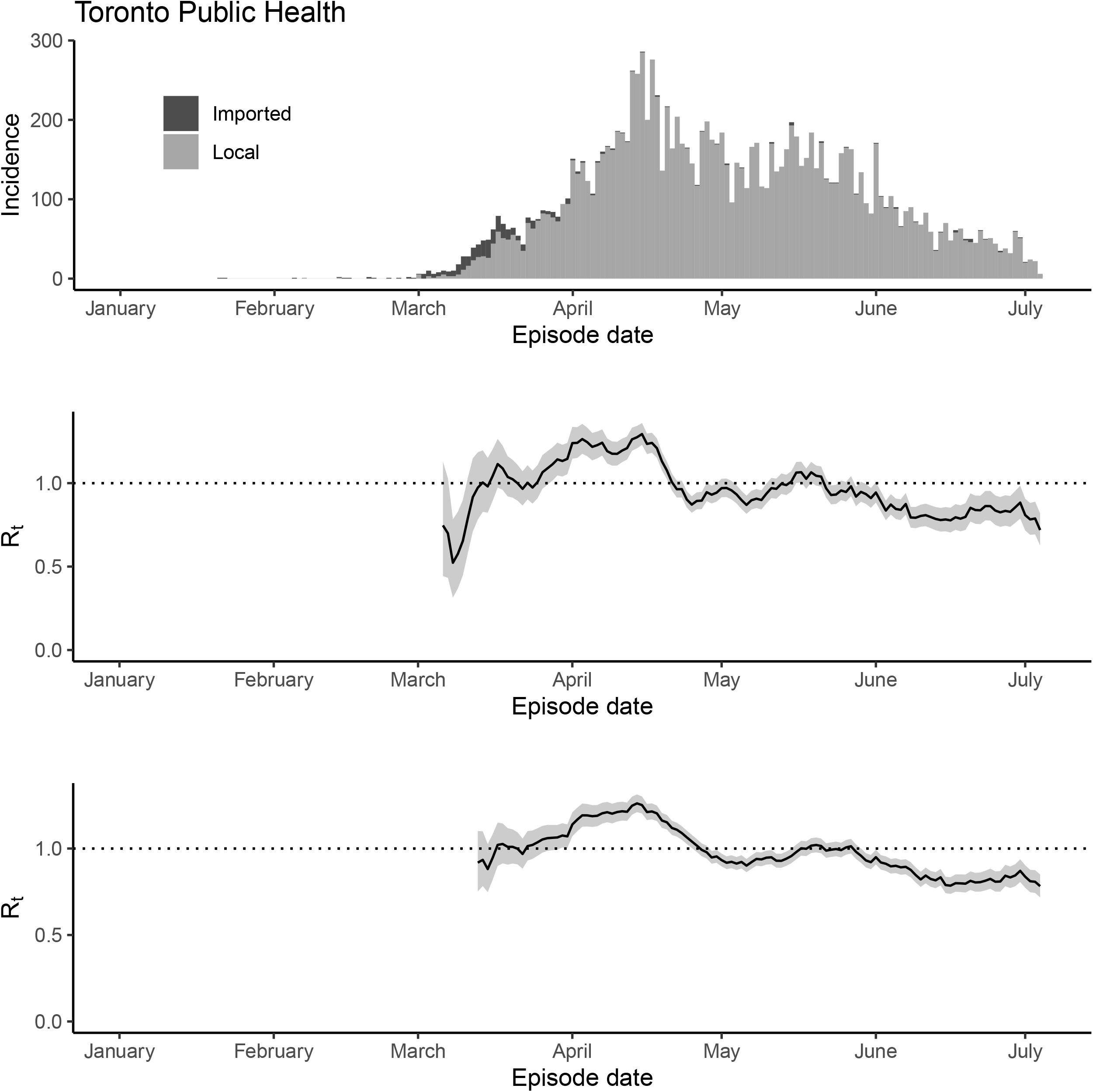
The epidemic trajectory of coronavirus disease 2019 in Toronto Public Health, Ontario, January 1-July 5, 2020: Upper panel: Daily number of new imported and local cases by date of symptom onset (aka accurate episode date) (upper panel); Rt with a 1-week window (middle panel); Rt with a 2-week window (lower panel).

**Figure S3.**
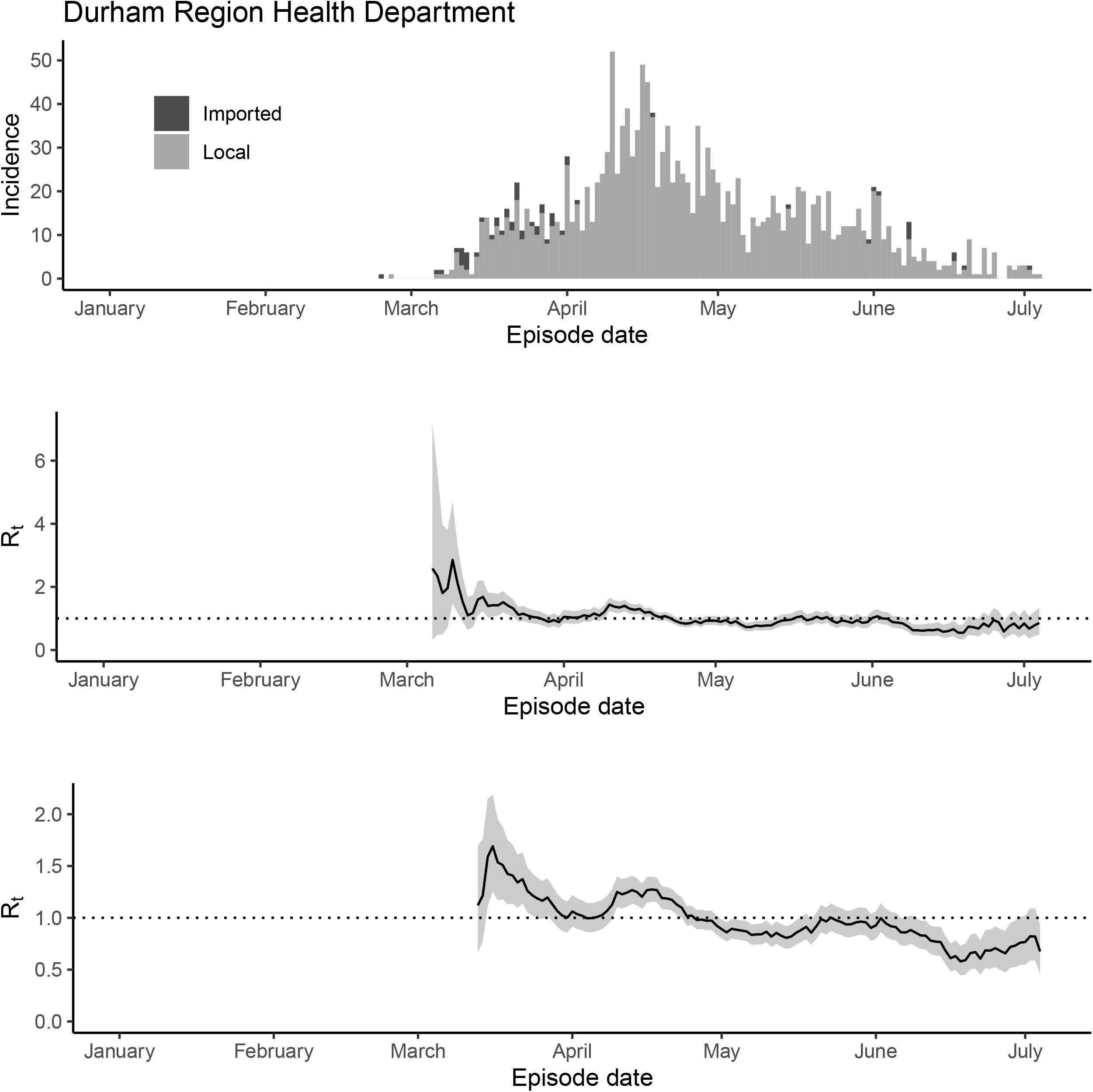
The epidemic trajectory of coronavirus disease 2019 in Durham Region Health Department, Ontario, January 1-July 5, 2020: Upper panel: Daily number of new imported and local cases by date of symptom onset (aka accurate episode date) (upper panel); Rt with a 1-week window (middle panel); Rt with a 2-week window (lower panel).

**Figure S4.**
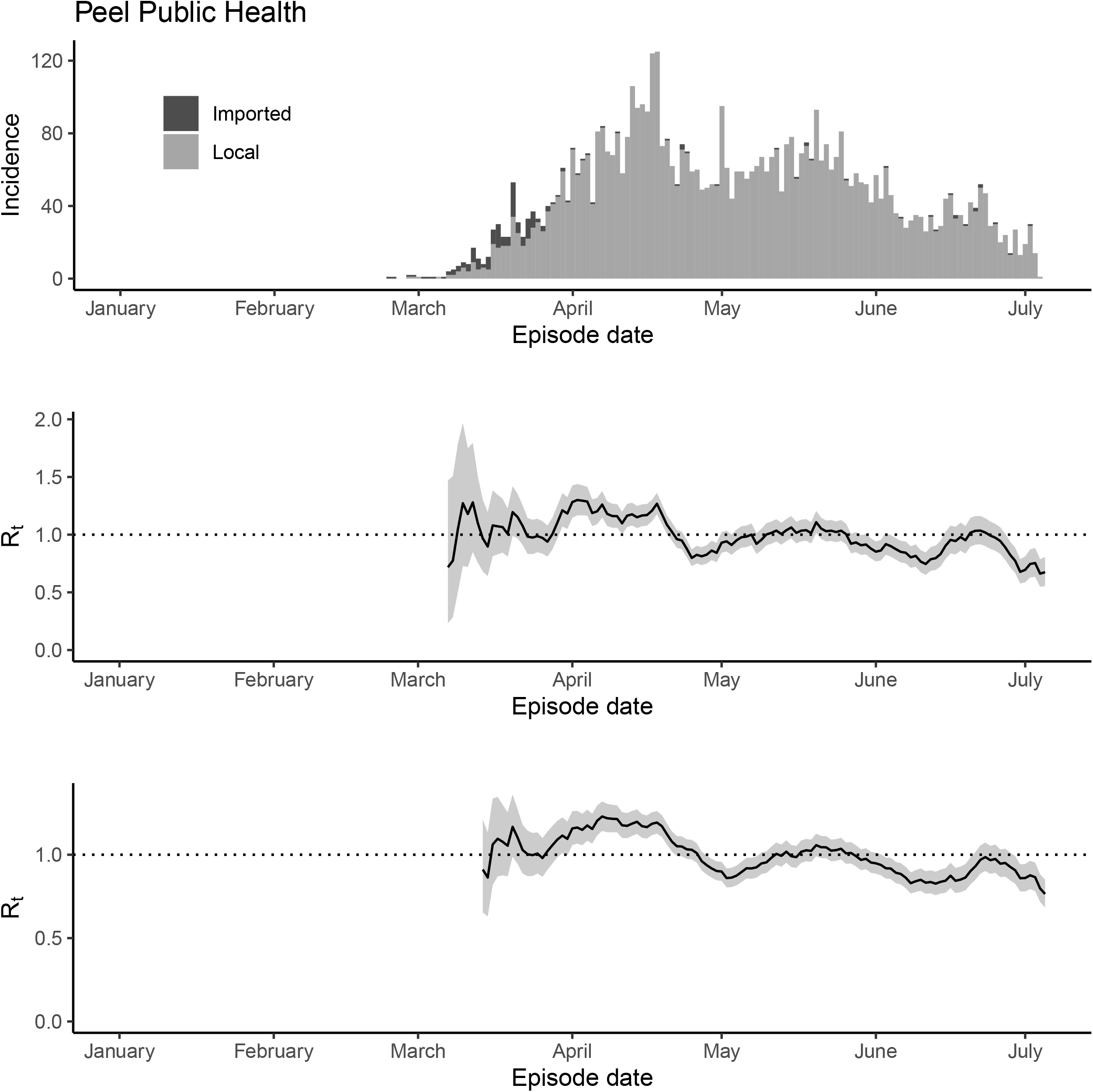
The epidemic trajectory of coronavirus disease 2019 in Peel Public Health, Ontario, January 1-July 5, 2020: Upper panel: Daily number of new imported and local cases by date of symptom onset (aka accurate episode date) (upper panel); Rt with a 1-week window (middle panel); Rt with a 2-week window (lower panel).

**Figure S5.**
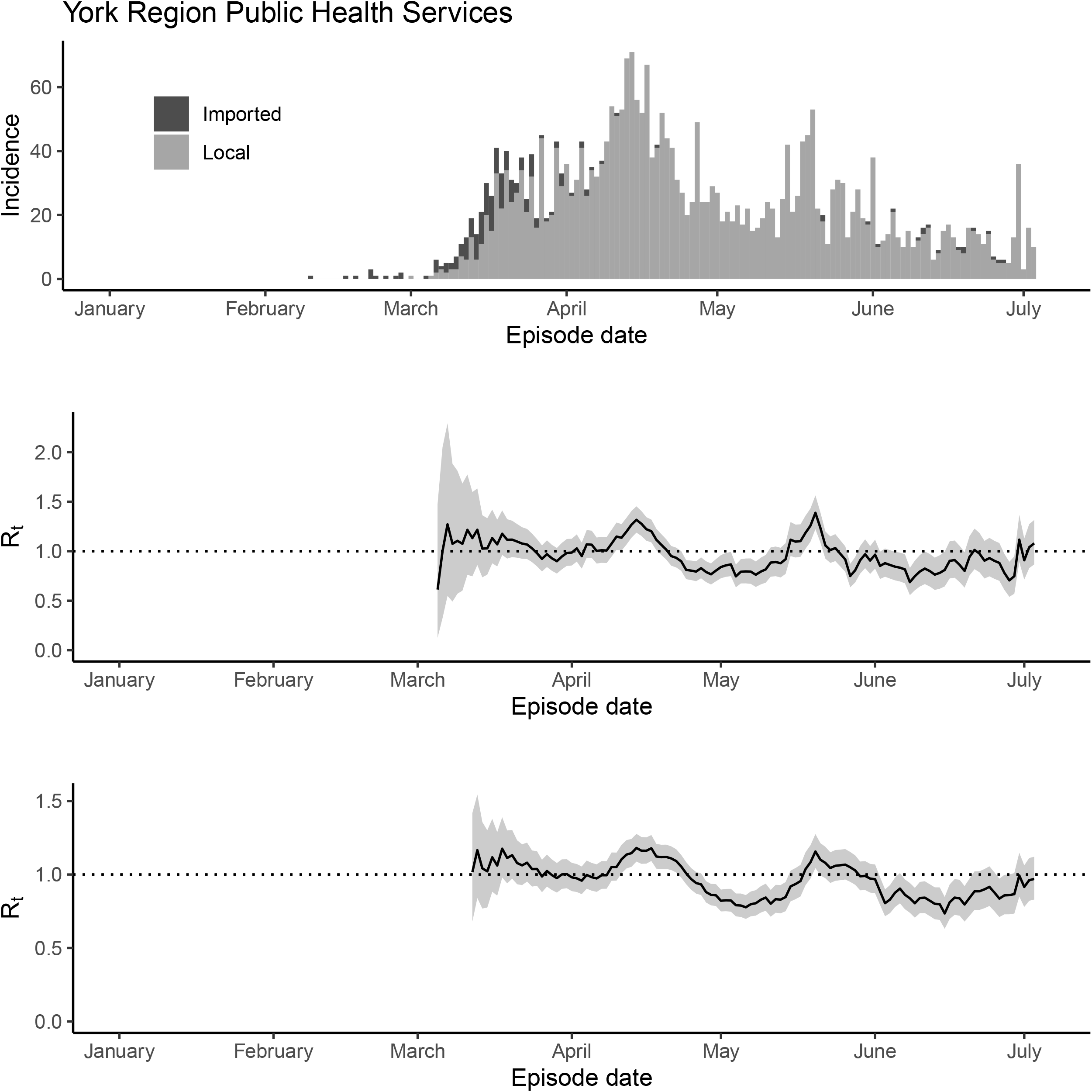
The epidemic trajectory of coronavirus disease 2019 in York Region Public Health Services, Ontario, January 1-July 5, 2020: Upper panel: Daily number of new imported and local cases by date of symptom onset (aka accurate episode date) (upper panel); Rt with a 1-week window (middle panel); Rt with a 2-week window (lower panel).

**Figure S6.**
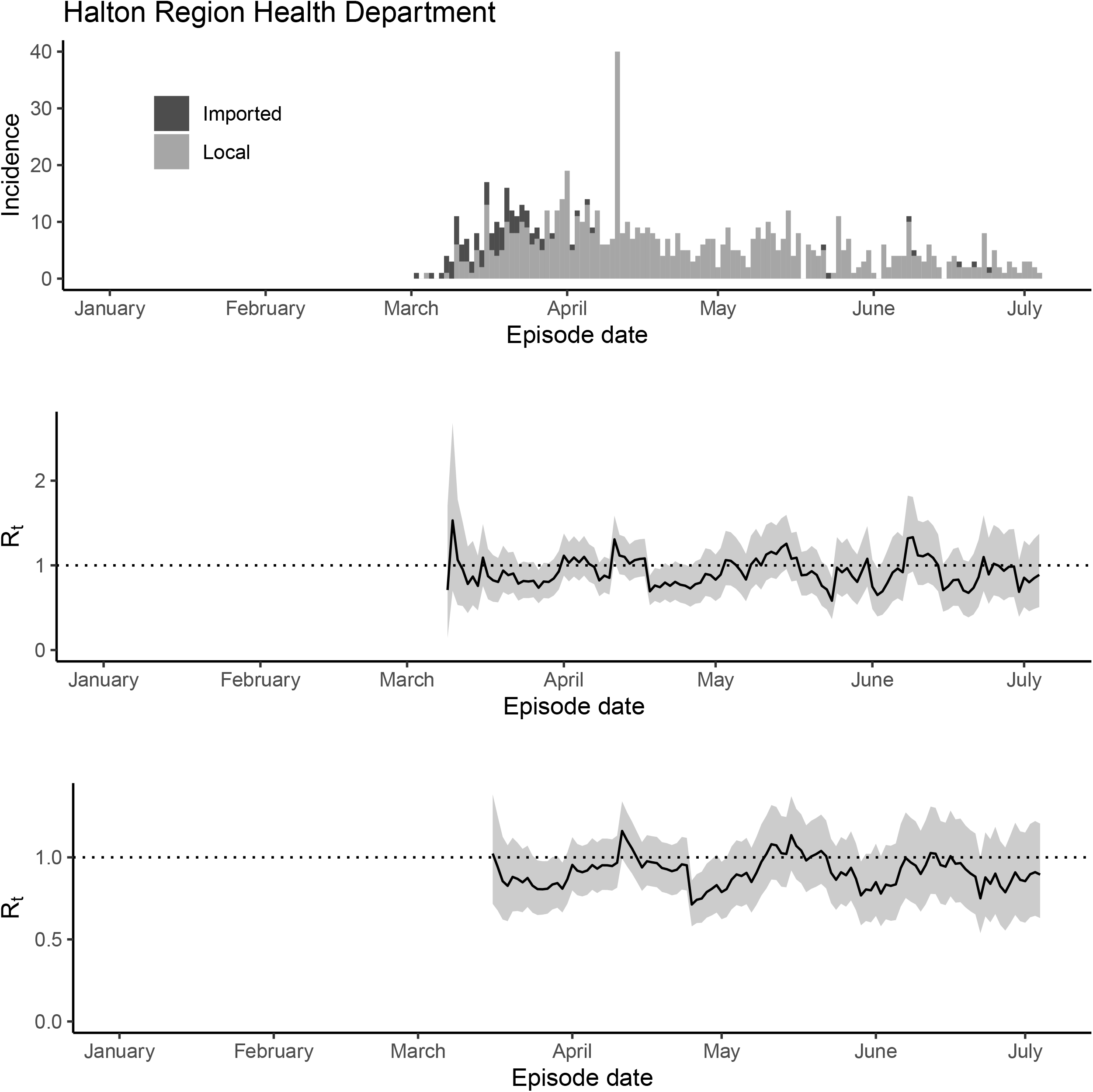
The epidemic trajectory of coronavirus disease 2019 in Halton Public Health Services, Ontario, January 1-July 5, 2020: Upper panel: Daily number of new imported and local cases by date of symptom onset (aka accurate episode date) (upper panel); Rt with a 1-week window (middle panel); Rt with a 2-week window (lower panel).

**Figure S7.**
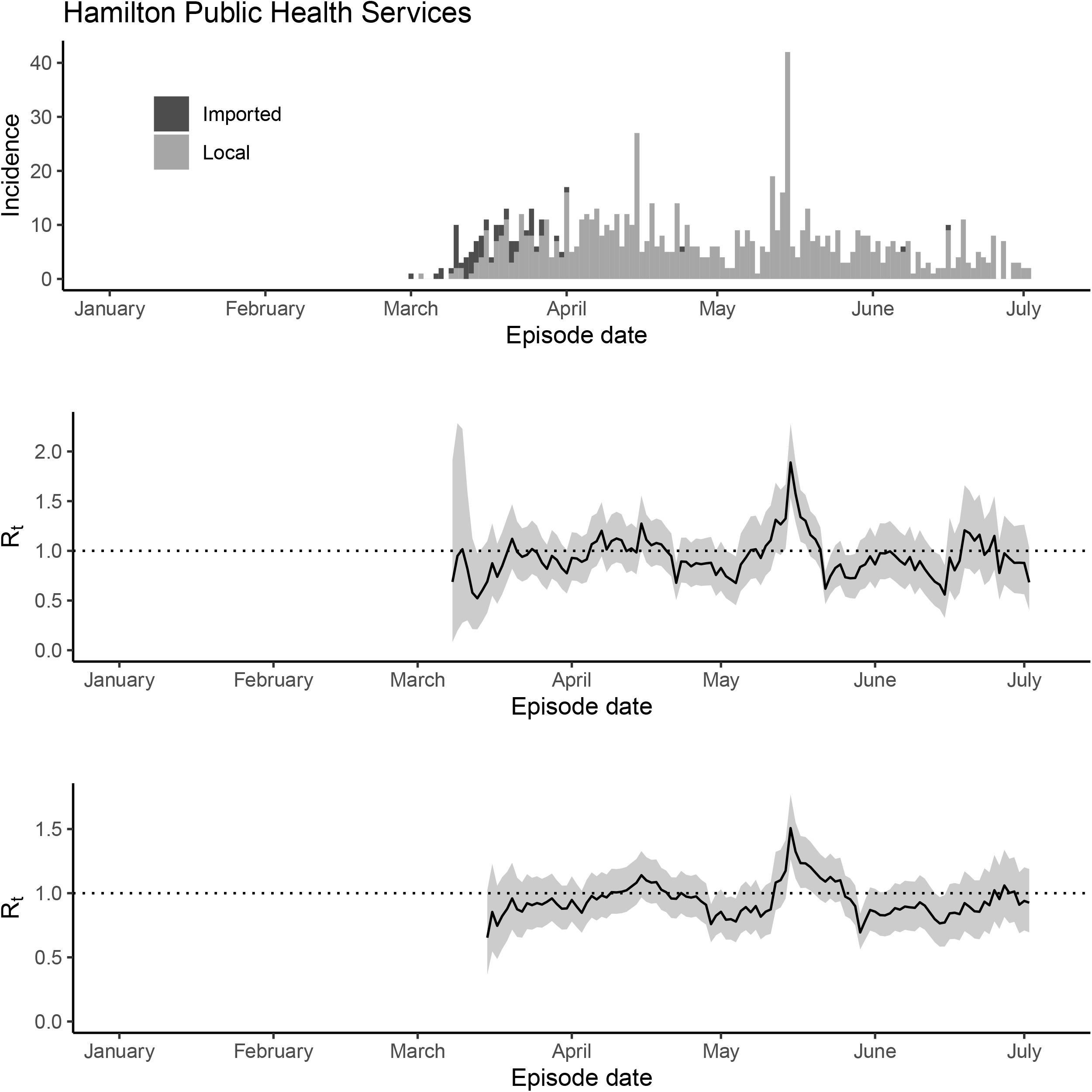
The epidemic trajectory of coronavirus disease 2019 in Hamilton Public Health Services, Ontario, January 1-July 5, 2020: Upper panel: Daily number of new imported and local cases by date of symptom onset (aka accurate episode date) (upper panel); Rt with a 1-week window (middle panel); Rt with a 2-week window (lower panel).

**Figure S8.**
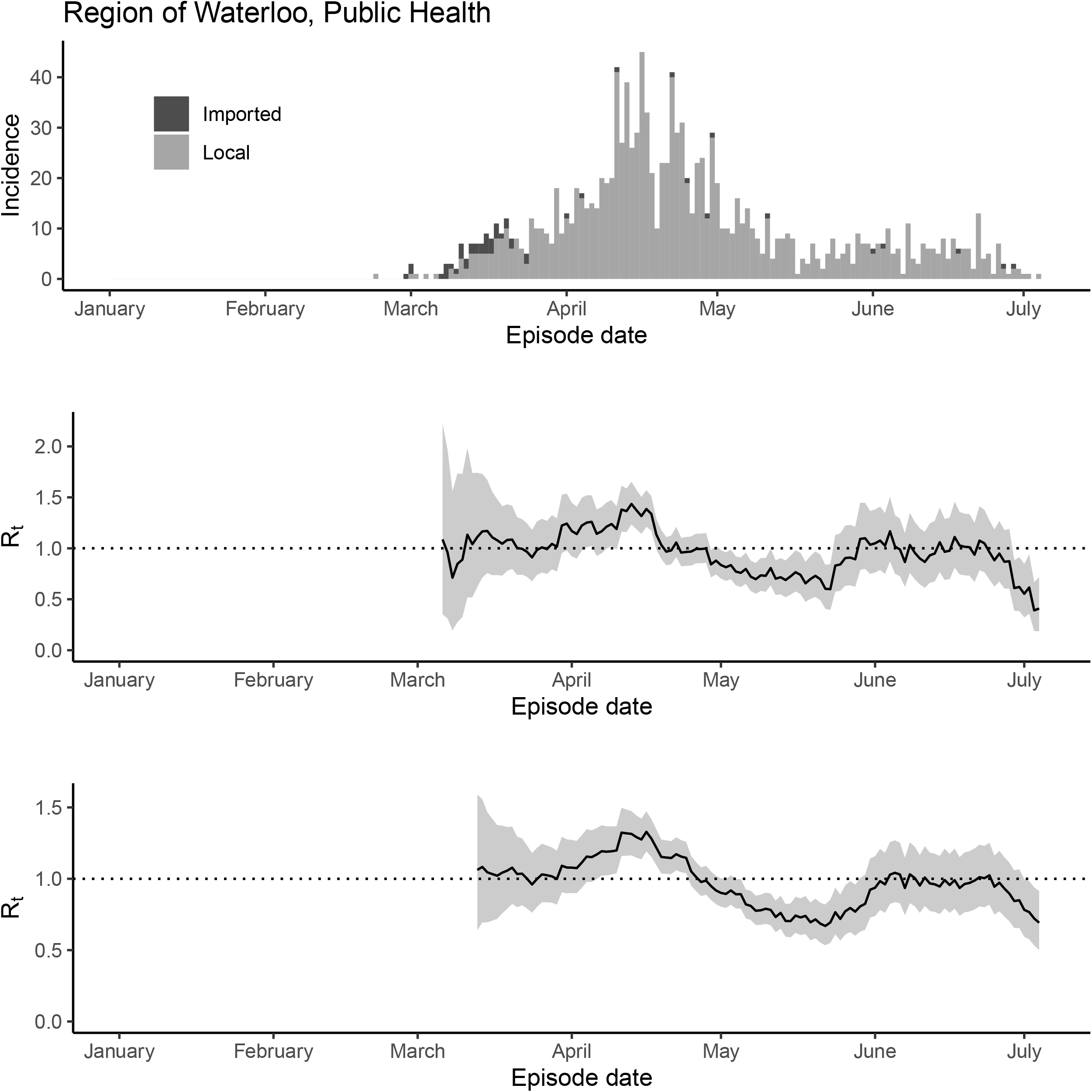
The epidemic trajectory of coronavirus disease 2019 in Region of Waterloo, Public Health Services, Ontario, January 1-July 5, 2020: Upper panel: Daily number of new imported and local cases by date of symptom onset (aka accurate episode date) (upper panel); Rt with a 1-week window (middle panel); Rt with a 2-week window (lower panel).

**Figure S9.**
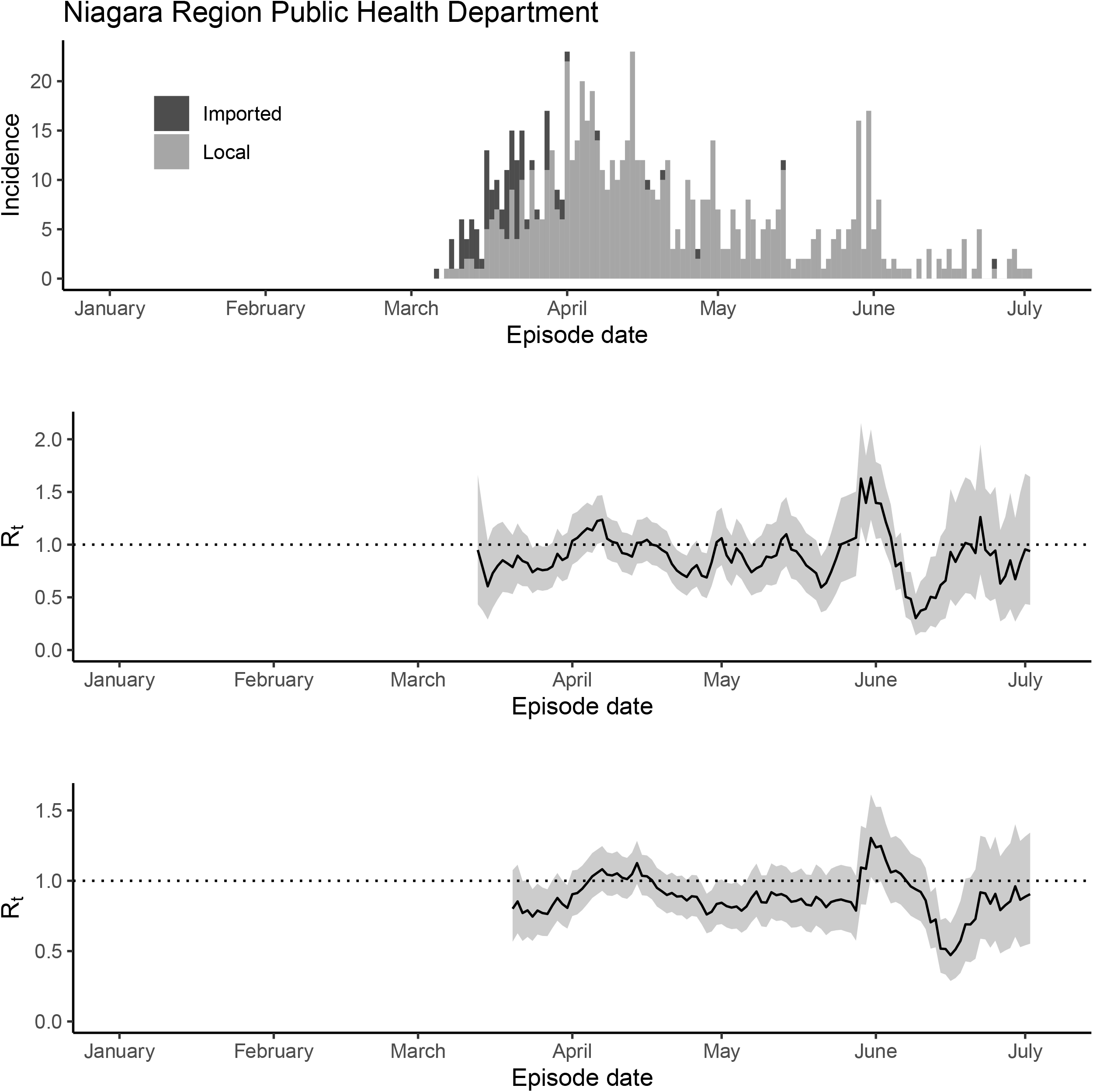
The epidemic trajectory of coronavirus disease 2019 in Niagara Region Public Health Department, Ontario, January 1-July 5, 2020: Upper panel: Daily number of new imported and local cases by date of symptom onset (aka accurate episode date) (upper panel); Rt with a 1-week window (middle panel); Rt with a 2-week window (lower panel). North Bay Parry Sound District Health Unit

**Figure S10.**
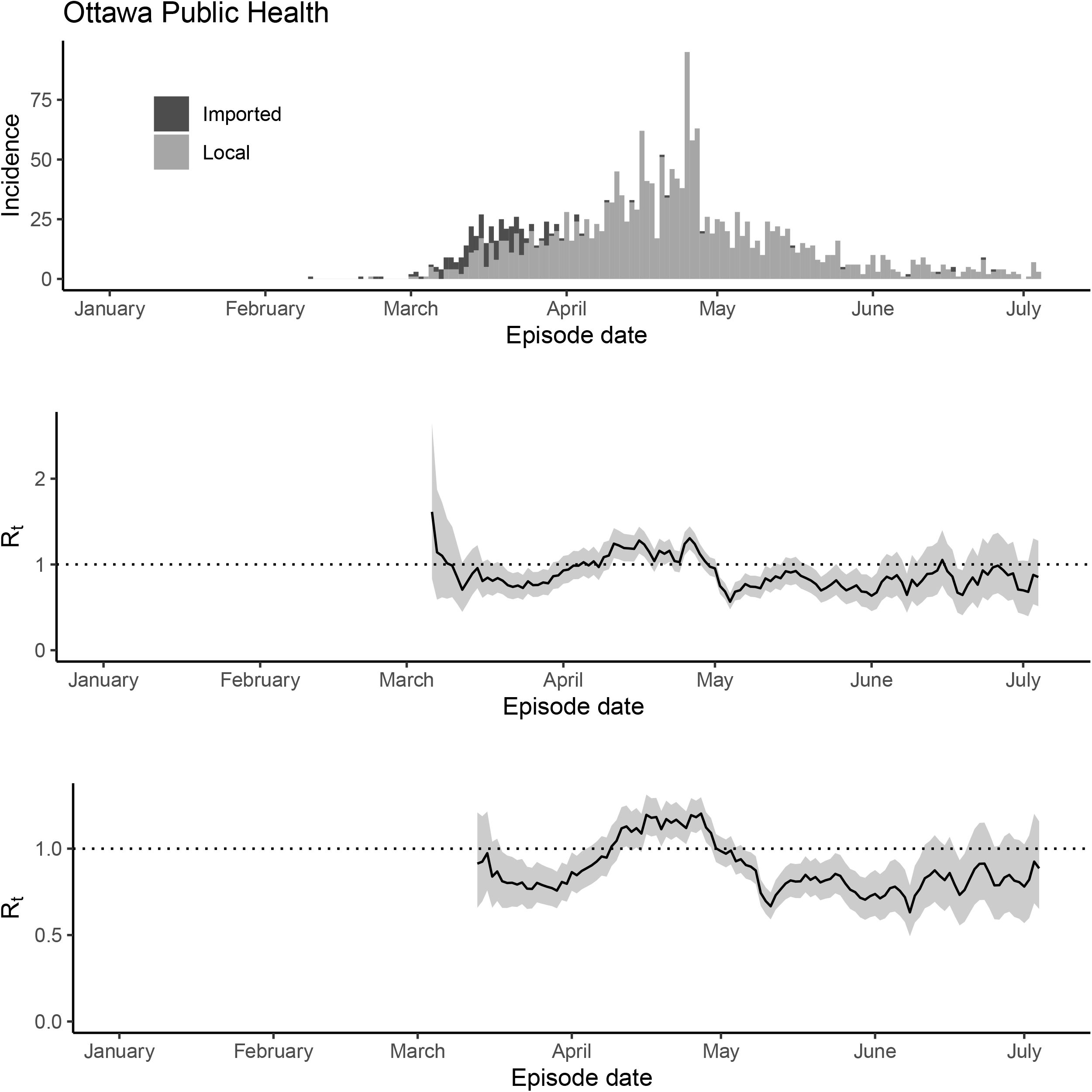
The epidemic trajectory of coronavirus disease 2019 in Ottawa Public Health Services, Ontario, January 1-July 5, 2020: Upper panel: Daily number of new imported and local cases by date of symptom onset (aka accurate episode date) (upper panel); Rt with a 1-week window (middle panel); Rt with a 2-week window (lower panel).

**Figure S11.**
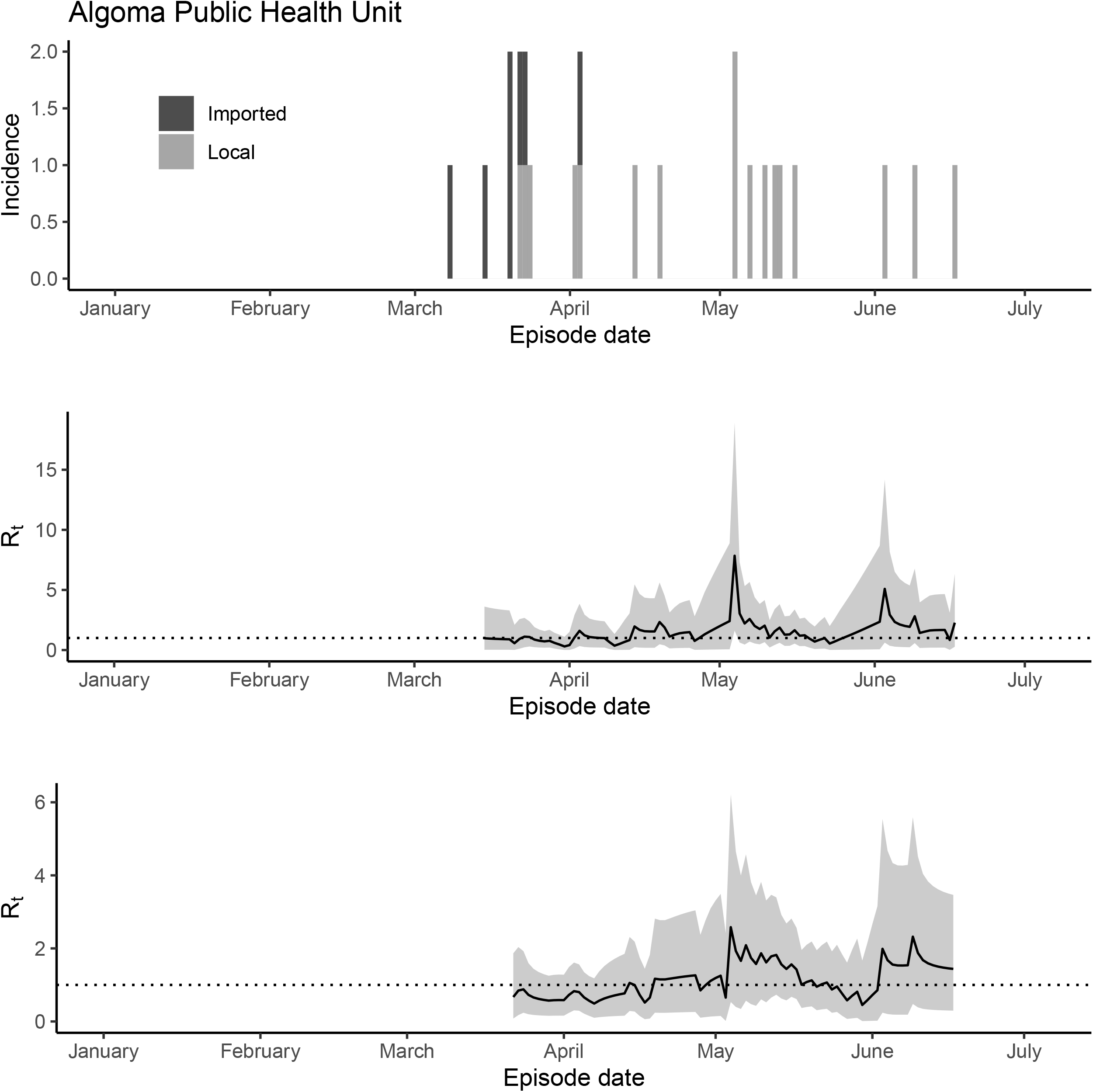
The epidemic trajectory of coronavirus disease 2019 in Algoma Public Health Unit, Ontario, January 1-July 5, 2020: Upper panel: Daily number of new imported and local cases by date of symptom onset (aka accurate episode date) (upper panel); Rt with a 1-week window (middle panel); Rt with a 2-week window (lower panel).

**Figure S12.**
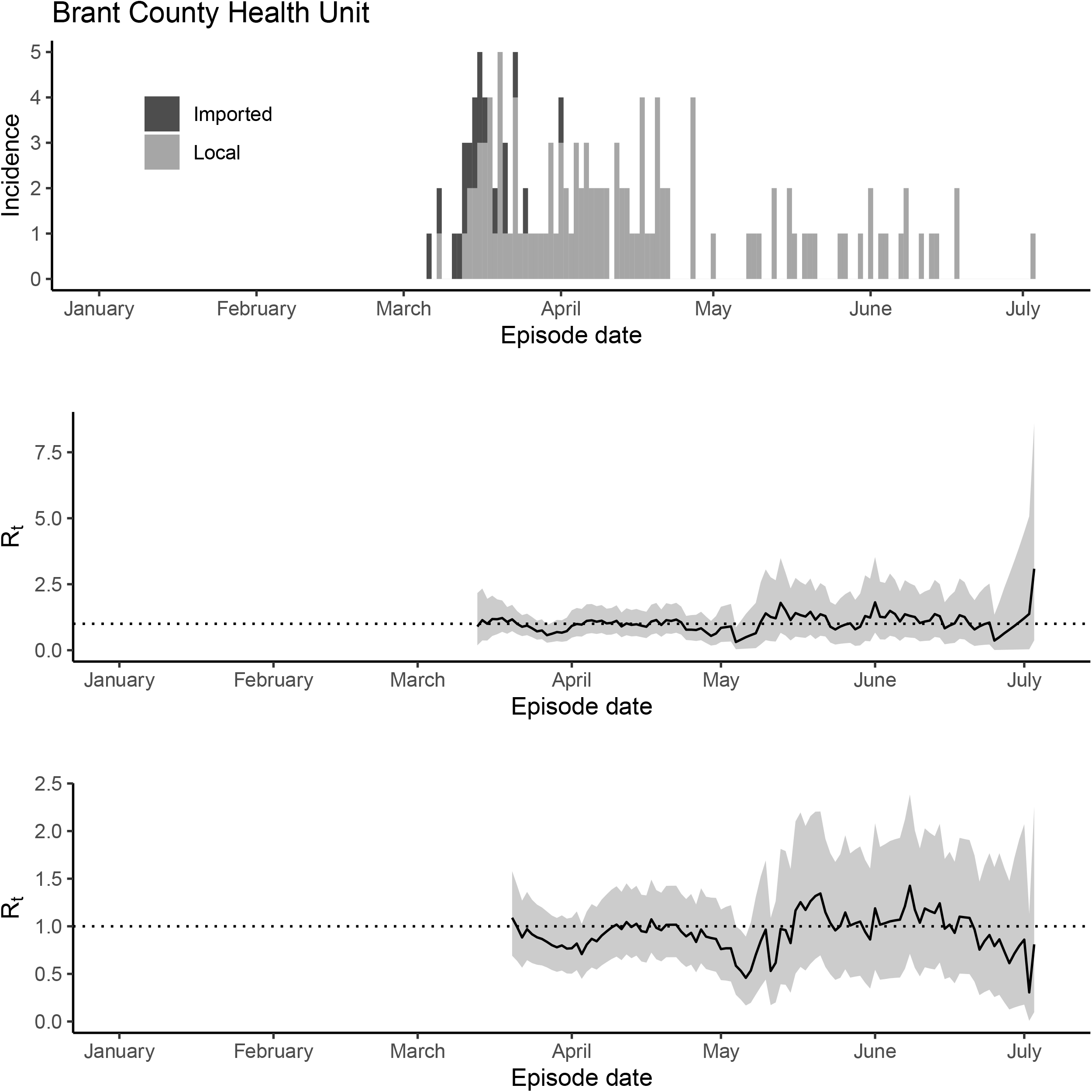
The epidemic trajectory of coronavirus disease 2019 in Brant County Health Unit, Ontario, January 1-July 5, 2020: Upper panel: Daily number of new imported and local cases by date of symptom onset (aka accurate episode date) (upper panel); Rt with a 1-week window (middle panel); Rt with a 2-week window (lower panel).

**Figure S13.**
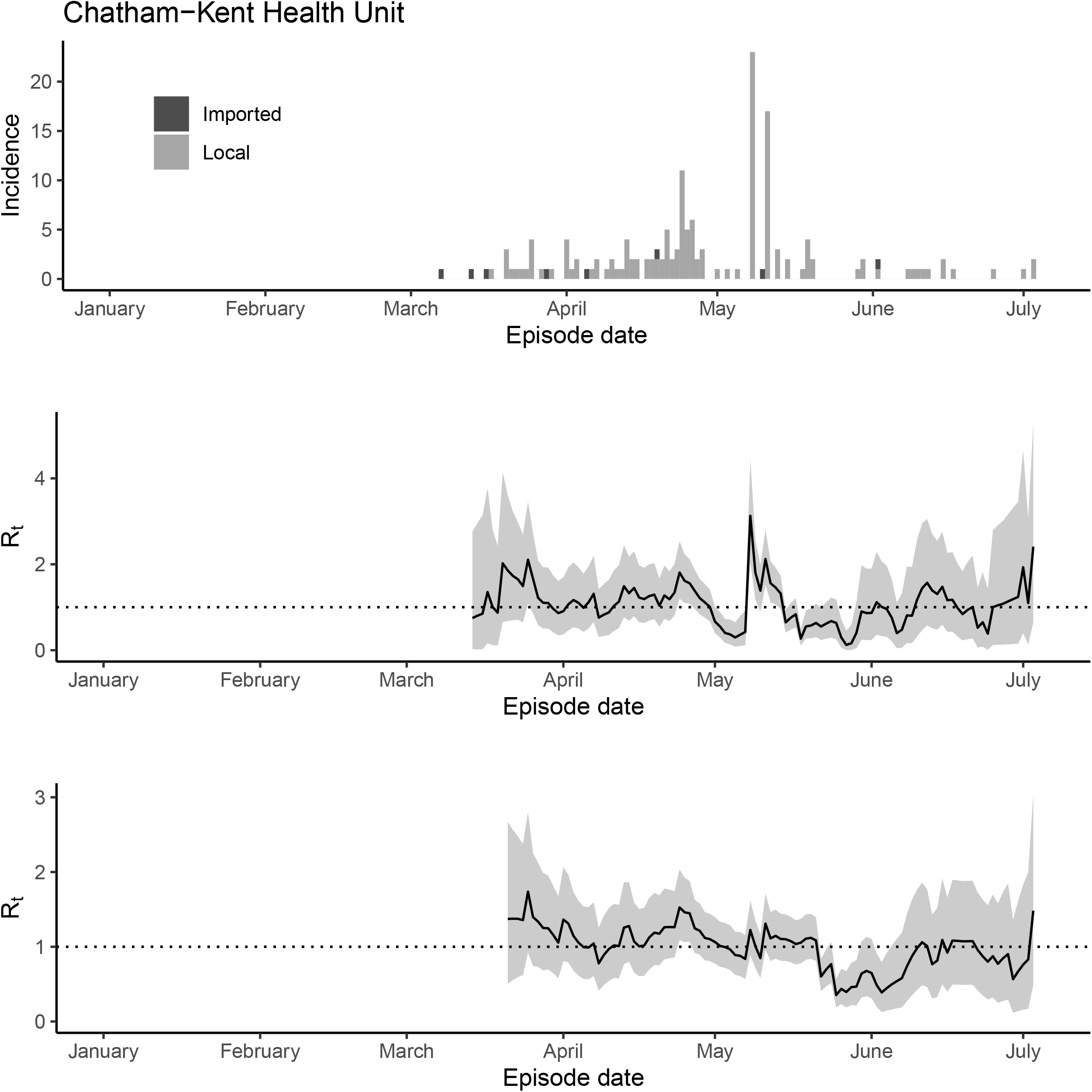
The epidemic trajectory of coronavirus disease 2019 in Chatham-Kent Health Unit, Ontario, January 1-July 5, 2020: Upper panel: Daily number of new imported and local cases by date of symptom onset (aka accurate episode date) (upper panel); Rt with a 1-week window (middle panel); Rt with a 2-week window (lower panel).

**Figure S14.**
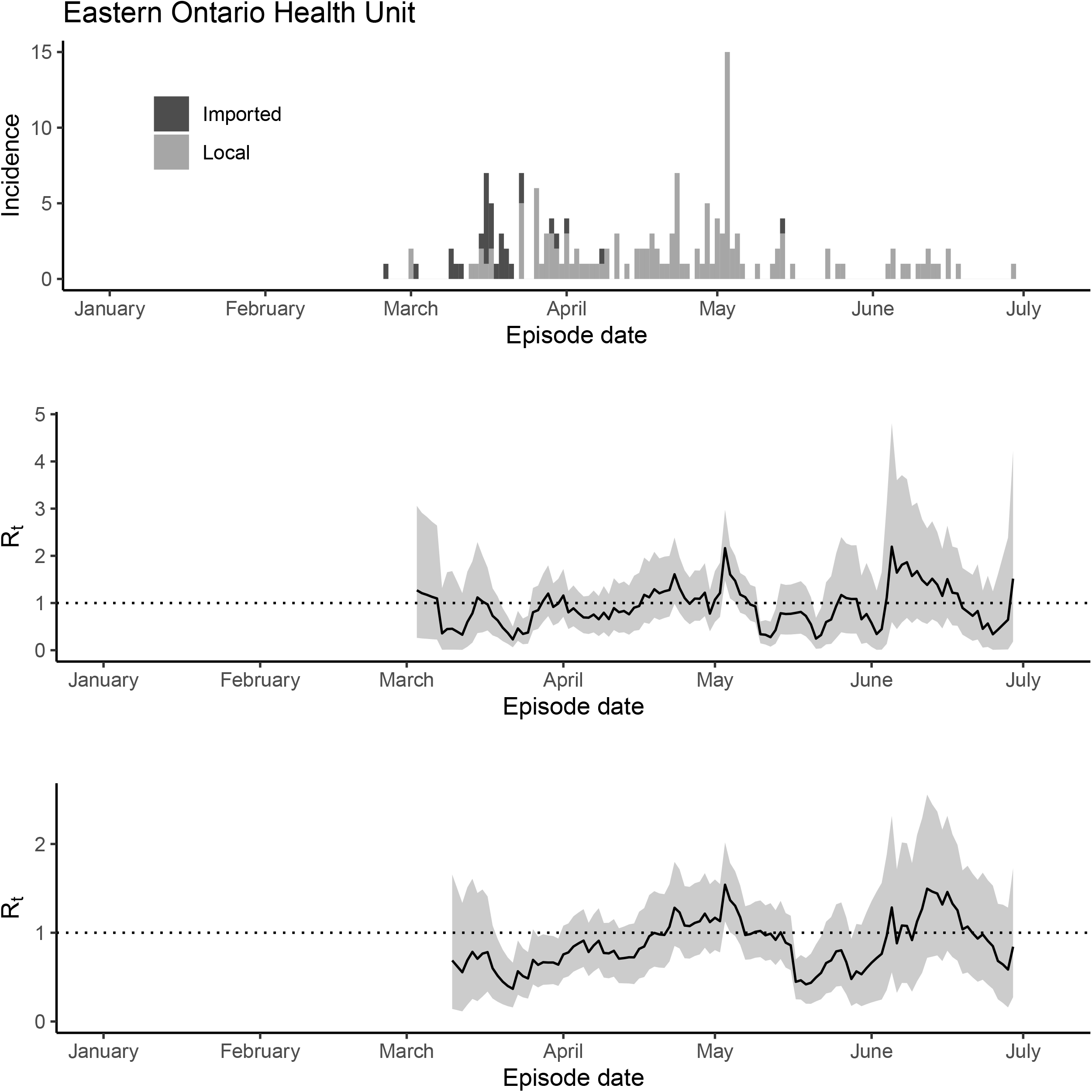
The epidemic trajectory of coronavirus disease 2019 in Eastern Ontario Health Unit, Ontario, January 1-July 5, 2020: Upper panel: Daily number of new imported and local cases by date of symptom onset (aka accurate episode date) (upper panel); Rt with a 1-week window (middle panel); Rt with a 2-week window (lower panel).

**Figure S15.**
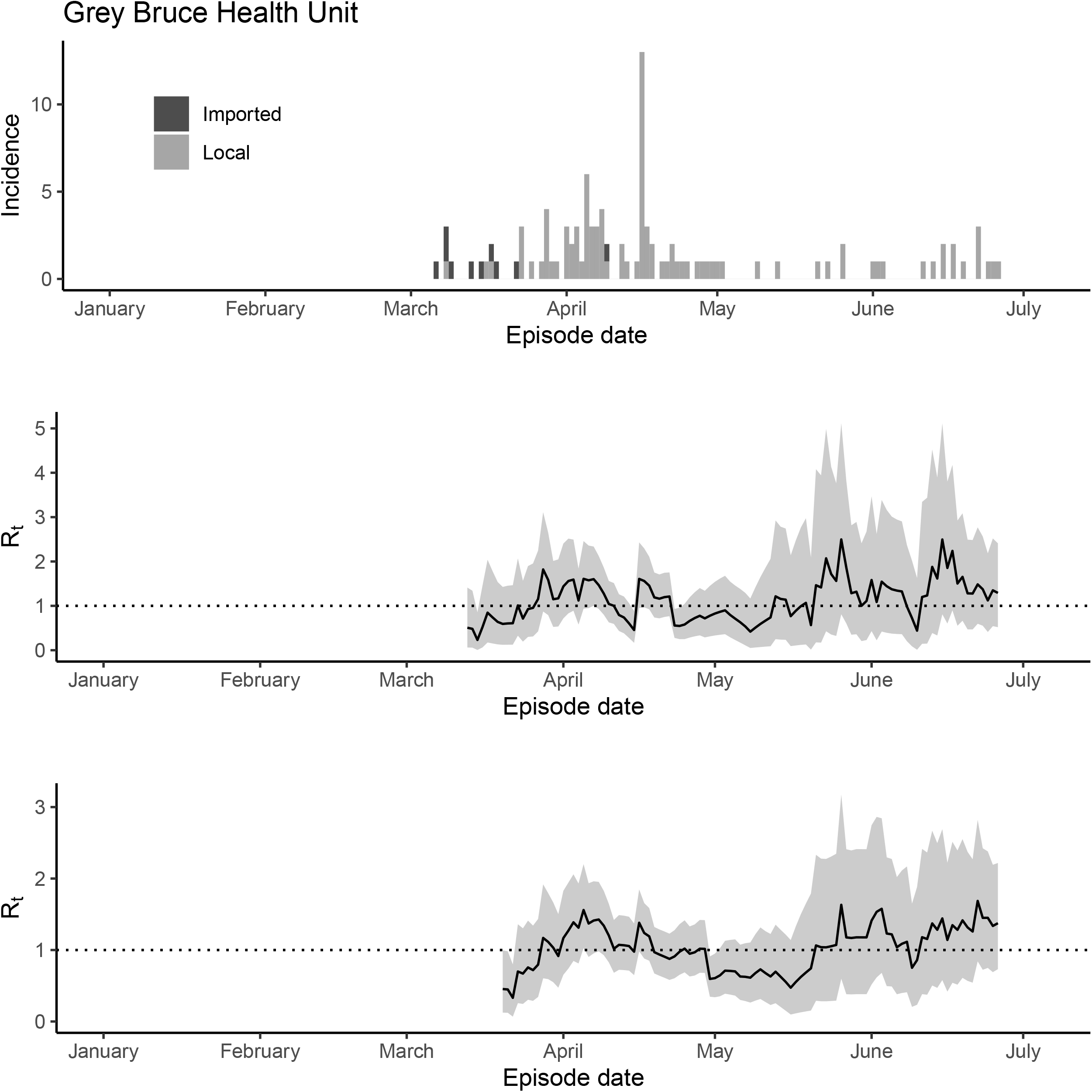
The epidemic trajectory of coronavirus disease 2019 in Grey Bruce Health Unit, Ontario, January 1-July 5, 2020: Upper panel: Daily number of new imported and local cases by date of symptom onset (aka accurate episode date) (upper panel); Rt with a 1-week window (middle panel); Rt with a 2-week window (lower panel).

**Figure S16.**
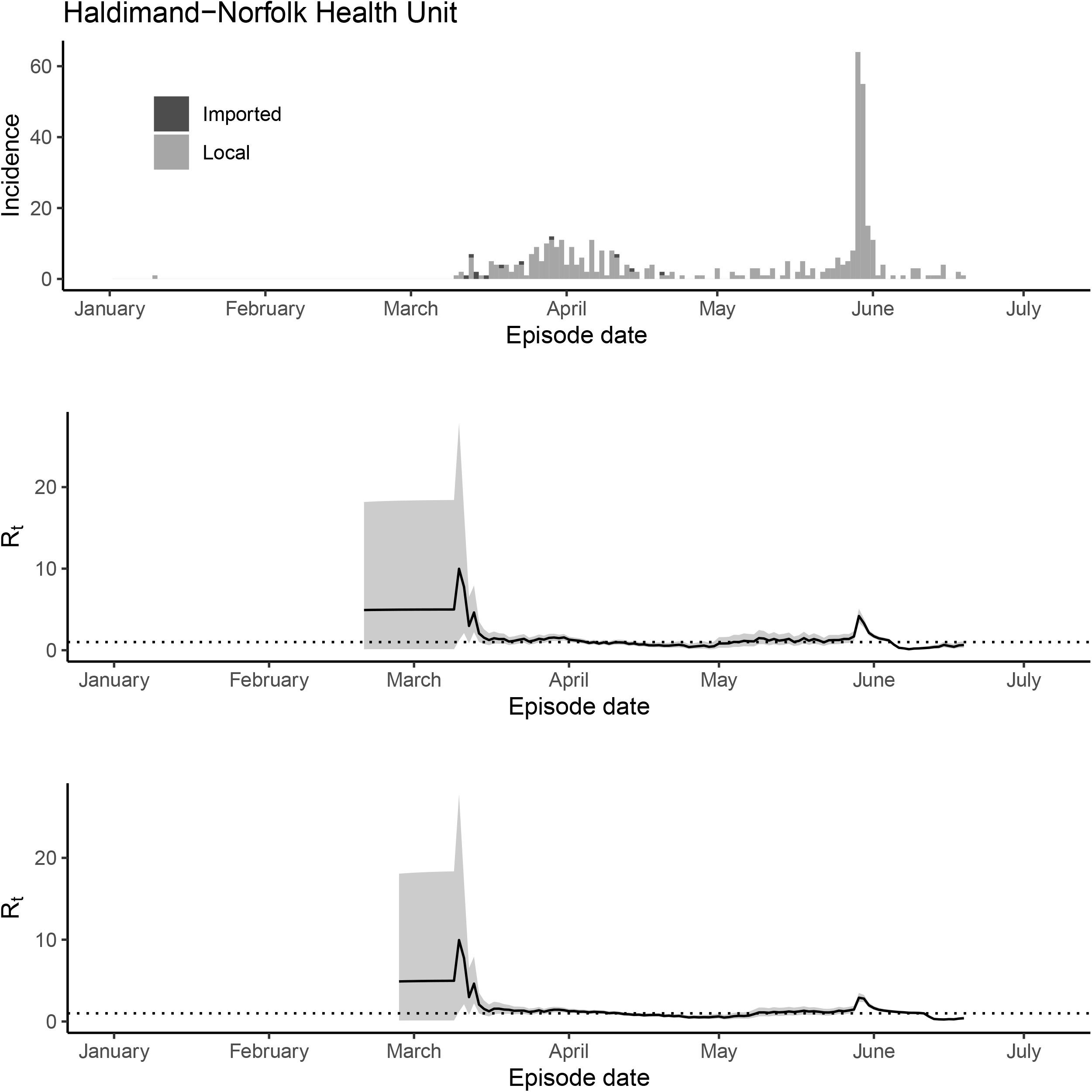
The epidemic trajectory of coronavirus disease 2019 in Haldimand-Norfolk Health Unit, Ontario, January 1-July 5, 2020: Upper panel: Daily number of new imported and local cases by date of symptom onset (aka accurate episode date) (upper panel); Rt with a 1-week window (middle panel); Rt with a 2-week window (lower panel).

**Figure S17.**
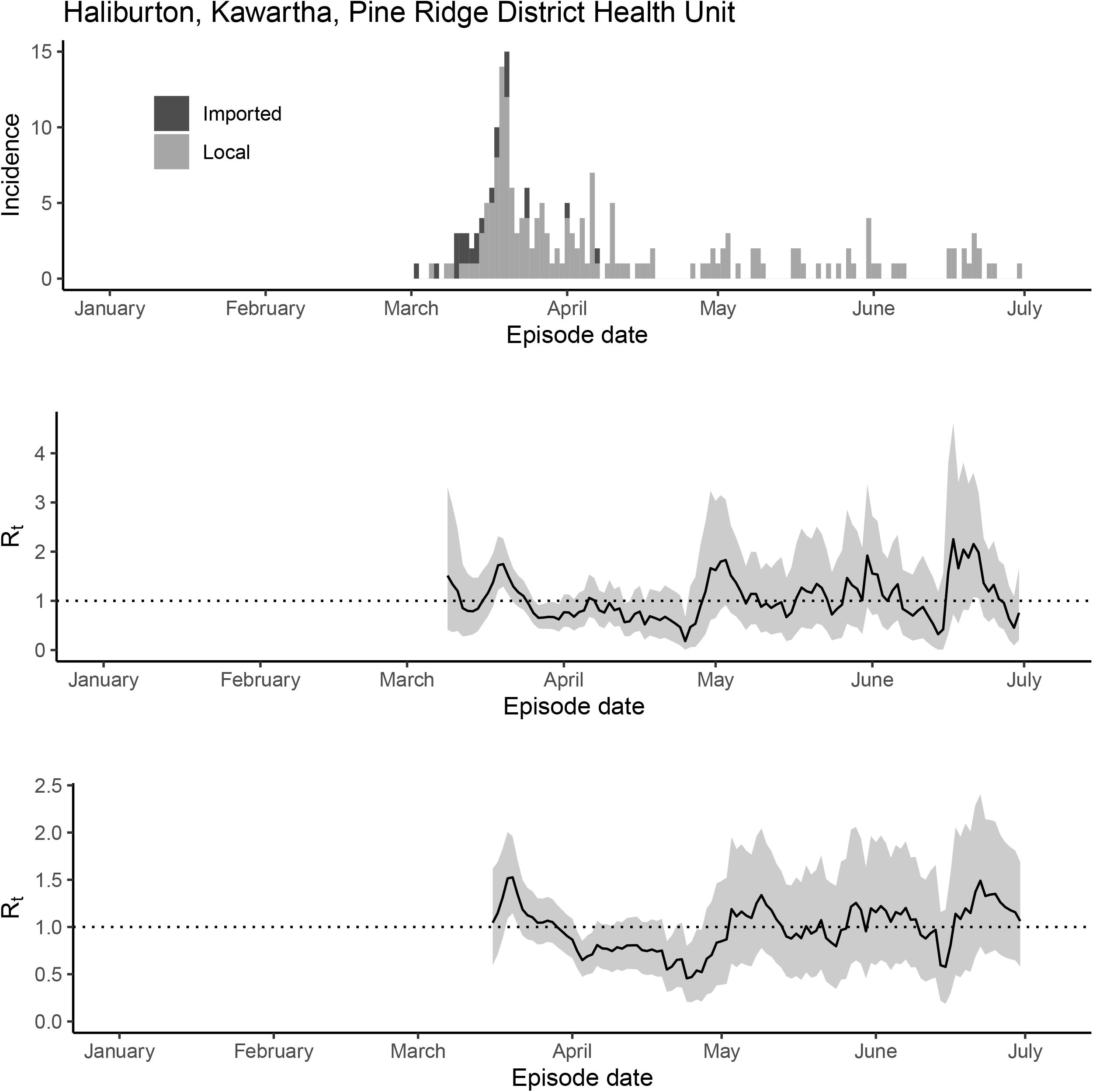
The epidemic trajectory of coronavirus disease 2019 in Haliburton, Kawartha, Pine Ridge District Health Unit, Ontario, January 1-July 5, 2020: Upper panel: Daily number of new imported and local cases by date of symptom onset (aka accurate episode date) (upper panel); Rt with a 1-week window (middle panel); Rt with a 2-week window (lower panel).

**Figure S18.**
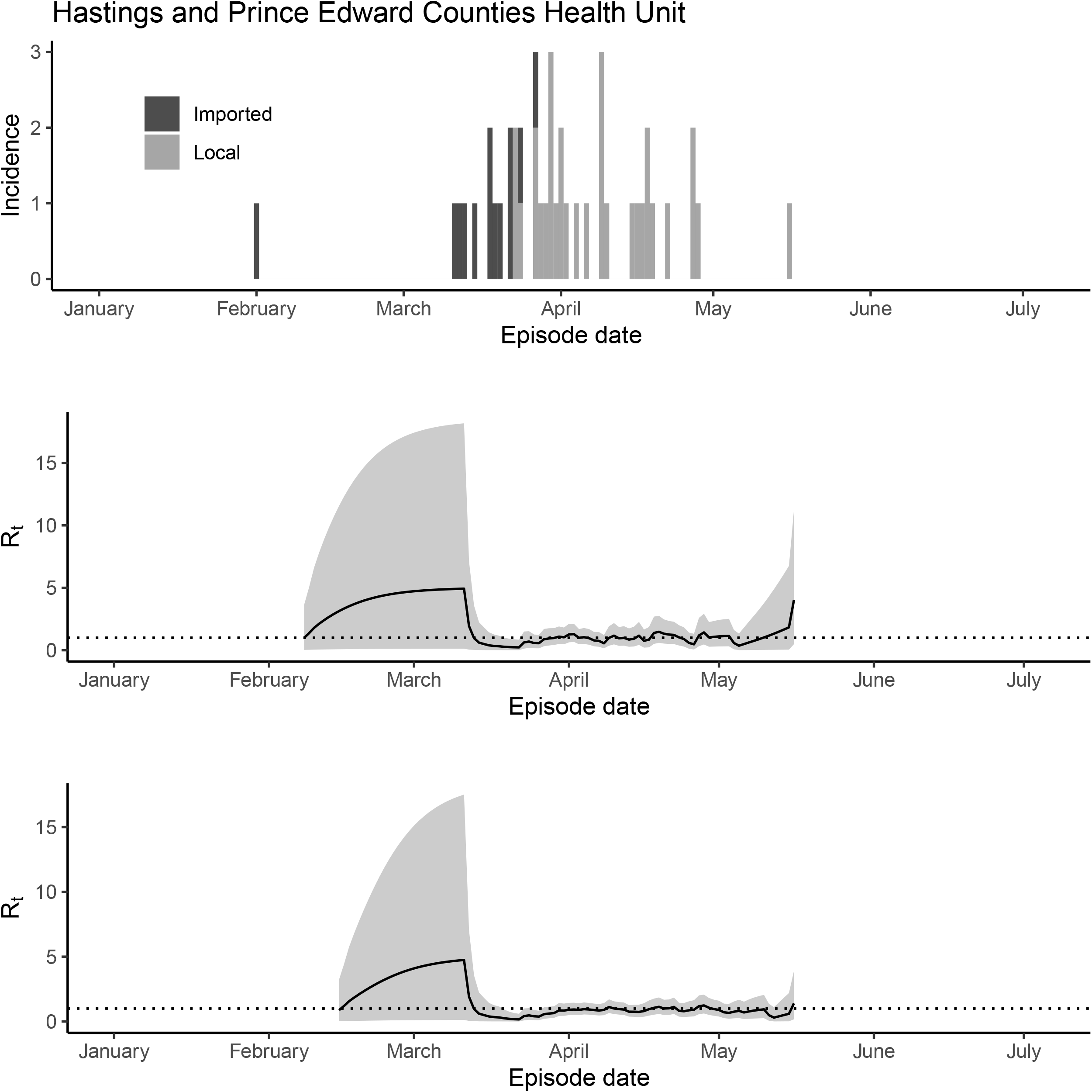
The epidemic trajectory of coronavirus disease 2019 in Hastings and Prince Edward Counties Health Unit, Ontario, January 1-July 5, 2020: Upper panel: Daily number of new imported and local cases by date of symptom onset (aka accurate episode date) (upper panel); Rt with a 1-week window (middle panel); Rt with a 2-week window (lower panel).

**Figure S19.**
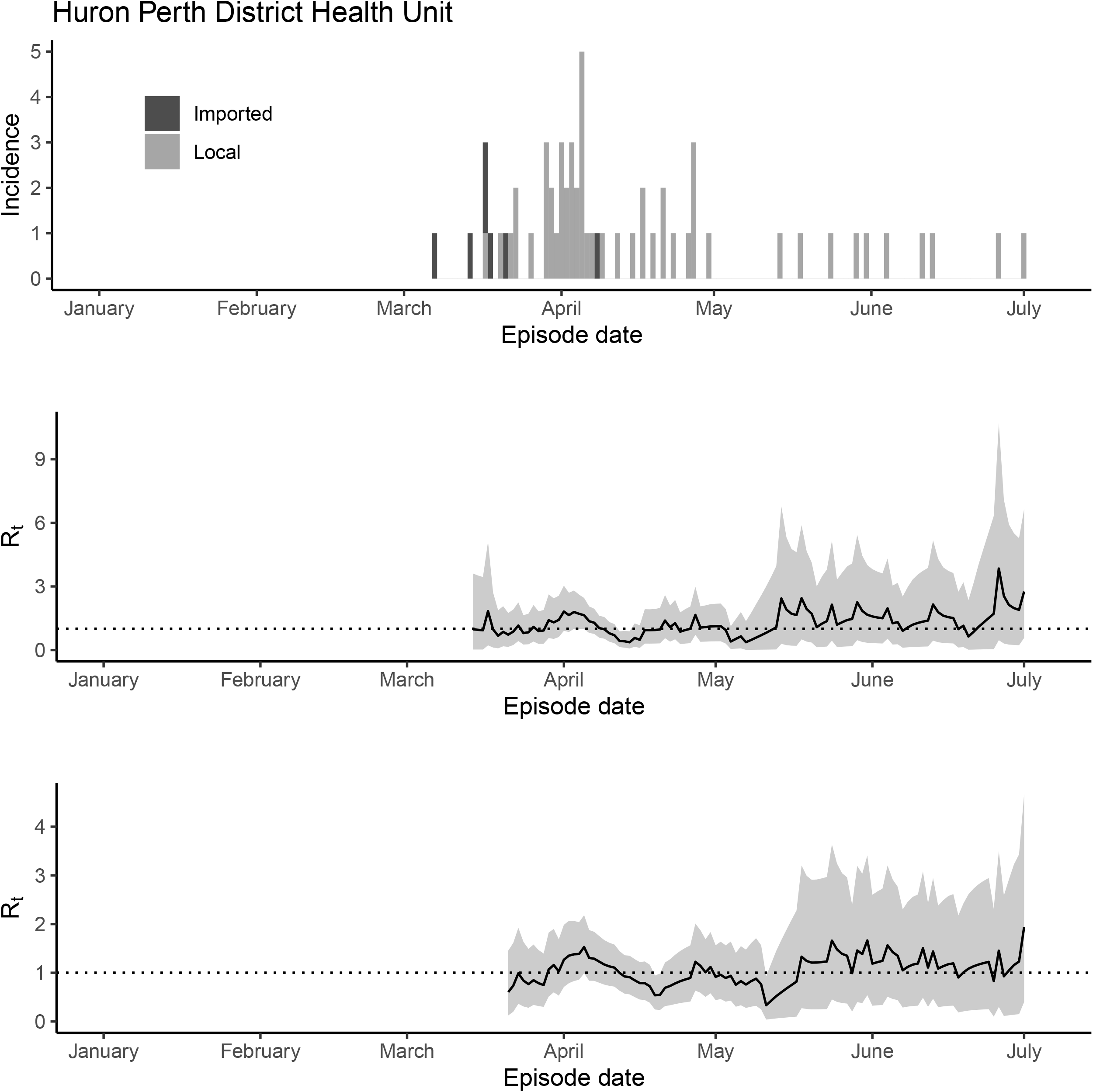
The epidemic trajectory of coronavirus disease 2019 in Huron Perth District Health Unit, Ontario, January 1-July 5, 2020: Upper panel: Daily number of new imported and local cases by date of symptom onset (aka accurate episode date) (upper panel); Rt with a 1-week window (middle panel); Rt with a 2-week window (lower panel).

**Figure S20.**
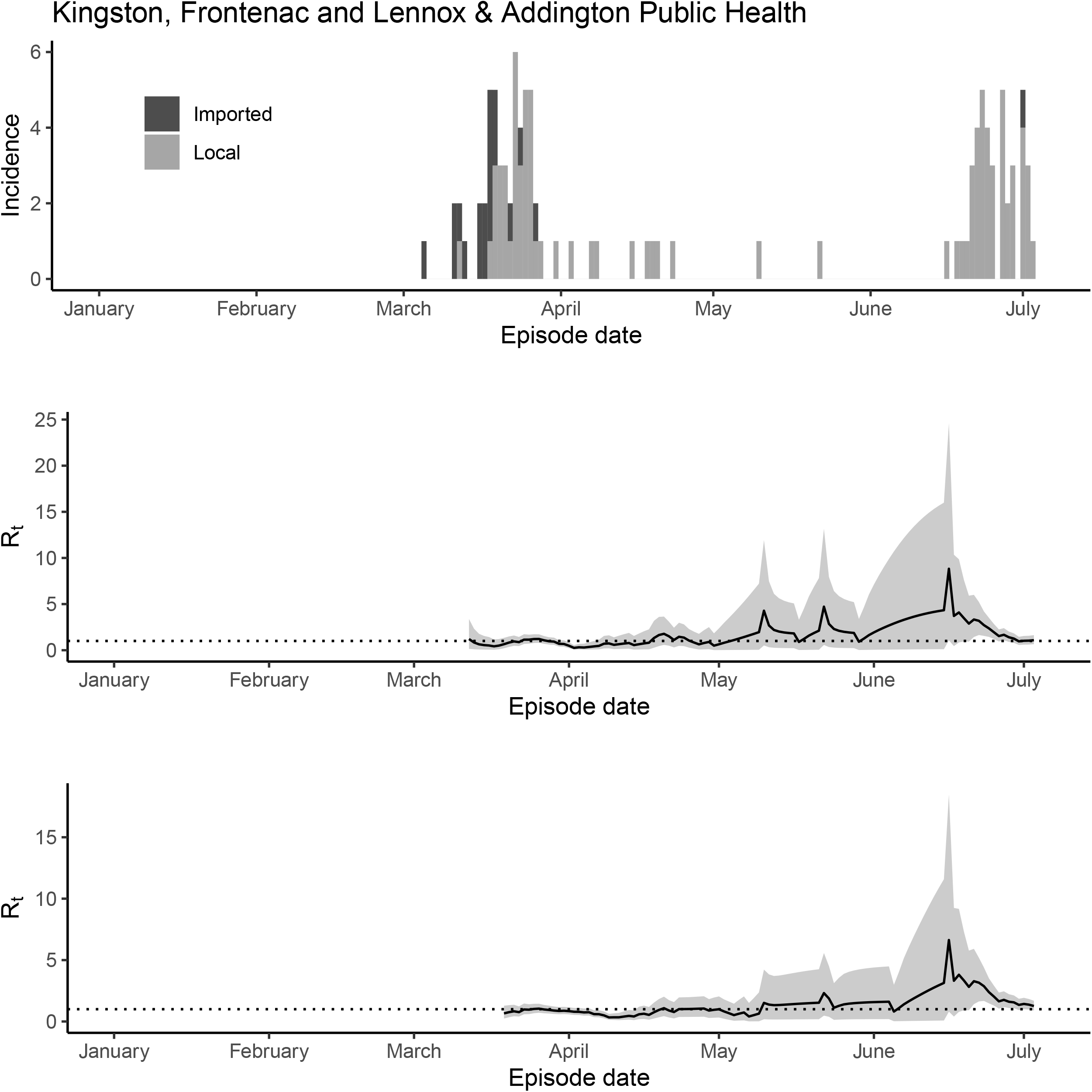
The epidemic trajectory of coronavirus disease 2019 in Kingston, Frontenac and Lennox & Addington Public Health, Ontario, January 1-July 5, 2020: Upper panel: Daily number of new imported and local cases by date of symptom onset (aka accurate episode date) (upper panel); Rt with a 1-week window (middle panel); Rt with a 2-week window (lower panel).

**Figure S21.**
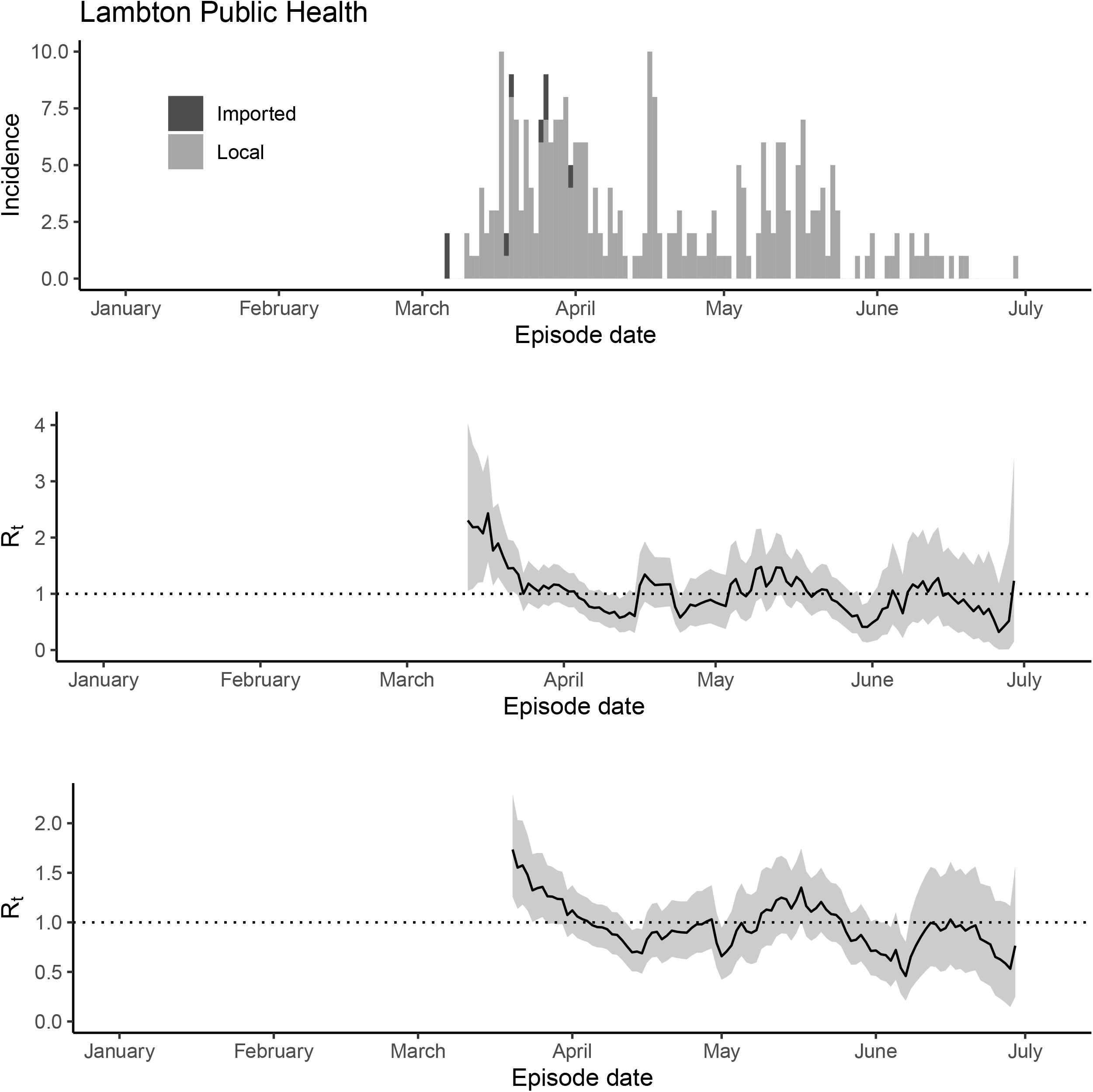
The epidemic trajectory of coronavirus disease 2019 in Lambton Public Health, Ontario, January 1-July 5, 2020: Upper panel: Daily number of new imported and local cases by date of symptom onset (aka accurate episode date) (upper panel); Rt with a 1-week window (middle panel); Rt with a 2-week window (lower panel).

**Figure S22.**
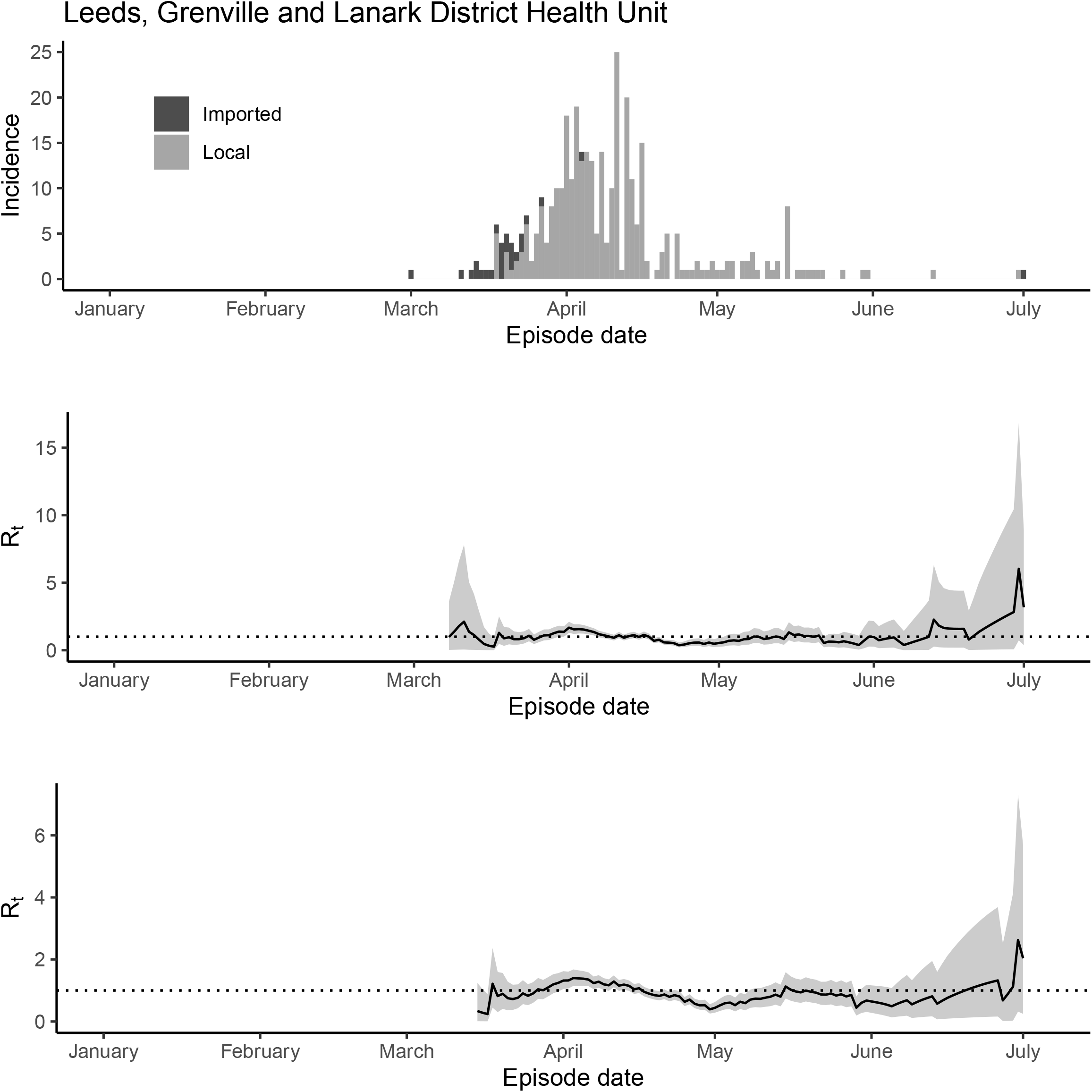
The epidemic trajectory of coronavirus disease 2019 in Leeds, Grenville and Lanark District Health Unit, Ontario, January 1-July 5, 2020: Upper panel: Daily number of new imported and local cases by date of symptom onset (aka accurate episode date) (upper panel); Rt with a 1-week window (middle panel); Rt with a 2-week window (lower panel).

**Figure S23.**
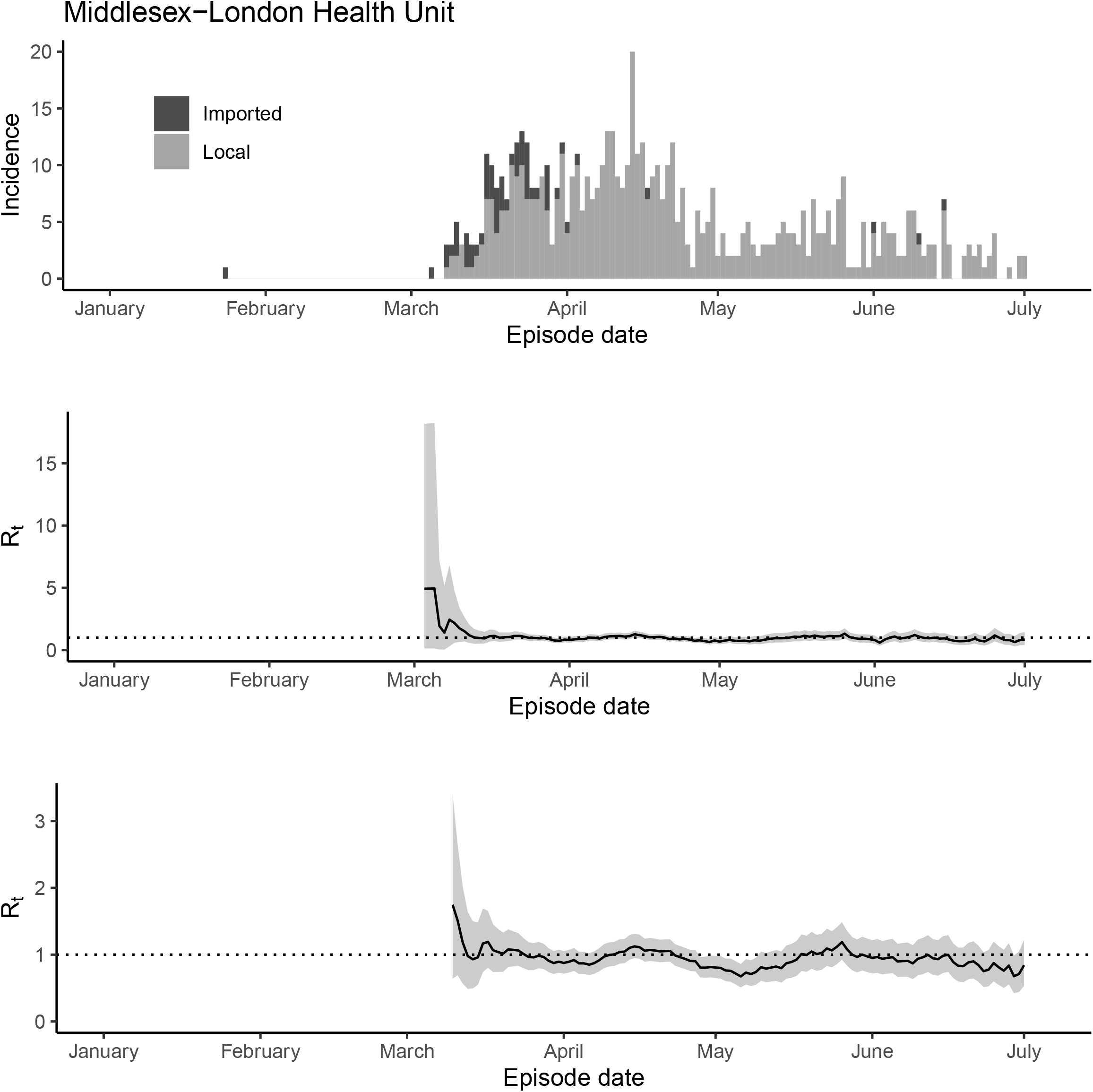
The epidemic trajectory of coronavirus disease 2019 in Middlesex-London Health Unit, Ontario, January 1-July 5, 2020: Upper panel: Daily number of new imported and local cases by date of symptom onset (aka accurate episode date) (upper panel); Rt with a 1-week window (middle panel); Rt with a 2-week window (lower panel).

**Figure S24.**
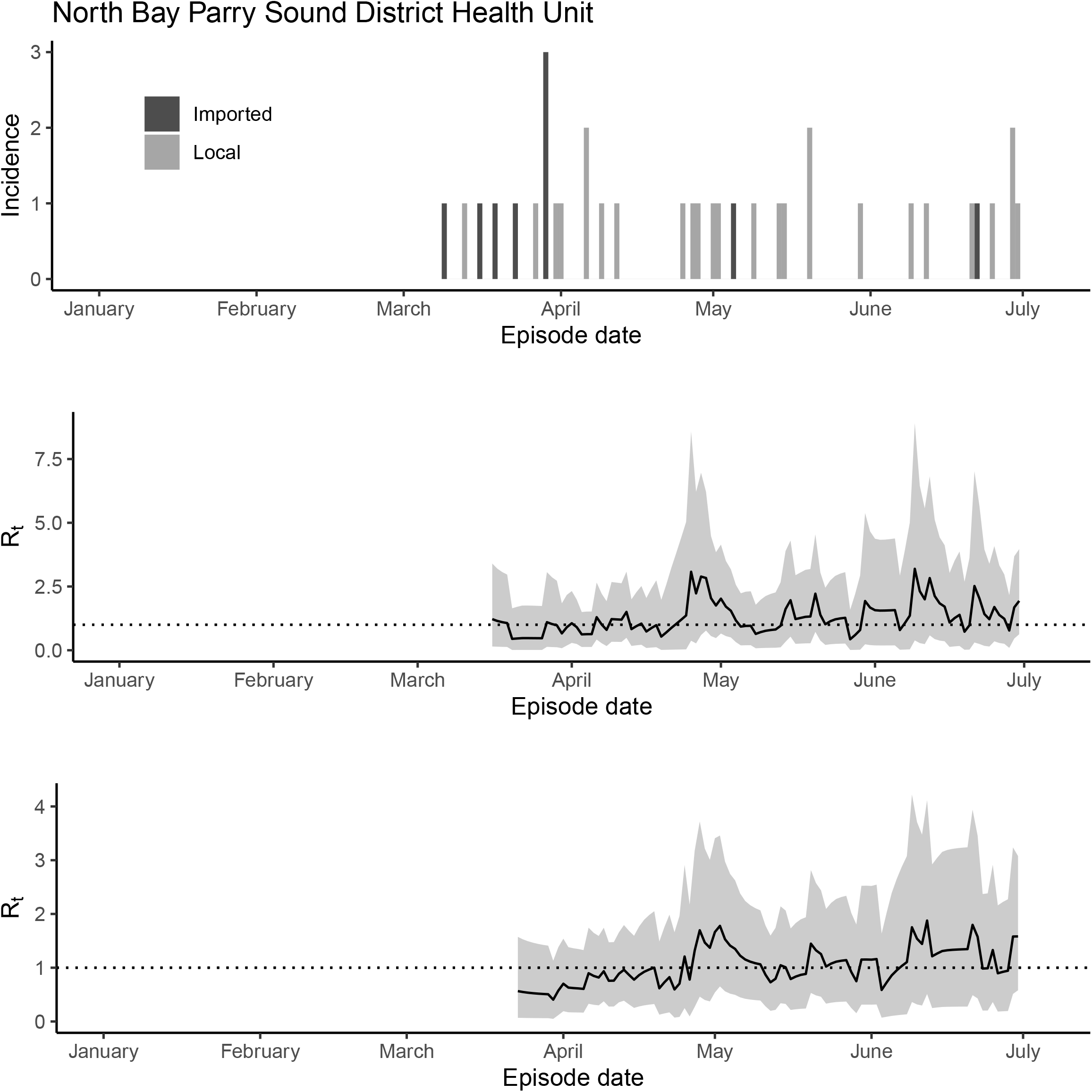
The epidemic trajectory of coronavirus disease 2019 in North Bay Parry Sound District Health Unit, Ontario, January 1-July 5, 2020: Upper panel: Daily number of new imported and local cases by date of symptom onset (aka accurate episode date) (upper panel); Rt with a 1-week window (middle panel); Rt with a 2-week window (lower panel).

**Figure S25.**
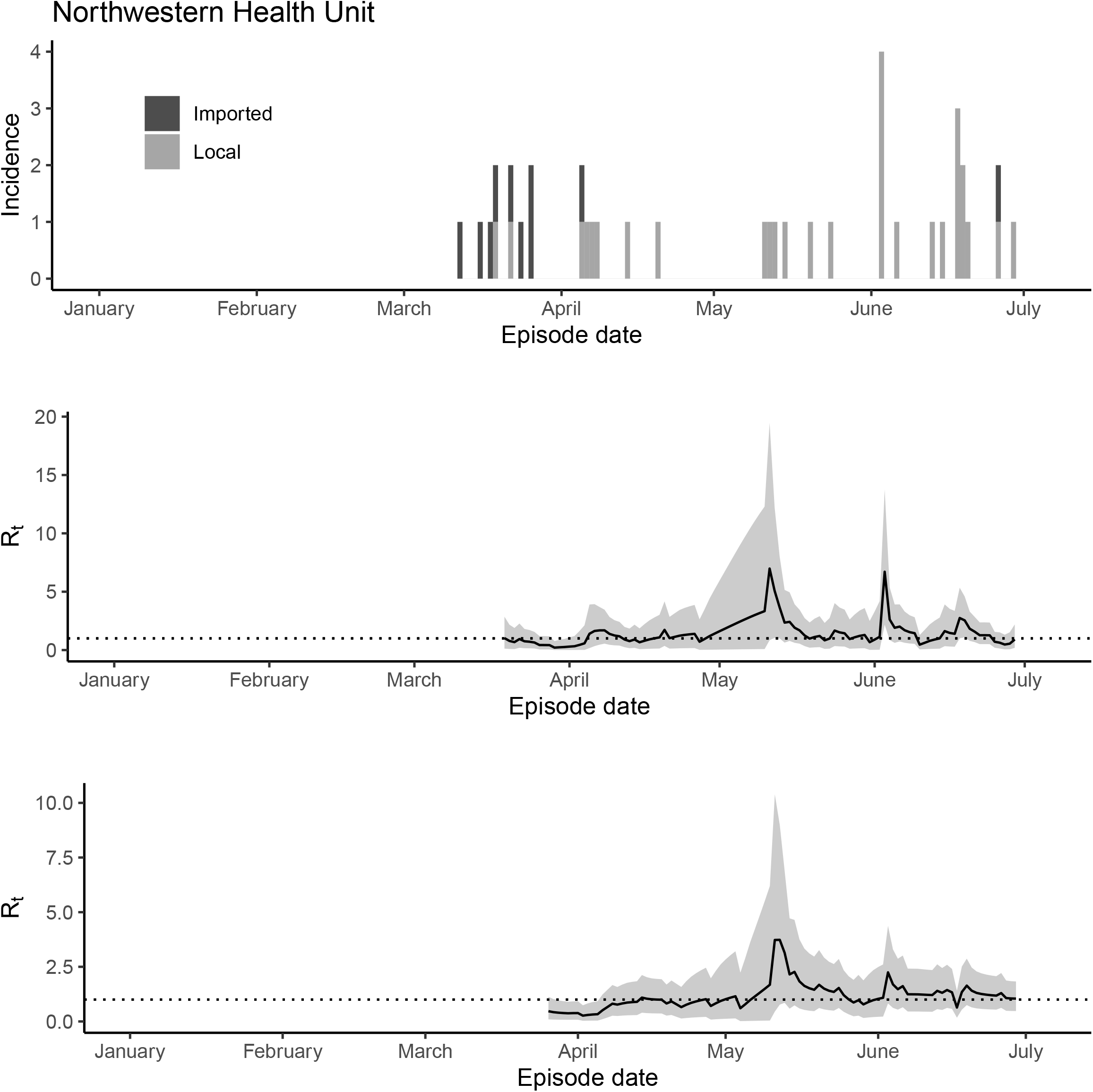
The epidemic trajectory of coronavirus disease 2019 in Northwestern Health Unit, Ontario, January 1-July 5, 2020: Upper panel: Daily number of new imported and local cases by date of symptom onset (aka accurate episode date) (upper panel); Rt with a 1-week window (middle panel); Rt with a 2-week window (lower panel).

**Figure S26.**
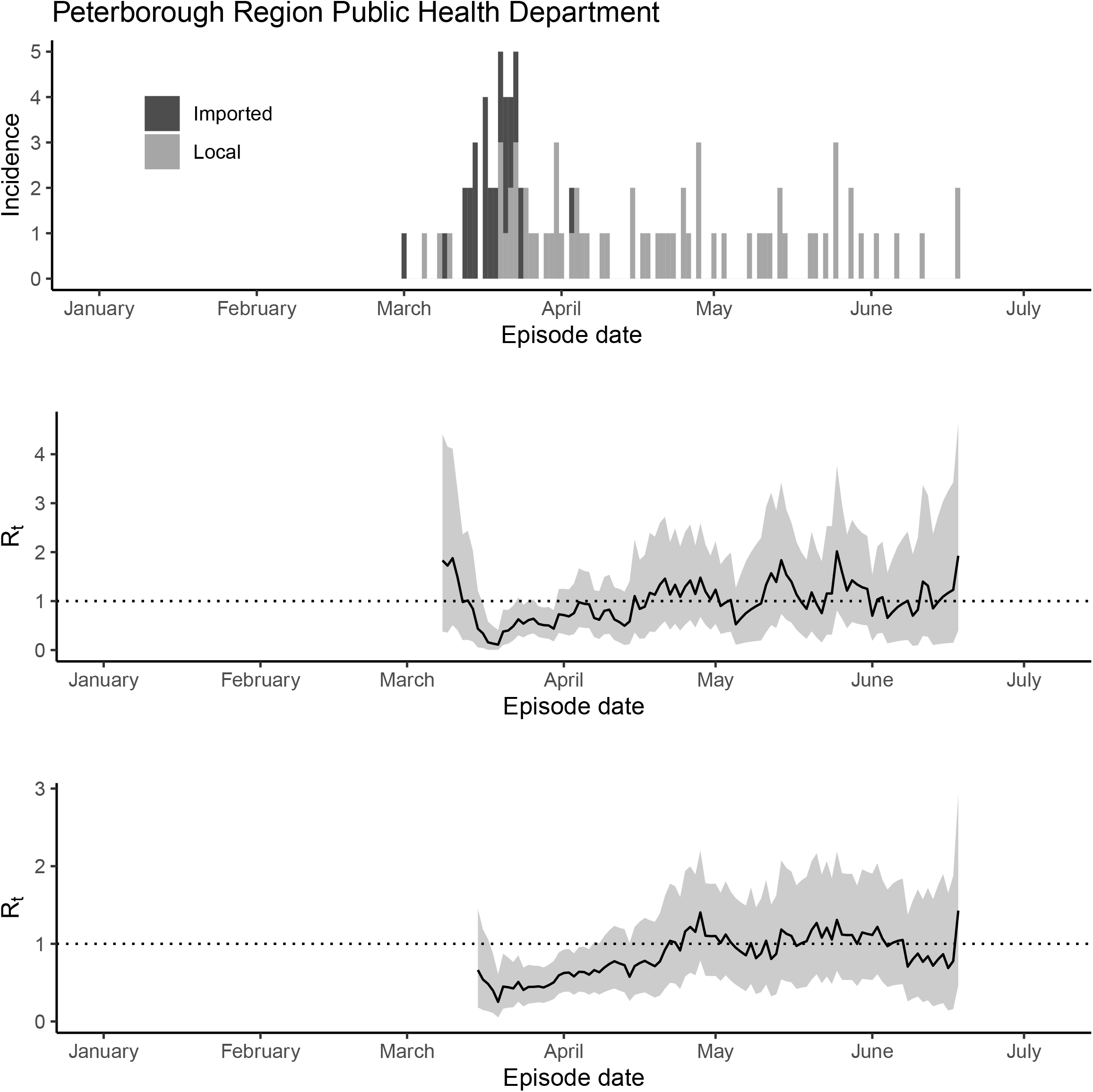
The epidemic trajectory of coronavirus disease 2019 in Peterborough Public Health, Ontario, January 1-July 5, 2020: Upper panel: Daily number of new imported and local cases by date of symptom onset (aka accurate episode date) (upper panel); Rt with a 1-week window (middle panel); Rt with a 2-week window (lower panel).

**Figure S27.**
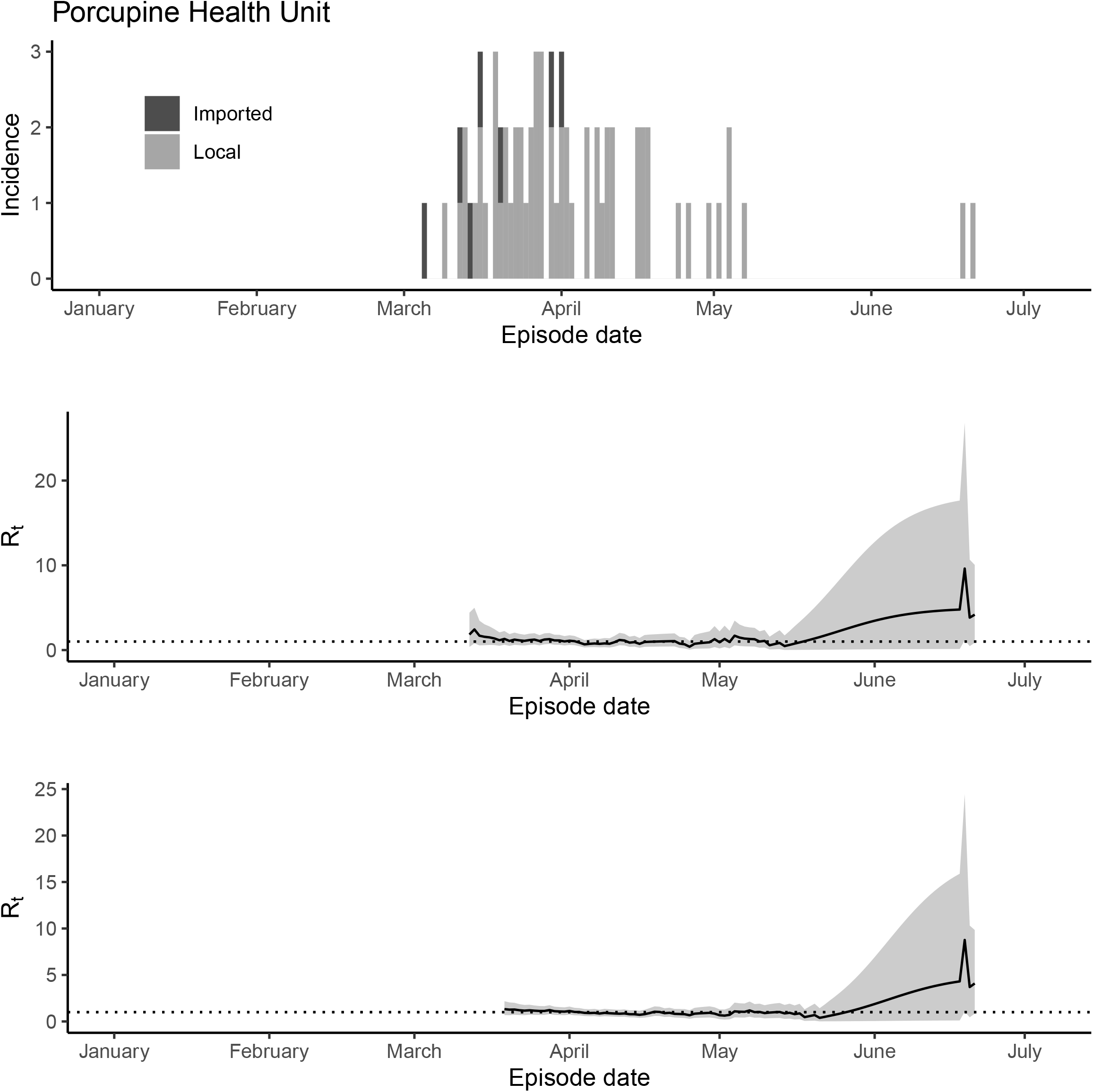
The epidemic trajectory of coronavirus disease 2019 in Porcupine Health Unit, Ontario, January 1-July 5, 2020: Upper panel: Daily number of new imported and local cases by date of symptom onset (aka accurate episode date) (upper panel); Rt with a 1-week window (middle panel); Rt with a 2-week window (lower panel).

**Figure S28.**
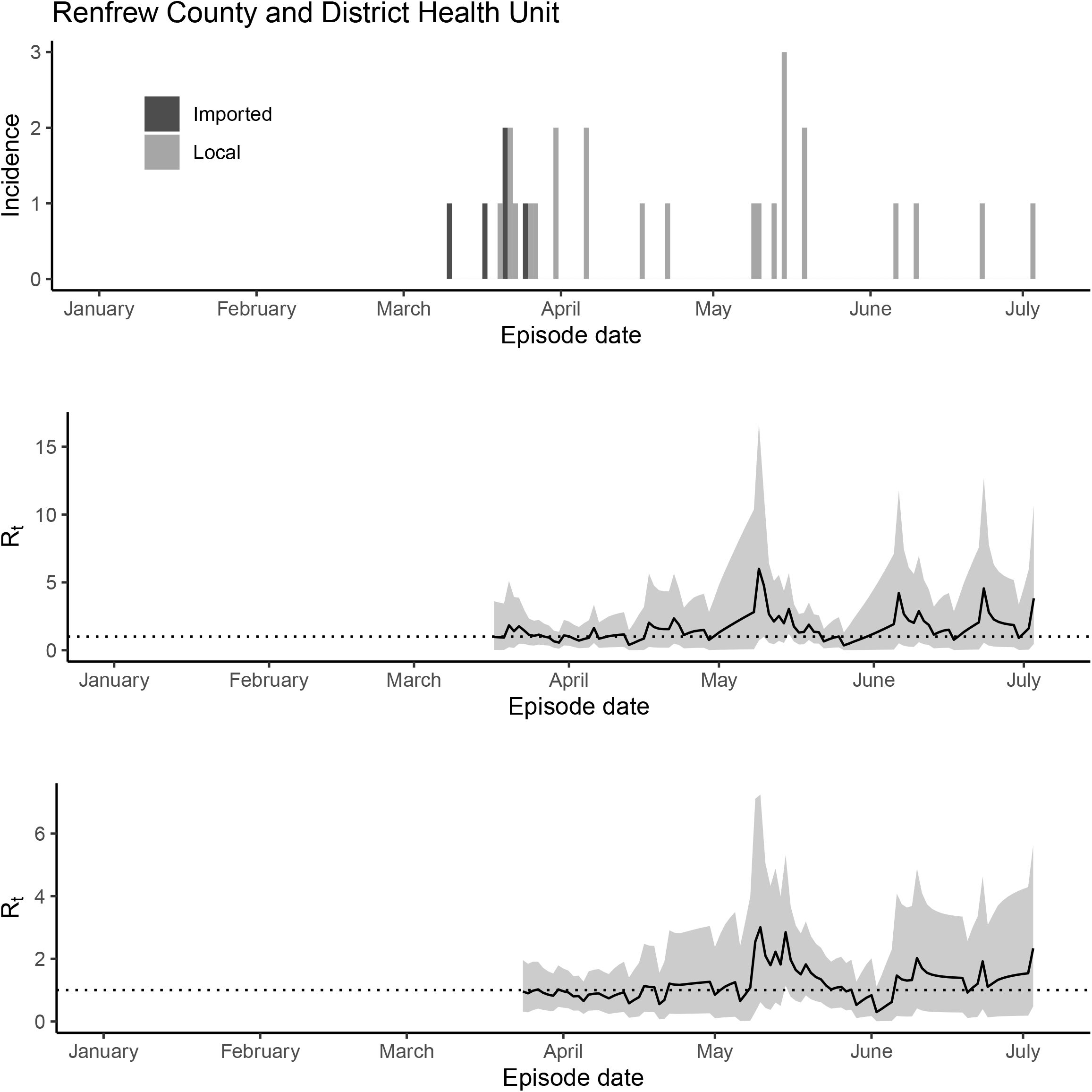
The epidemic trajectory of coronavirus disease 2019 in Renfrew County and District Health Unit, Ontario, January 1-July 5, 2020: Upper panel: Daily number of new imported and local cases by date of symptom onset (aka accurate episode date) (upper panel); Rt with a 1-week window (middle panel); Rt with a 2-week window (lower panel).

**Figure S29.**
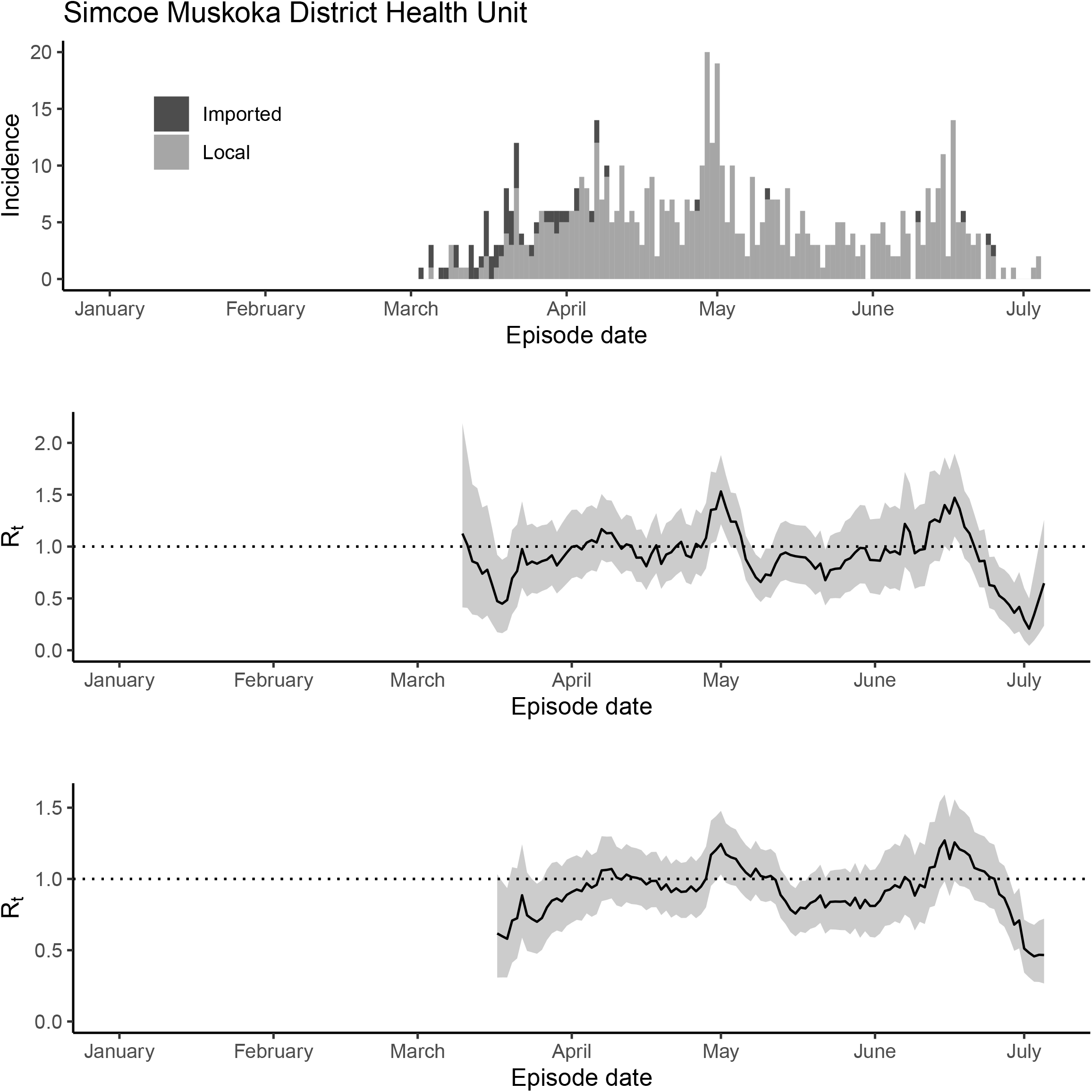
The epidemic trajectory of coronavirus disease 2019 in Simcoe Muskoka District Health Unit, Ontario, January 1-July 5, 2020: Upper panel: Daily number of new imported and local cases by date of symptom onset (aka accurate episode date) (upper panel); Rt with a 1-week window (middle panel); Rt with a 2-week window (lower panel).

**Figure S30.**
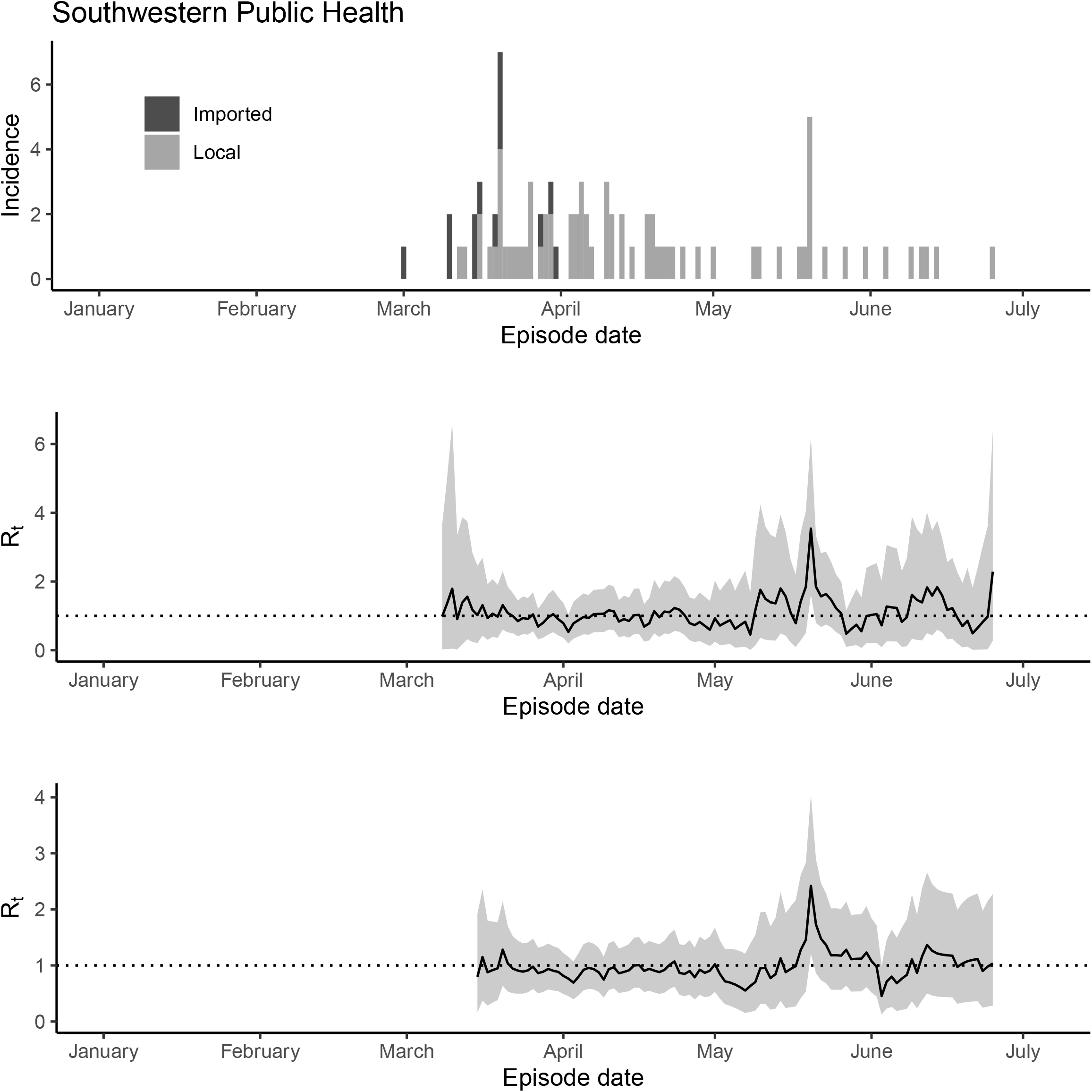
The epidemic trajectory of coronavirus disease 2019 in Southwestern Health Unit, Ontario, January 1-July 5, 2020: Upper panel: Daily number of new imported and local cases by date of symptom onset (aka accurate episode date) (upper panel); Rt with a 1-week window (middle panel); Rt with a 2-week window (lower panel).

**Figure S31.**
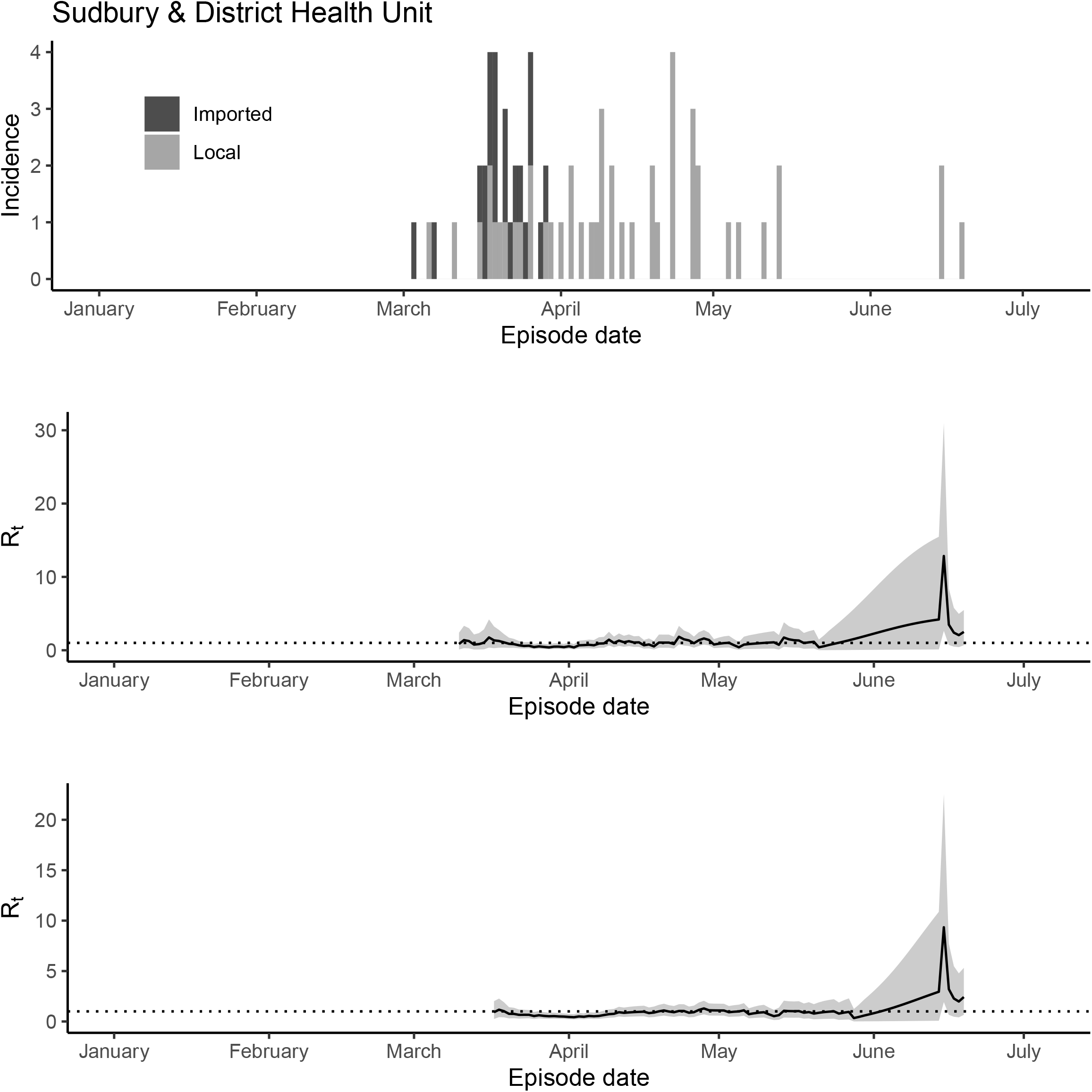
The epidemic trajectory of coronavirus disease 2019 in Sudbury & Districts Health Unit, Ontario, January 1-July 5, 2020: Upper panel: Daily number of new imported and local cases by date of symptom onset (aka accurate episode date) (upper panel); Rt with a 1-week window (middle panel); Rt with a 2-week window (lower panel).

**Figure S32.**
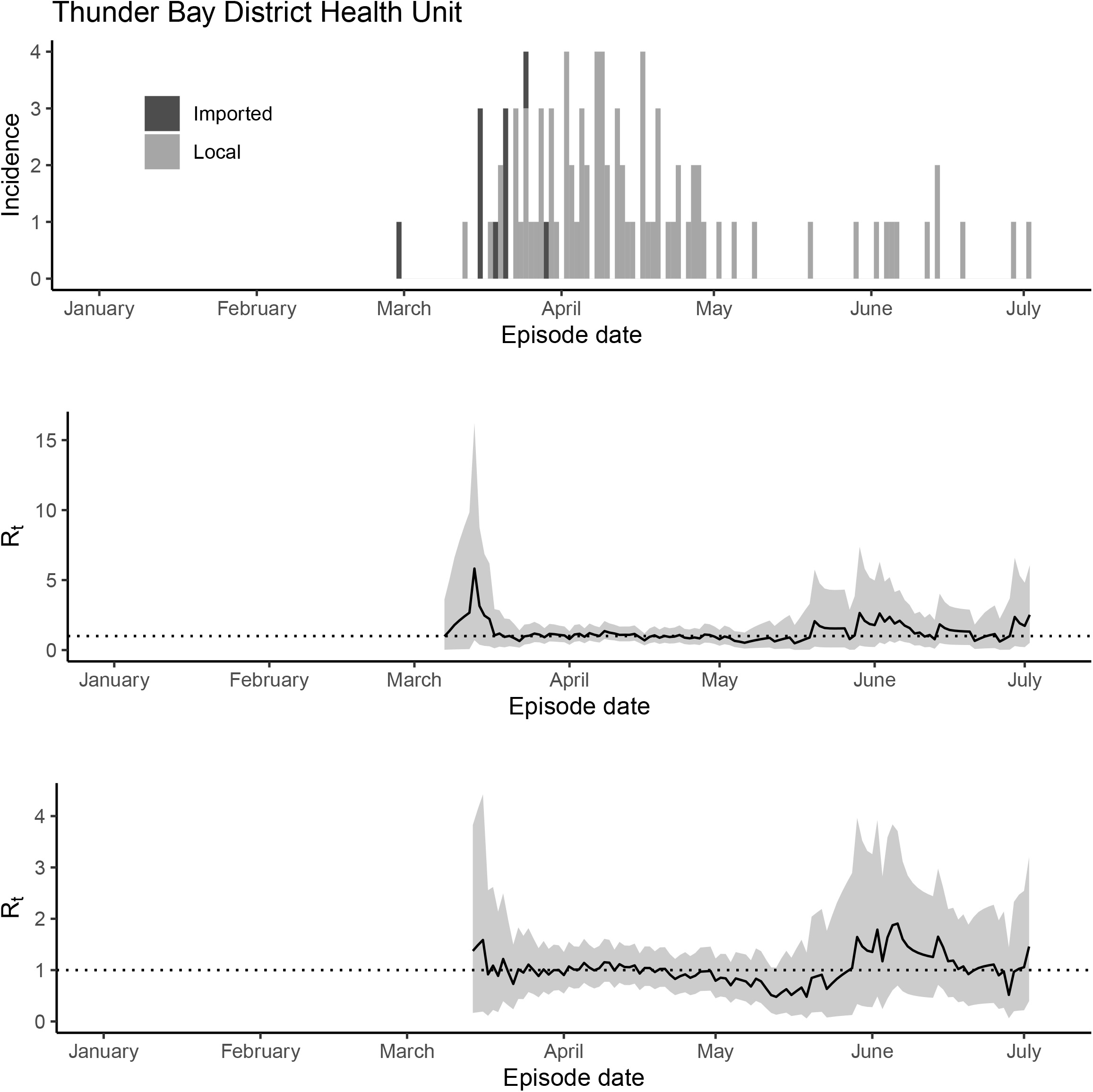
The epidemic trajectory of coronavirus disease 2019 in Thunder Bay District Health Unit, Ontario, January 1-July 5, 2020: Upper panel: Daily number of new imported and local cases by date of symptom onset (aka accurate episode date) (upper panel); Rt with a 1-week window (middle panel); Rt with a 2-week window (lower panel).

**Figure S33.**
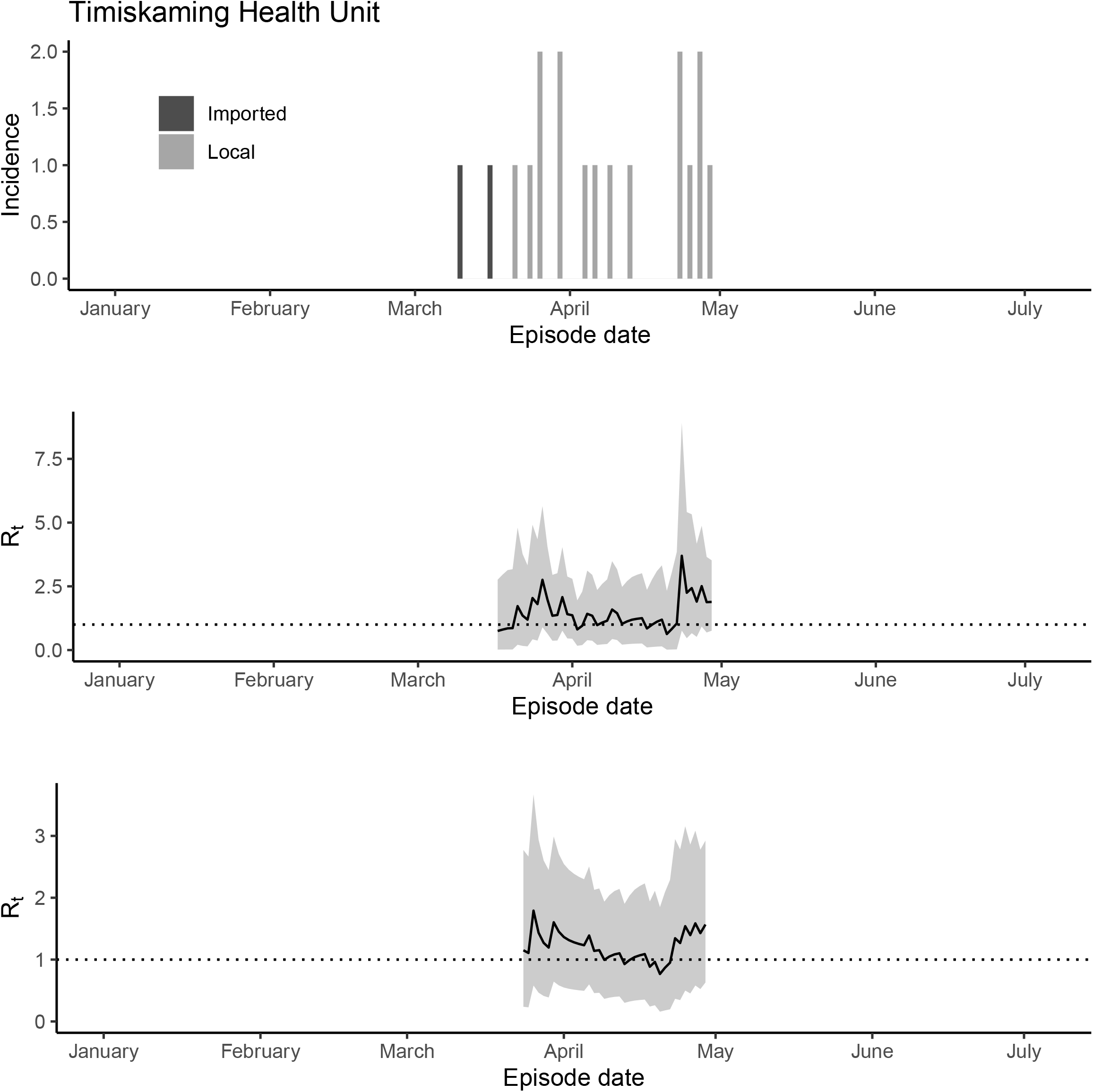
The epidemic trajectory of coronavirus disease 2019 in Timiskaming Health Unit, Ontario, January 1-July 5, 2020: Upper panel: Daily number of new imported and local cases by date of symptom onset (aka accurate episode date) (upper panel); Rt with a 1-week window (middle panel); Rt with a 2-week window (lower panel).

**Figure S34.**
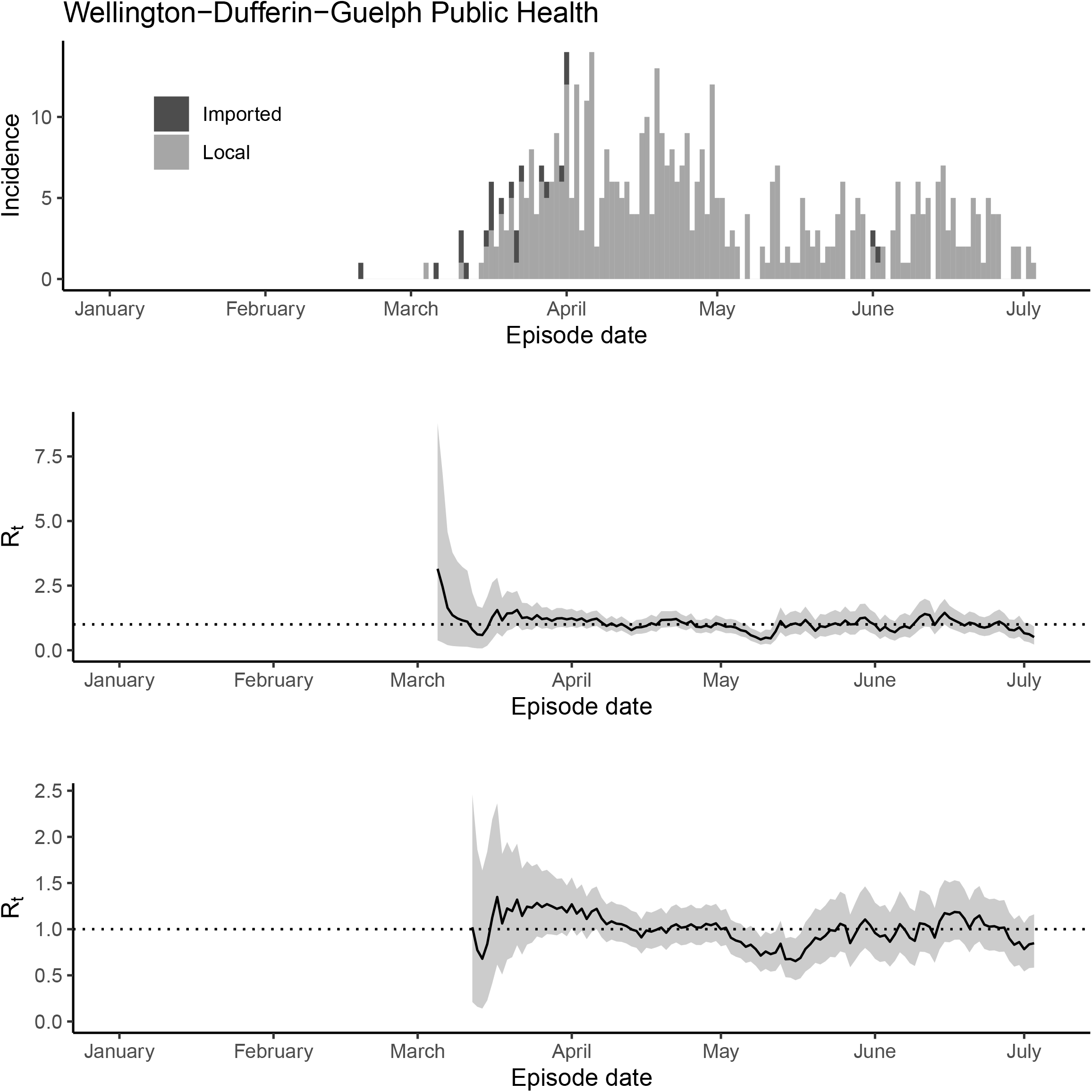
The epidemic trajectory of coronavirus disease 2019 in Windsor-Dufferin-Guelph Public Health, Ontario, January 1-July 5, 2020: Upper panel: Daily number of new imported and local cases by date of symptom onset (aka accurate episode date) (upper panel); Rt with a 1-week window (middle panel); Rt with a 2-week window (lower panel).

**Figure S35.**
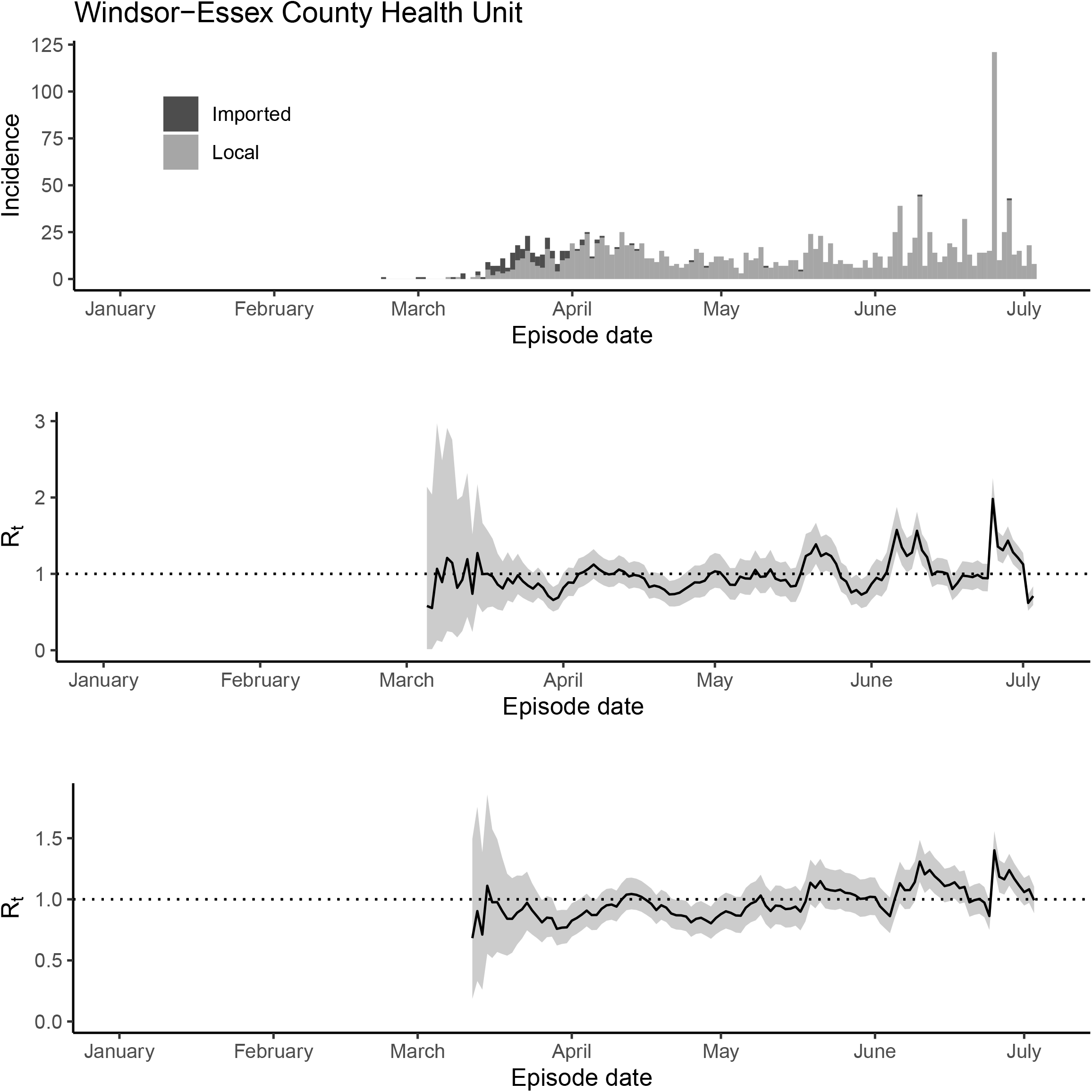
The epidemic trajectory of coronavirus disease 2019 in Windsor-Dufferin-Guelph Public Health, Ontario, January 1-July 5, 2020: Upper panel: Daily number of new imported and local cases by date of symptom onset (aka accurate episode date) (upper panel); Rt with a 1-week window (middle panel); Rt with a 2-week window (lower panel).

**Figure S36.**
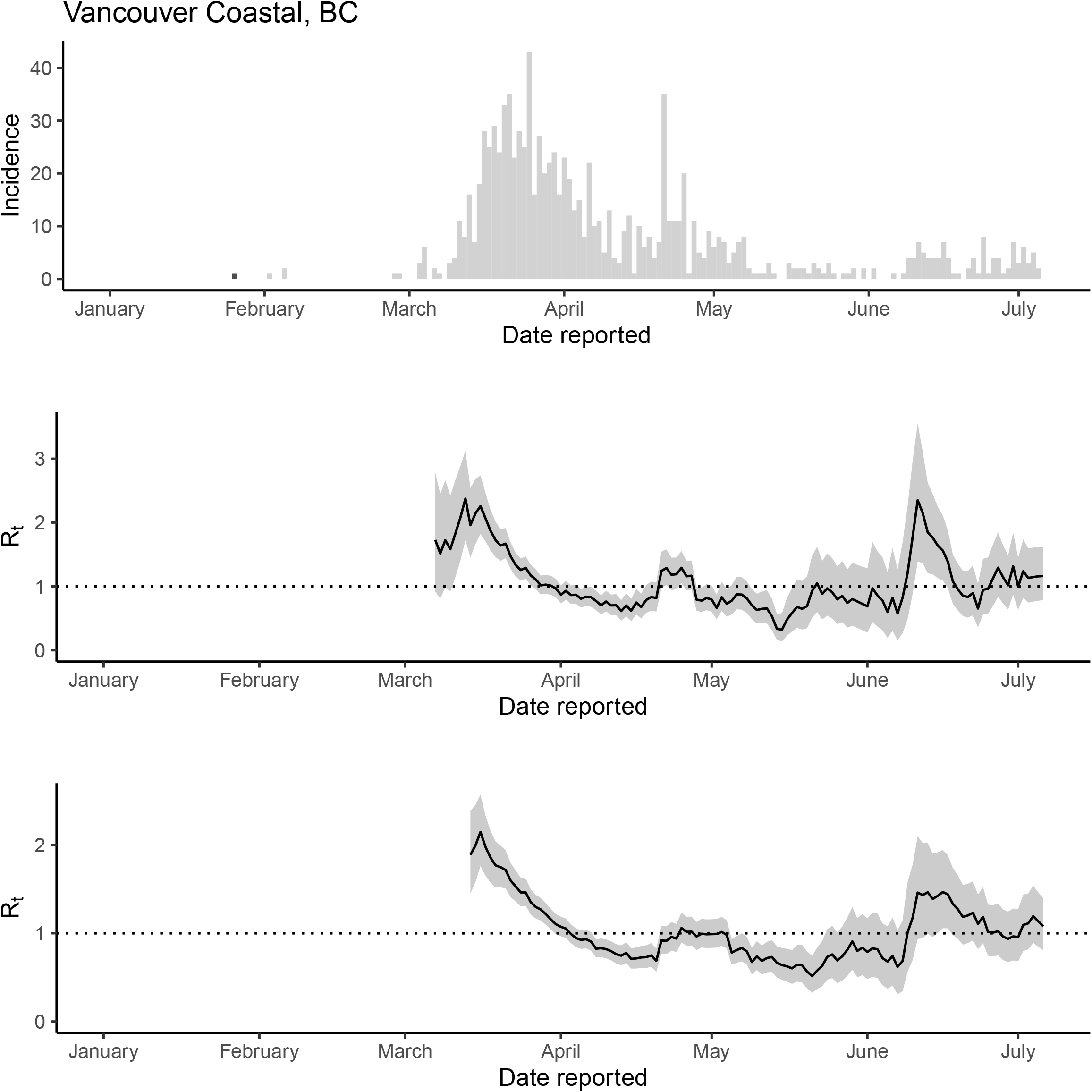
The epidemic trajectory of coronavirus disease 2019 in Vancouver Coastal Health Authority, British Columbia, January 1-July 6, 2020: Daily number of new cases by date of report (upper panel); *R_t_* with a 1-week window (middle panel); *R_t_* with a 2-week window (lower panel).

**Figure S37.**
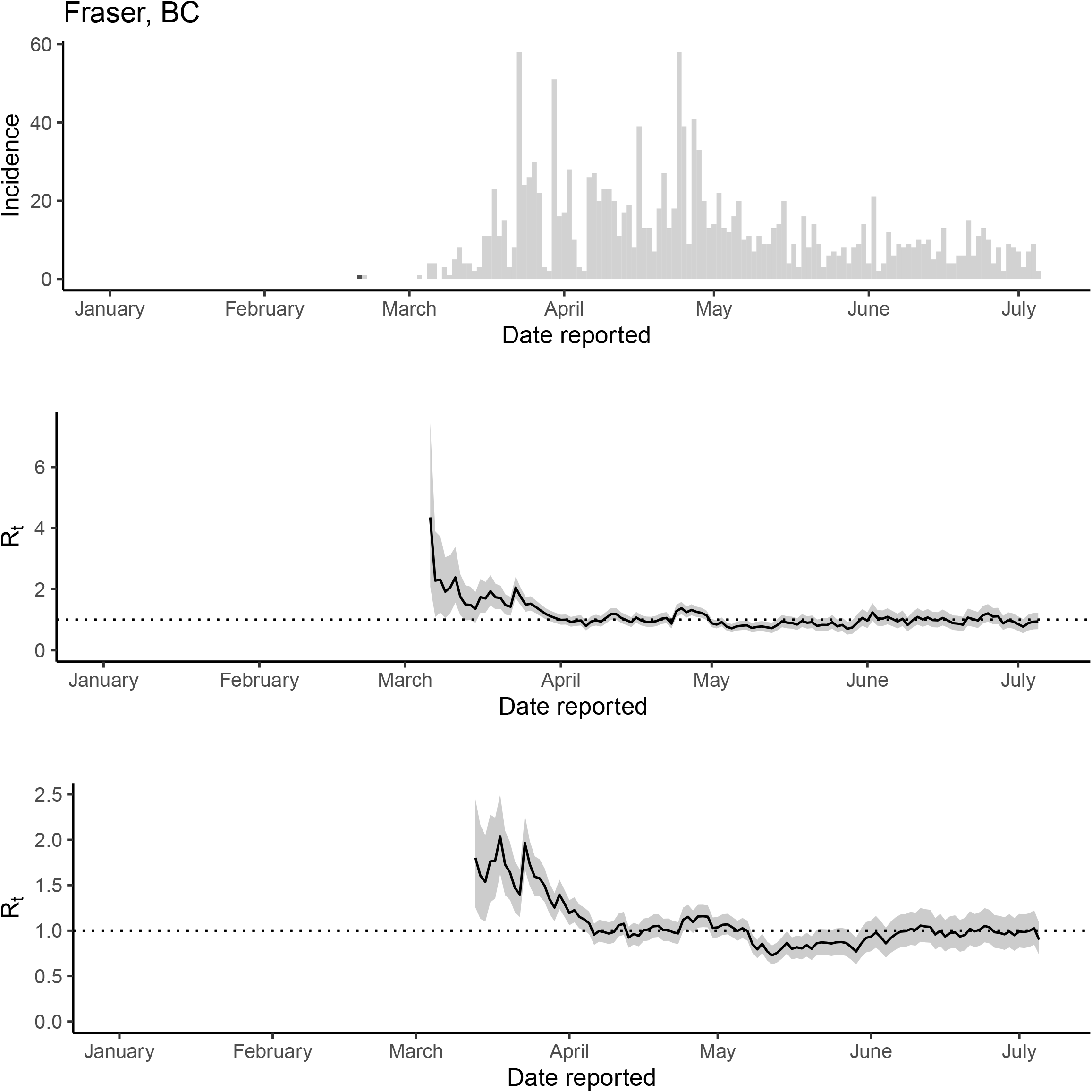
The epidemic trajectory of coronavirus disease 2019 in Fraser Health Authority, British Columbia, January 1-July 6, 2020: Daily number of new cases by date of report (upper panel); *R_t_* with a 1-week window (middle panel); *R_t_* with a 2-week window (lower panel).

**Figure S38.**
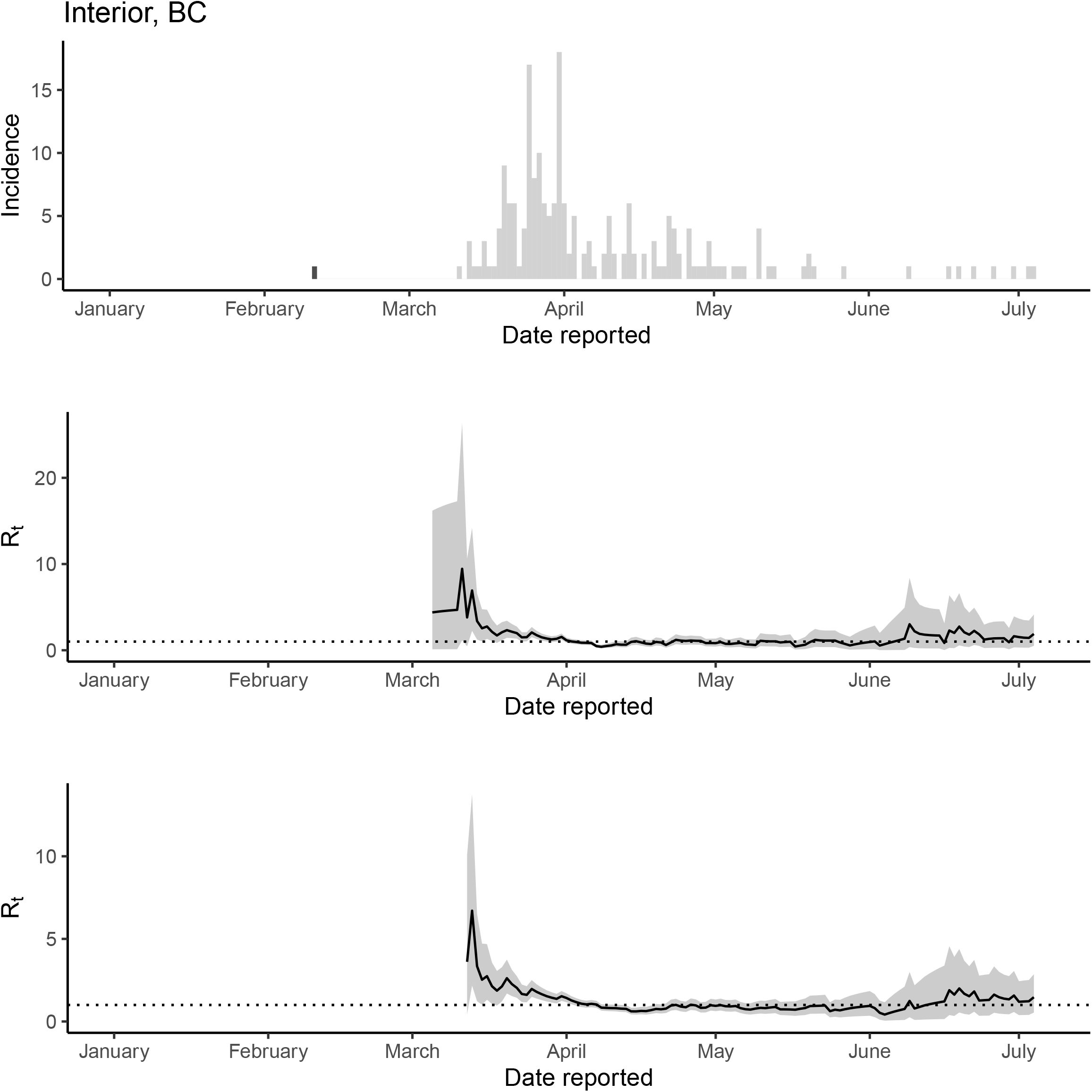
The epidemic trajectory of coronavirus disease 2019 in Interior Health Authority, British Columbia, January 1-July 6, 2020: Daily number of new cases by date of report (upper panel); *R_t_* with a 1-week window (middle panel); *R_t_* with a 2-week window (lower panel).

**Figure S39.**
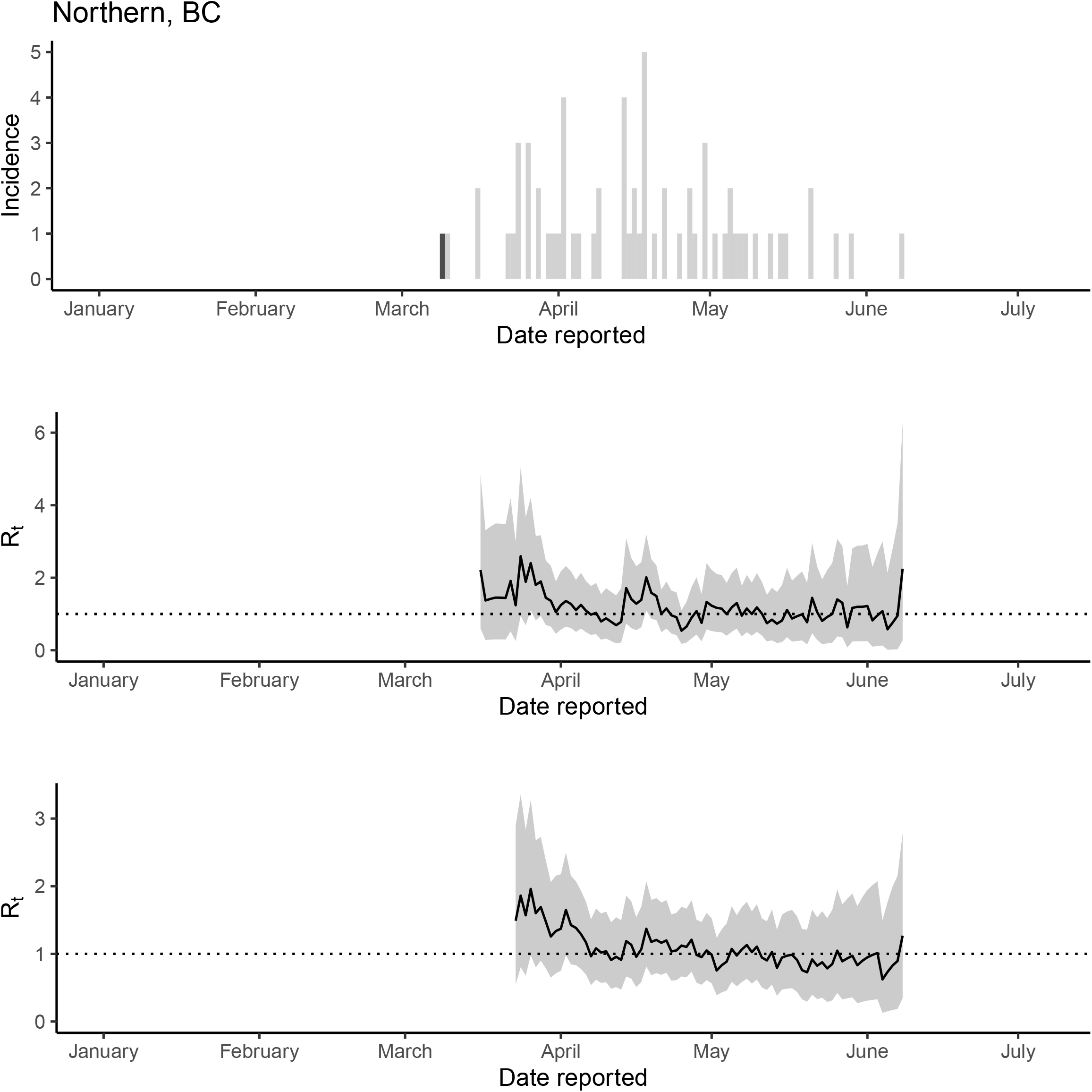
The epidemic trajectory of coronavirus disease 2019 in Northern Health Authority, British Columbia, January 1-July 6, 2020: Daily number of new cases by date of report (upper panel); *R_t_* with a 1-week window (middle panel); *R_t_* with a 2-week window (lower panel).

**Figure S40.**
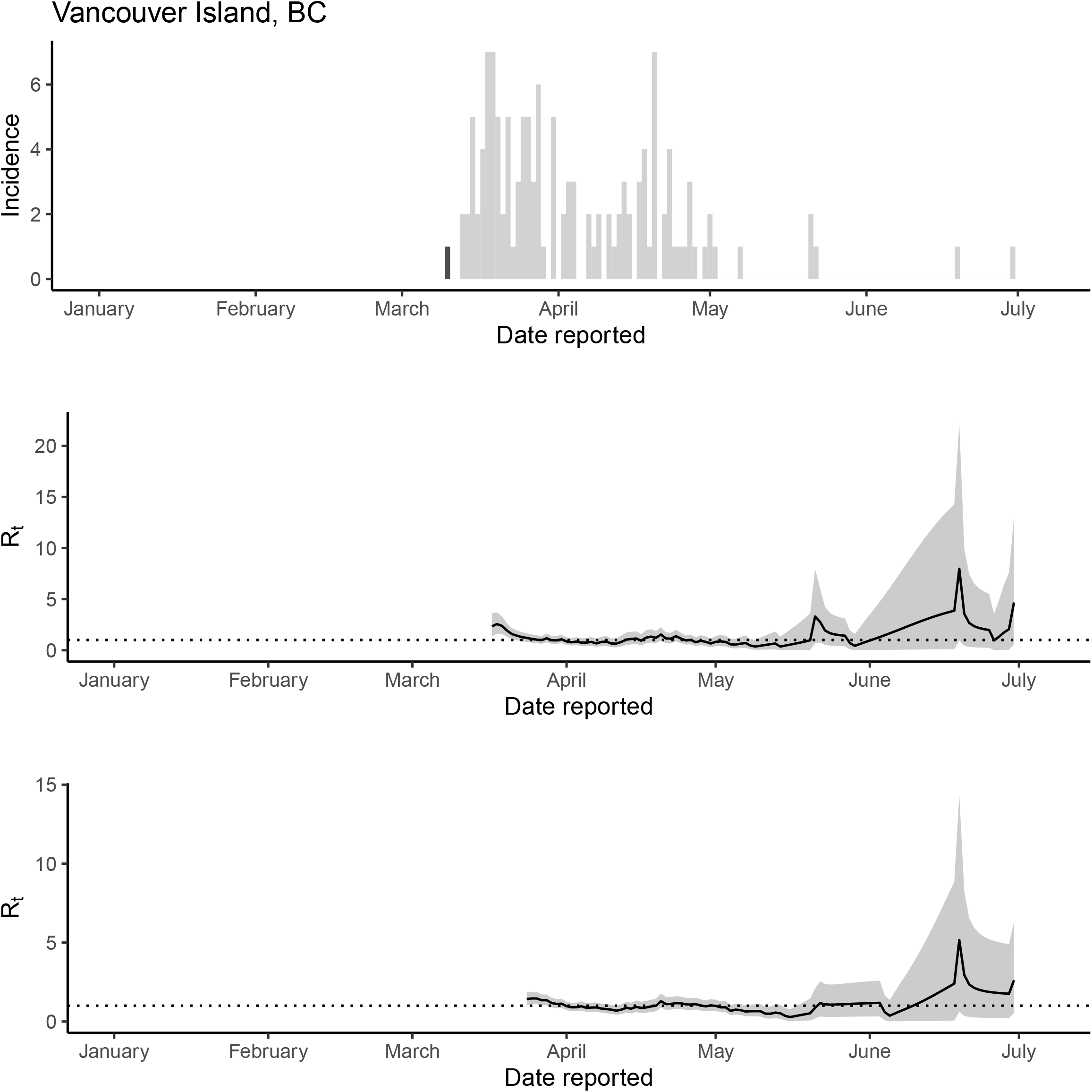
The epidemic trajectory of coronavirus disease 2019 in Vancouver Island Health Authority, British Columbia, January 1-July 6, 2020: Daily number of new cases by date of report (upper panel); *R_t_* with a 1-week window (middle panel); *R_t_* with a 2-week window (lower panel).

**Figure S41.**
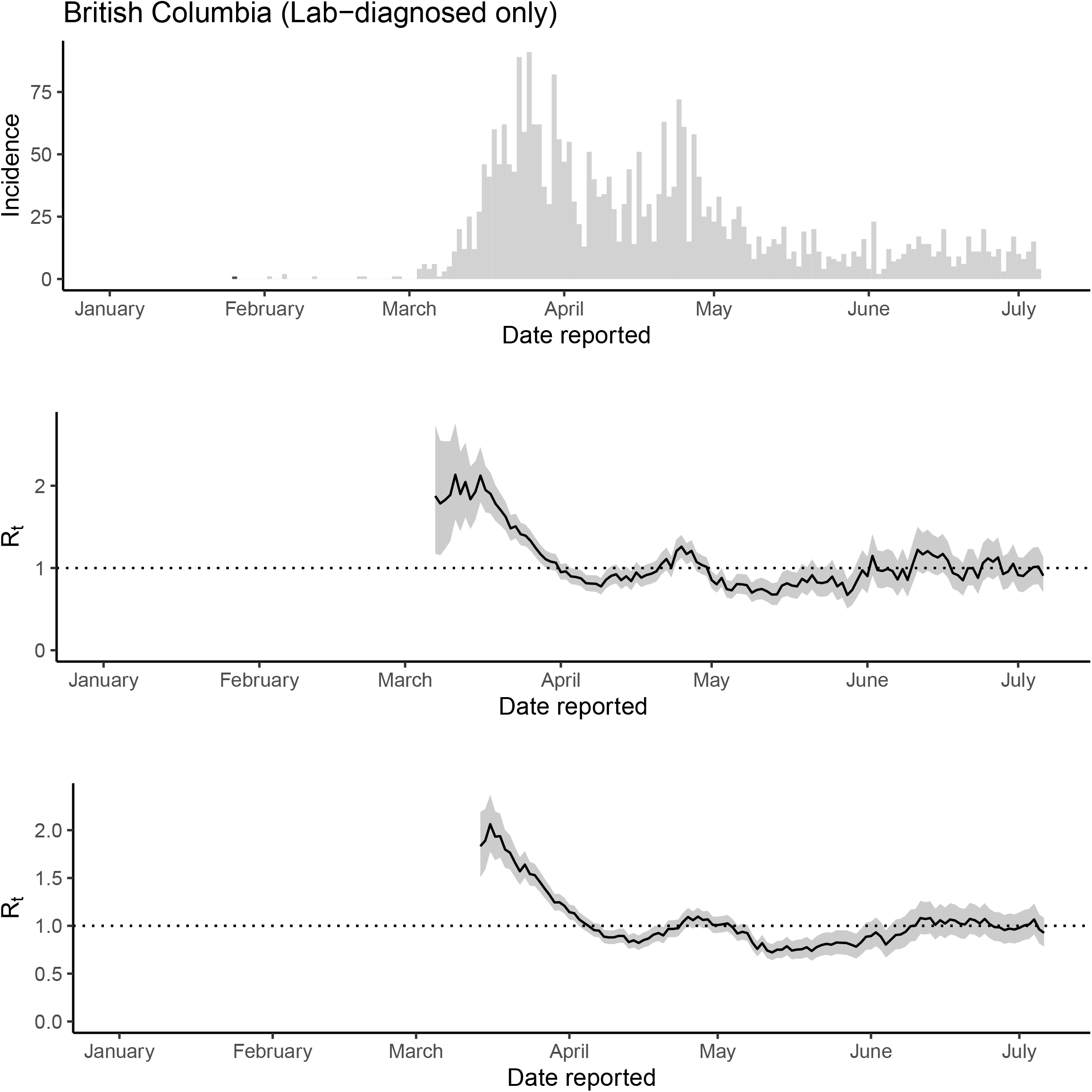
Sensitivity analysis (lab-diagnosed cases only) The epidemic trajectory of coronavirus disease 2019 in British Columbia, 2020: Daily number of new lab-diagnosed cases by date of report (upper panel), *R_t_* with a 1-week window (middle panel) and *R_t_* with a 2-week window (lower panel).

**Figure S42.**
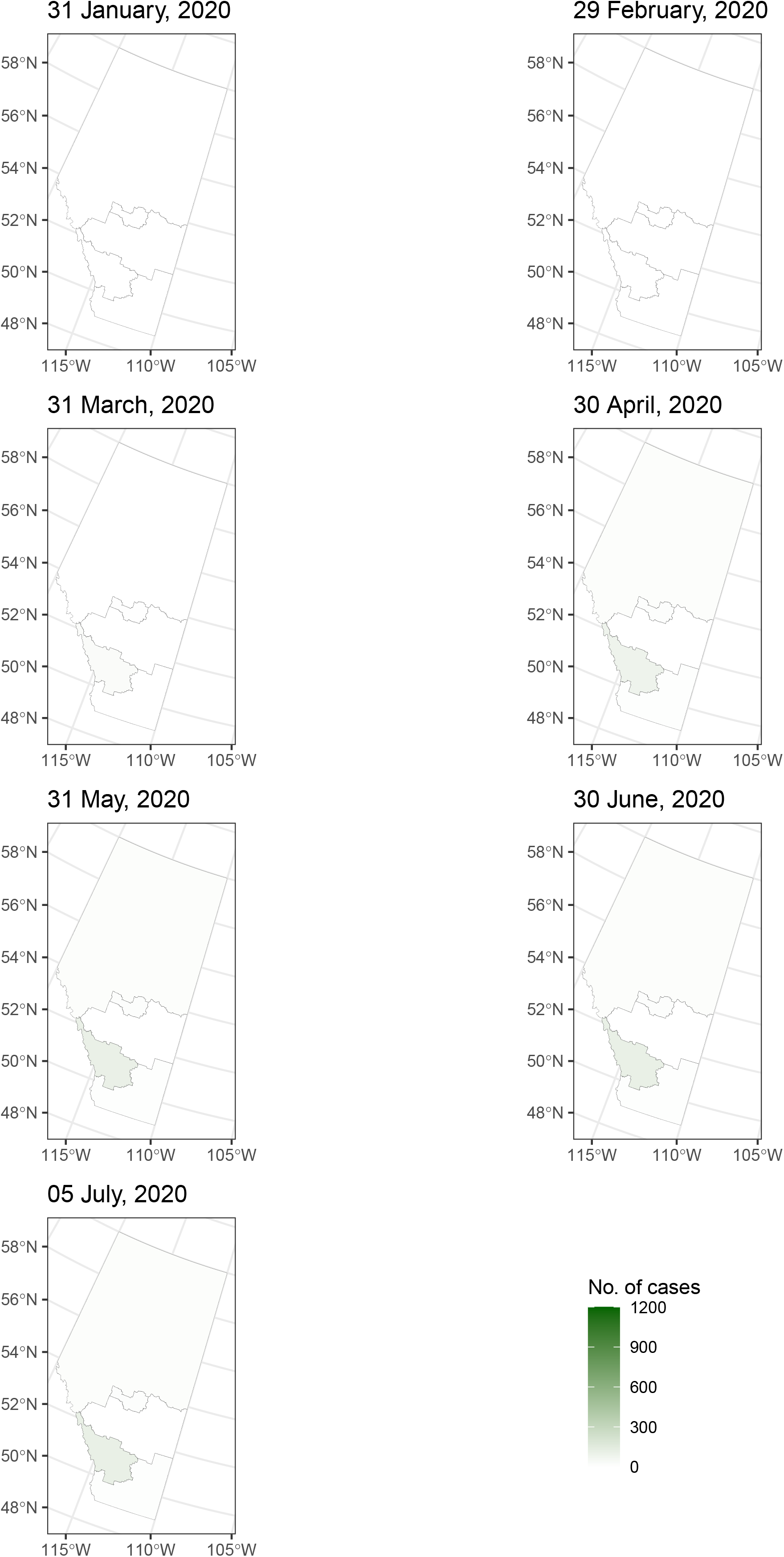
Map of cumulative number of deaths in Alberta by health service zone by the end of month and on July 6, 2020.

**Figure S43.**
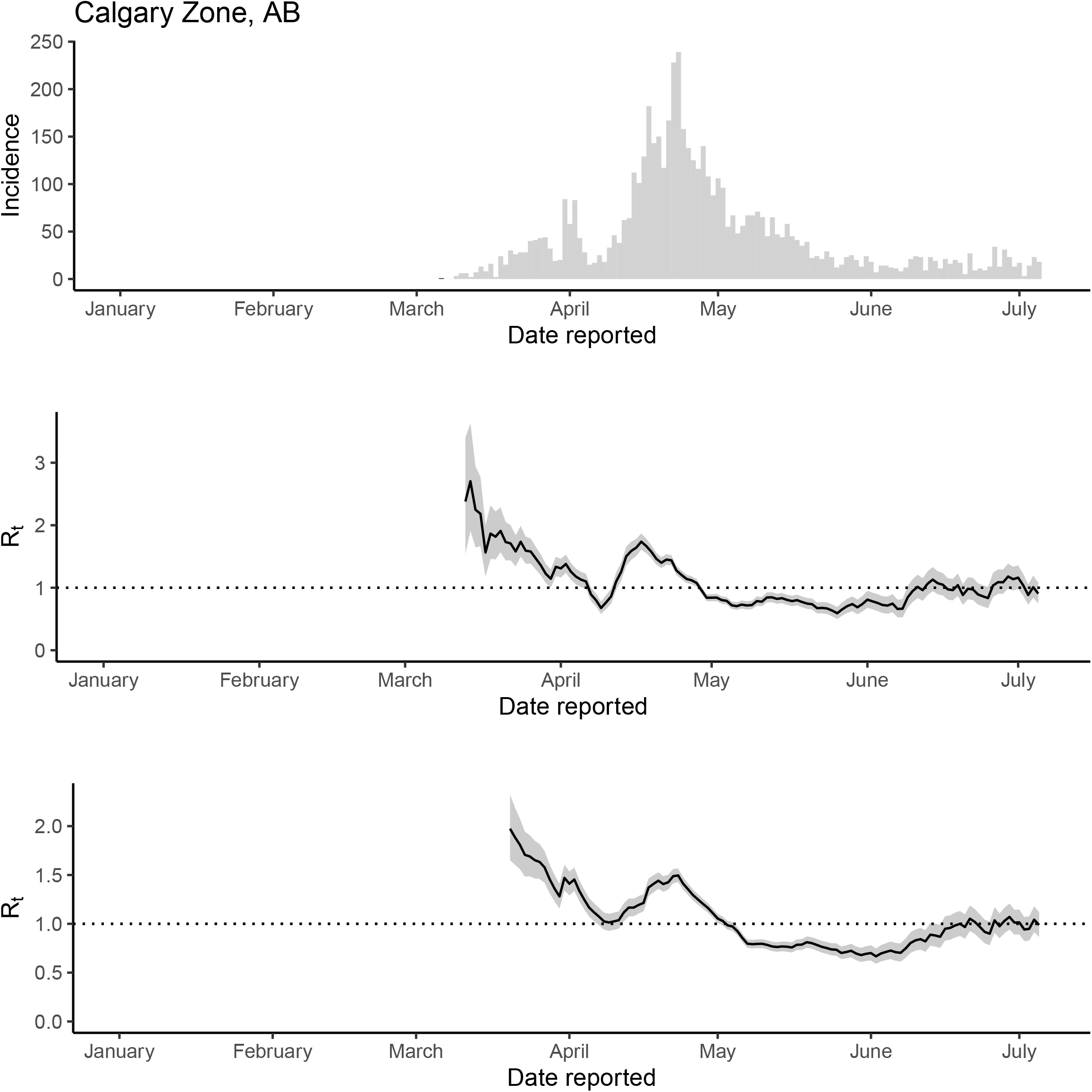
The epidemic trajectory of coronavirus disease 2019 in Calgary Zone, Alberta, January 1-July 5, 2020: Daily number of new cases by date of report (upper panel), *R_t_* with a 1-week window (middle panel) and *R_t_* with a 2-week window (lower panel).

**Figure S44.**
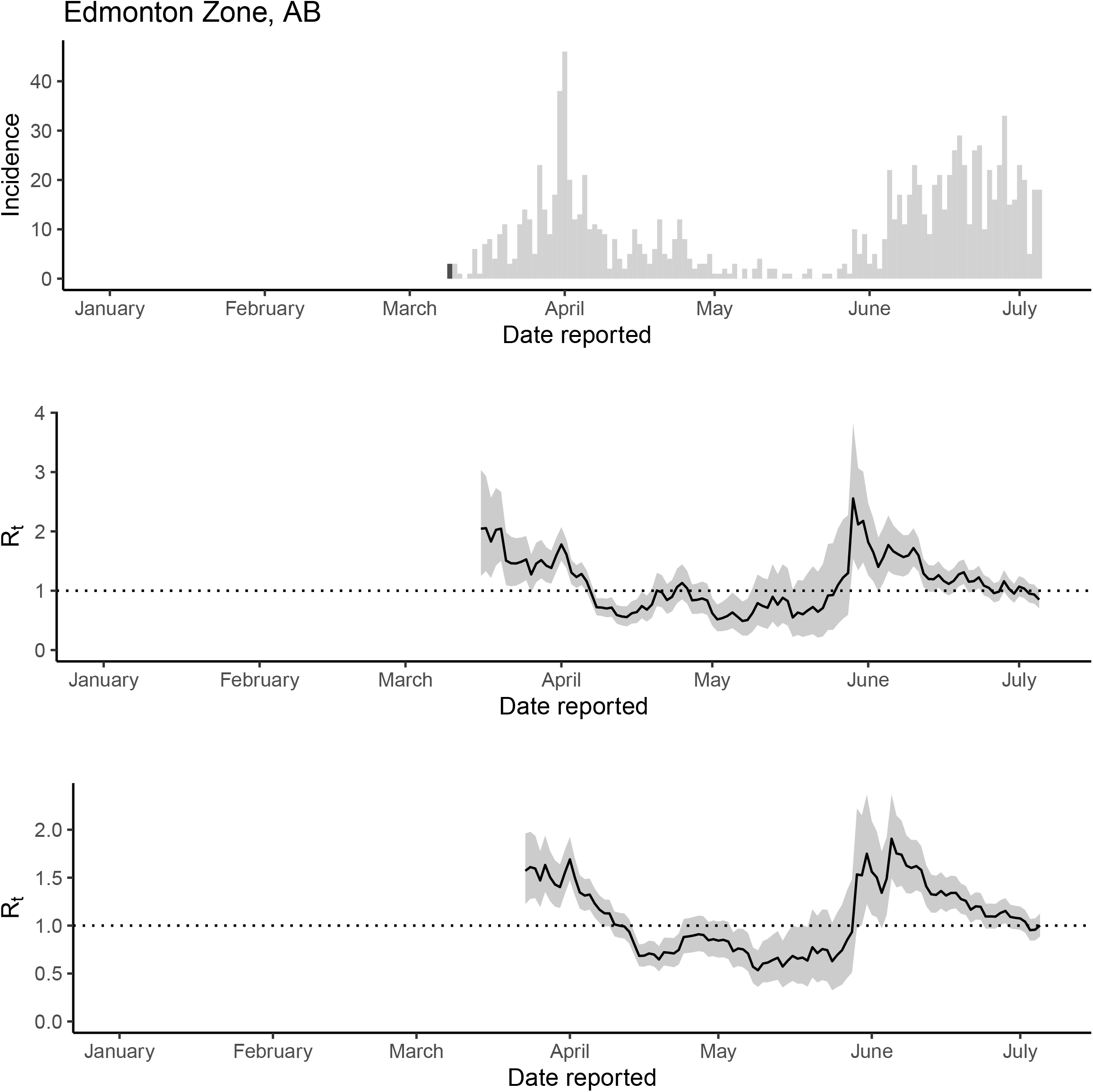
The epidemic trajectory of coronavirus disease 2019 in Edmonton Zone, Alberta, January 1-July 5, 2020: Daily number of new cases by date of report (upper panel), *R_t_* with a 1-week window (middle panel) and *R_t_* with a 2-week window (lower panel).

**Figure S45.**
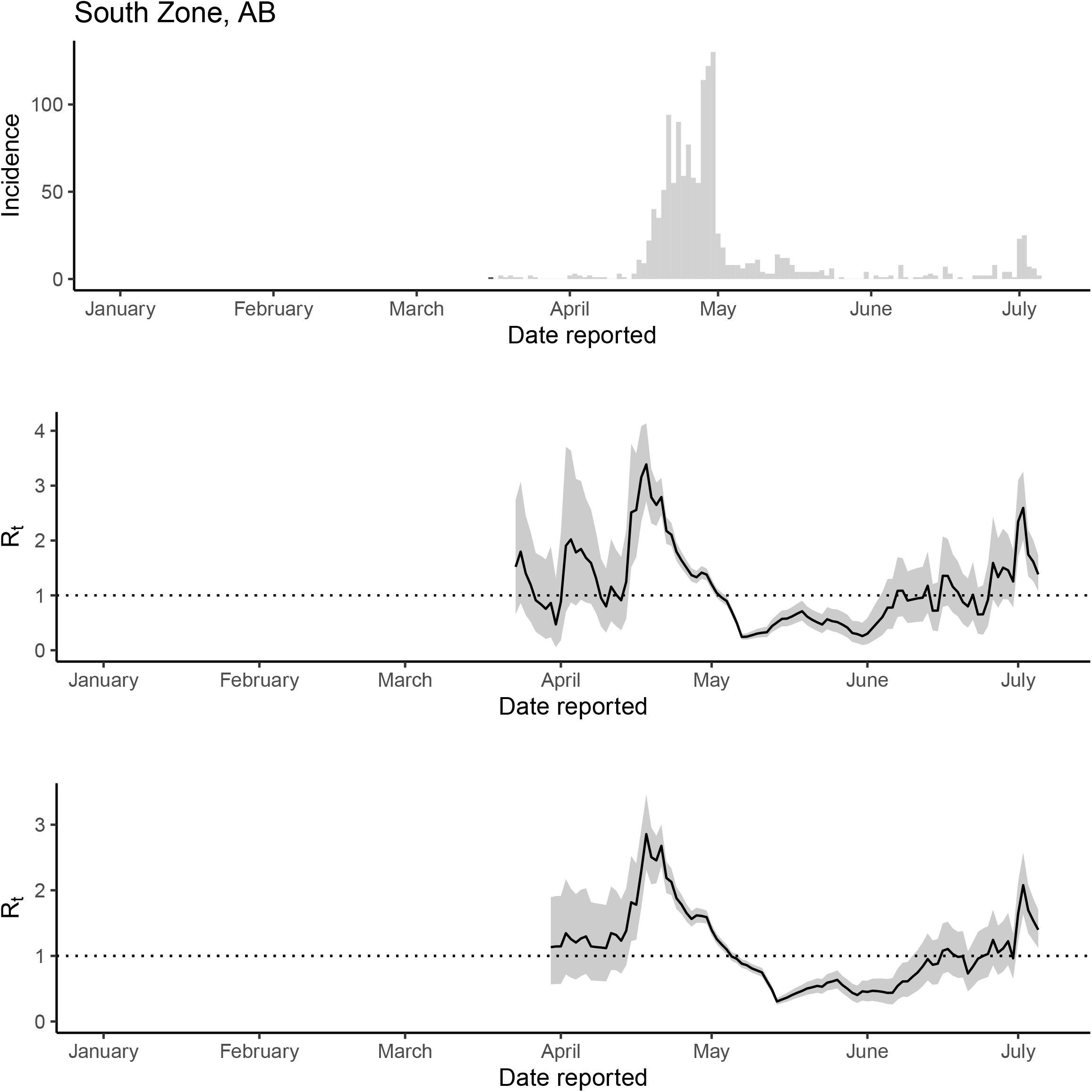
The epidemic trajectory of coronavirus disease 2019 in South Zone, Alberta, January 1-July 5, 2020: Daily number of new cases by date of report (upper panel), *R_t_* with a 1-week window (middle panel) and *R_t_* with a 2-week window (lower panel).

**Figure S46.**
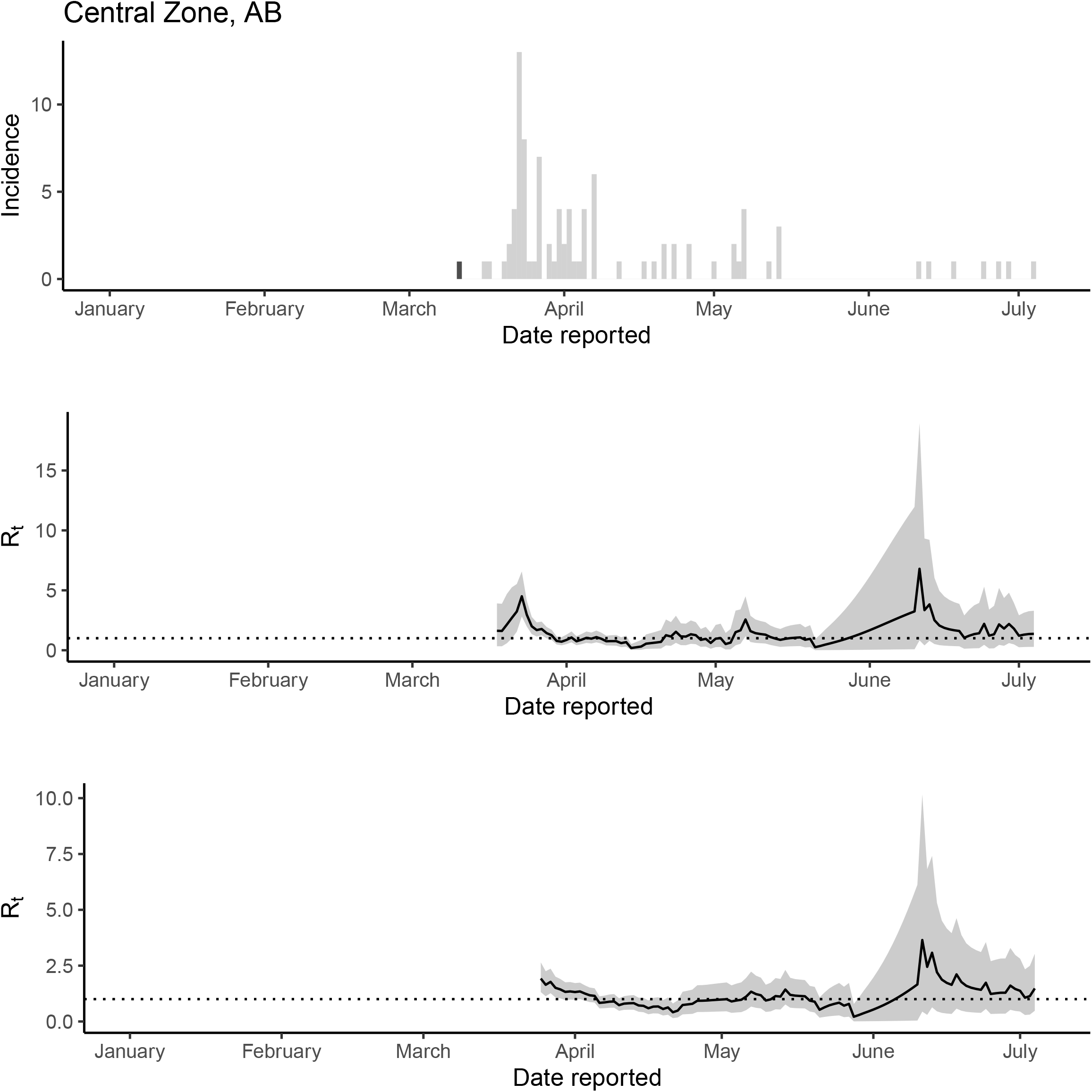
The epidemic trajectory of coronavirus disease 2019 in Central Zone, Alberta, January 1-July 5, 2020: Daily number of new cases by date of report (upper panel), *R_t_* with a 1-week window (middle panel) and *R_t_* with a 2-week window (lower panel).

**Figure S47.**
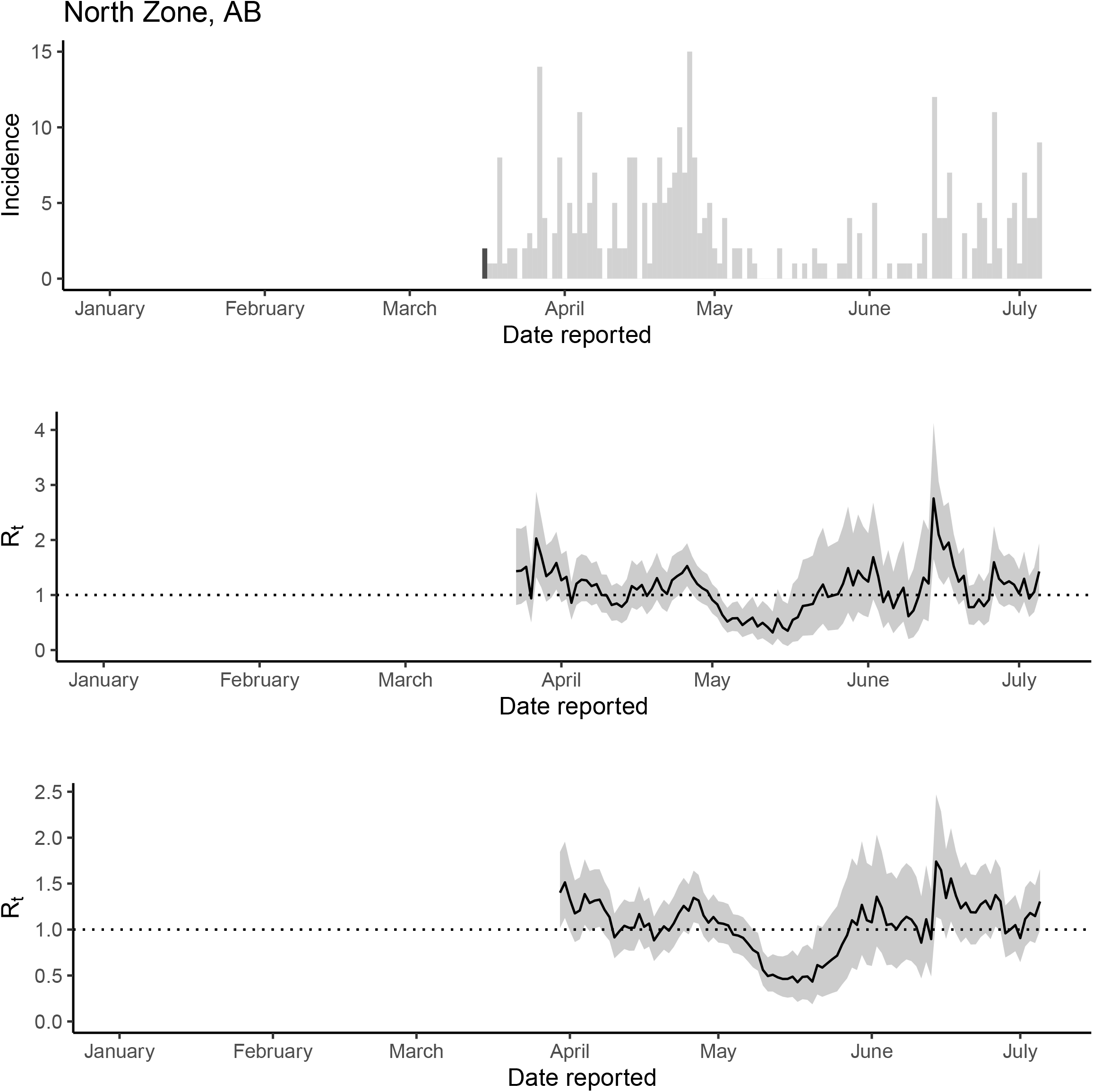
The epidemic trajectory of coronavirus disease 2019 in North Zone, Alberta, January 1-July 5, 2020: Daily number of new cases by date of report (upper panel), *R_t_* with a 1-week window (middle panel) and *R_t_* with a 2-week window (lower panel).

**Figure S48.**
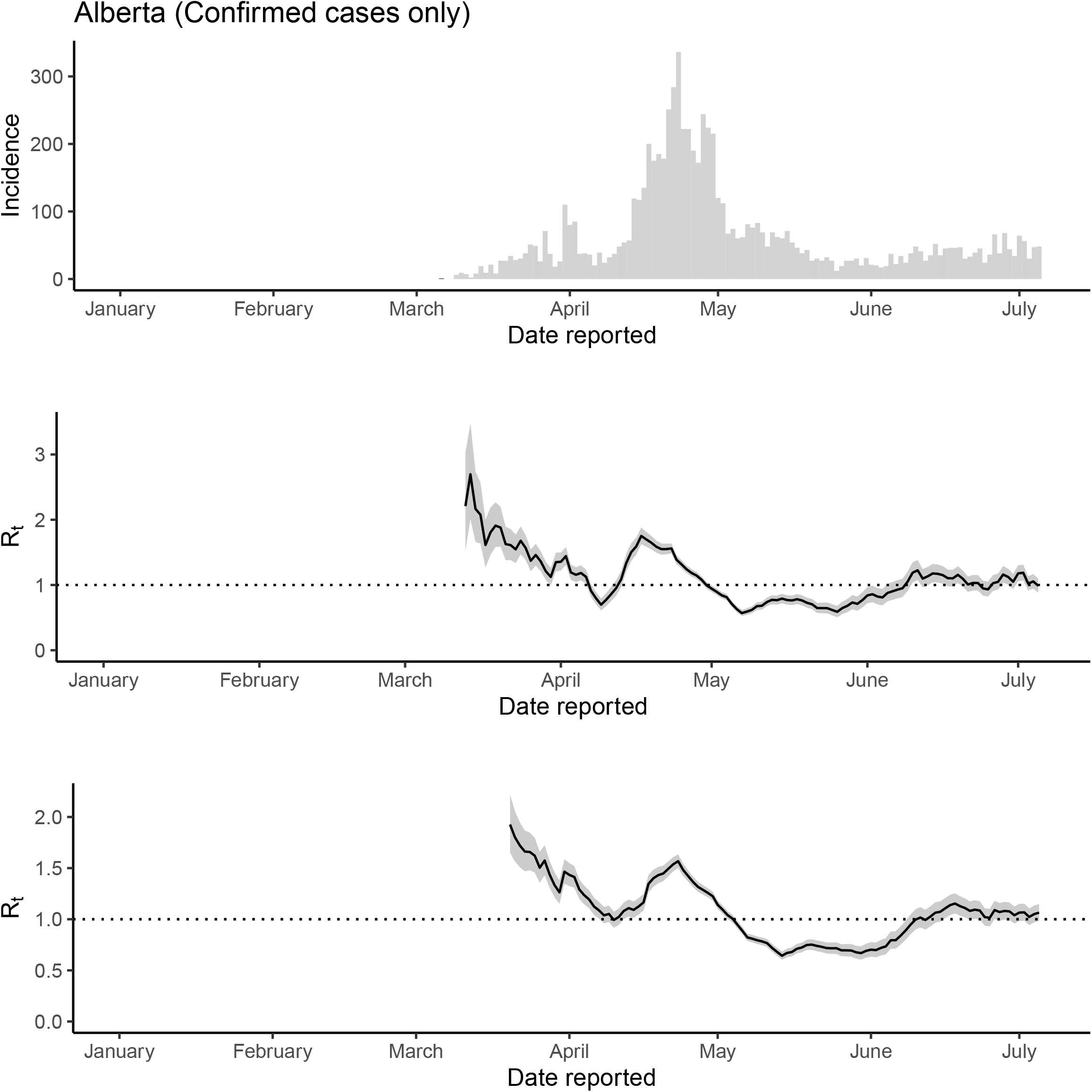
Sensitivity analysis (confirmed cases only) The epidemic trajectory of coronavirus disease 2019 in Alberta, 2020: Daily number of new confirmed cases by date of report (upper panel), *R_t_* with a 1-week window (middle panel) and *R_t_* with a 2-week window (lower panel).

**Figure S49.**
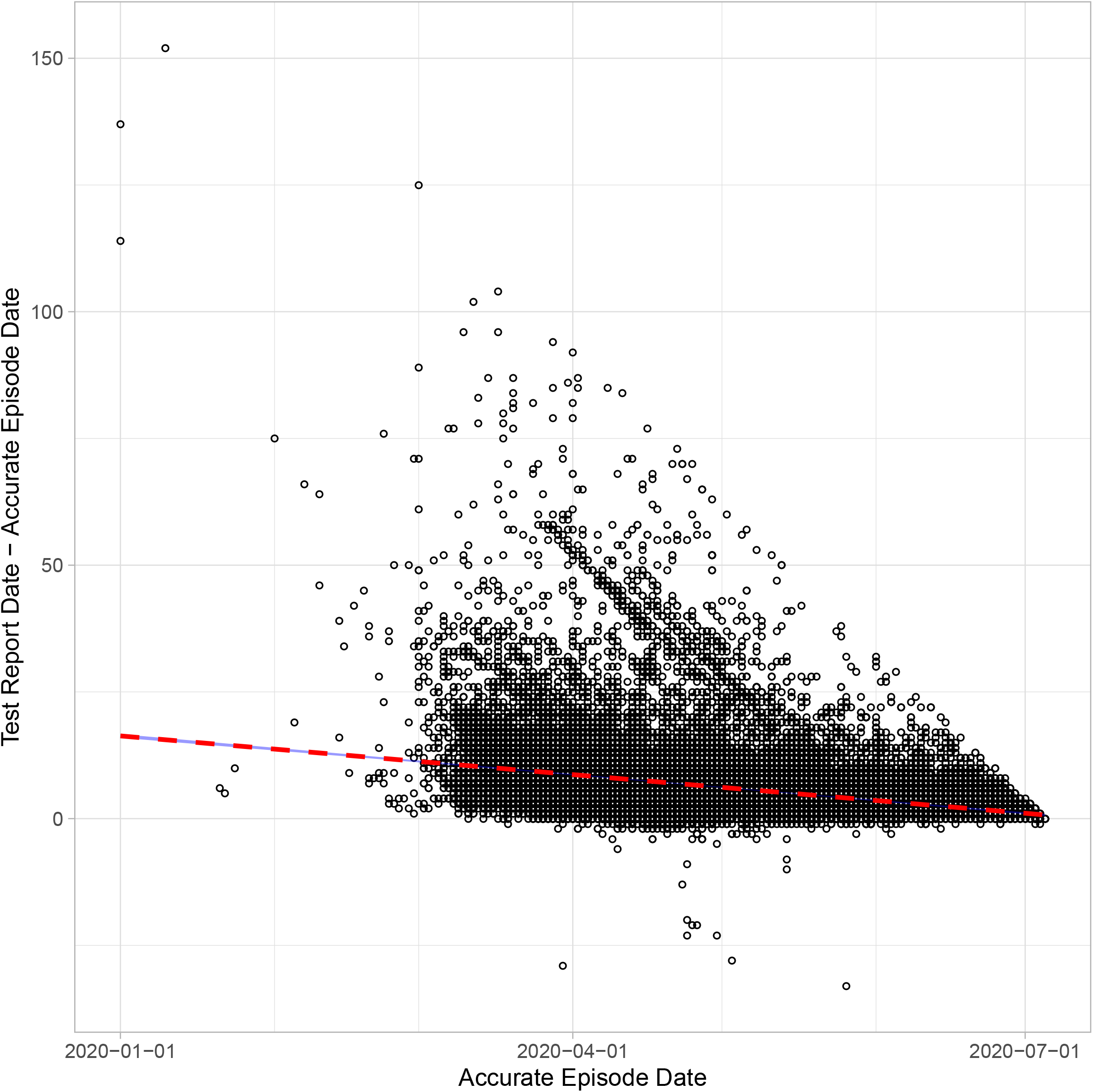
Scatter plot of the time difference between symptom onset (accurate episode date) and test report date of each case-patient in Ontario over time (accurate episode date), Jan 1-July 6, 2020.

